# Restoration of Gamma Center Frequency via Personalized Entrainment Marks Cognitive Preservation in Early Alzheimer’s Disease

**DOI:** 10.64898/2026.01.23.26344352

**Authors:** Yeseung Park, Hyeonwook Chae, Euisuk Yoon, Yejung Kim, Ji Won Han, Se Joon Woo, Seunghyup Yoo, Ki Woong Kim

## Abstract

**Background:** Gamma entrainment shows promise for Alzheimer’s disease (AD) treatment in preclinical models, but human trials have yielded heterogeneous results. We hypothesized that the clinical efficacy of gamma entrainment depends on individual neurophysiological receptivity, specifically the capacity for neural circuit plasticity.

**Methods:** In this open-label pilot study, we screened 37 individuals and enrolled 16 participants with early AD (CDR 0.5-1.0, amyloid-positive) who completed 12 weeks of home-based flickering light stimulation at individually optimized gamma frequencies (32-40 Hz). Pre- and post-intervention assessments included 64-channel EEG recordings and MMSE.

**Results:** Participants demonstrated dichotomous neurophysiological responses: 43.8% showed CF increase (ICF+) while 56.3% showed no change/decrease (ICF-). CF restoration was significantly associated with cognitive preservation (r=0.52, p=0.039). Notably, future responders exhibited distinct baseline signatures of “neural reserve,” characterized by higher temporal gamma power (Cohen’s d=0.70-0.92) and stronger frontotemporal connectivity (Cohen’s d=1.11-1.47). Almost 30% of screened candidates failed to show baseline entrainment, highlighting a distinct “non-responsive” biological subtype.

**Discussion:** CF restoration following personalized gamma entrainment identifies a neurophysiological subtype capable of meaningful plasticity. Rather than a universal remedy, gamma entrainment appears to act on specific neural substrates preserved in a subset of patients. These findings suggest that baseline electrophysiological profiling could unlock gamma entrainment’s therapeutic potential by stratifying likely responders for precision neuromodulation.

## Introduction

Alzheimer’s disease (AD) urgently requires novel therapeutic approaches that can modify disease progression without the risks associated with invasive interventions. Among emerging therapies, gamma entrainment through sensory stimulation has shown unprecedented success in animal models, yet translation to humans has proven challenging. While 40Hz sensory stimulation produces 40-60% amyloid reduction in transgenic mice through microglial activation and glymphatic clearance,^1–4^ human trials have yet to achieve comparable pathology reversal despite showing modest functional benefits.^5–8^

This translational gap likely stems from a fundamental oversight: the assumption that fixed 40Hz stimulation would be equally effective across biologically heterogeneous human populations. The OVERTURE trial, despite enrolling 76 participants for six months of daily 40Hz stimulation, failed to meet its primary cognitive endpoint and showed no measurable amyloid reduction.^7^ Similarly, Soula et al. demonstrated that only 25% of visual cortex neurons in AD model mice entrained to the supposedly optimal 40Hz frequency,^9^ underscoring that successful gamma entrainment requires precision matching between stimulation parameters and individual neural characteristics.

The neurobiology of gamma oscillations provides a clear rationale for personalized intervention. Gamma oscillations emerge from precisely timed interactions between excitatory pyramidal neurons and parvalbumin-positive (PV+) inhibitory interneurons, creating the temporal framework for cognitive processing.^1, 10^ In AD, this delicate synchrony deteriorates years before clinical symptoms, and the dominant frequency within the gamma band, i.e., the center frequency (CF), shifts progressively downward as interneurons succumb to metabolic stress and ion channel dysfunction.^11–13^ Reduced gamma power and altered center frequencies are directly correlated with memory deficits.^10, 11, 13^

We hypothesize that the capacity to restore CF following entrainment represents a marker of “neural reserve” defined as the remaining functional integrity of PV+ interneuron circuits that enables adaptive plasticity in response to external stimulation. This concept draws on evidence that CF is a modifiable parameter, but only within specific physiological limits determined by the metabolic and structural integrity of inhibitory networks.^14, 15^ In aging and early AD, successful entrainment depends on matching stimulation frequency to individual baseline CF, as circuits can only be “pulled” within a limited range of their natural frequency.^16, 17^ Indeed, upward CF shifts following motor learning and cognitive training have served as biomarkers for functional improvement,^20^ suggesting that frequency modulation reflects genuine circuit-level restoration. This framework predicts that patients can be stratified by their baseline electrophysiological signatures. Specifically, we posit that elevated gamma power, often manifesting as compensatory hyperactivity in early AD,^18, 19^ may paradoxically indicate preserved neural substrate capable of normalization through entrainment. In contrast, “hypoactive” patterns may reflect systems that have crossed the threshold of recoverable function, where insufficient PV+ interneuron integrity precludes adaptive reorganization.

To test these hypotheses, we designed a 12-week proof-of-concept study employing personalized gamma entrainment at individually optimized frequencies (32-40 Hz) in early AD patients. Specifically, we examined whether baseline “neural reserve” (manifesting as compensatory gamma activity) predicts entrainment success, and whether the restoration of CF serves as a biomarker for cognitive preservation. This approach represents a fundamental departure from fixed-frequency protocols, shifting the paradigm from universal stimulation parameters toward precision biomarkers for patient stratification and treatment monitoring in AD neuromodulation.

## Materials and methods

### Study Design

This open-label, single-arm pilot study evaluated the effects of 12-week personalized gamma entrainment using flickering light stimulation at individualized center frequency (FLS-CF) on electrophysiological and cognitive outcomes in individuals with early AD. This study was conducted at Seoul National University Bundang Hospital from February 2023 to February 2024. The study protocol was reviewed and approved by the Seoul National University Bundang Hospital Institutional Review Board (Seongnam, Gyeonggi-do, Republic of Korea; IRB No. B-2302-810-302). All participants or their legal representatives provided written informed consent before participating in the study.

The study was registered in the Clinical Research Information Service (CRIS), Republic of Korea (Registration No. KCT0010618).

This paper presents a focused analysis of electrophysiological changes and their relationship to cognitive outcomes. Given the novel finding of CF change as a potential biomarker for treatment response, we prioritized in-depth analysis of EEG-based measures and their mechanistic implications.

### Participants

We screened 37 individuals aged 50 years or older with mild cognitive impairment (MCI) or mild dementia due to AD. Participants were recruited from the outpatient geriatric psychiatry clinic and through hospital-wide advertisements. Geriatric neuropsychiatrists specialized in dementia research conducted a face-to-face standardized diagnostic interview using the Korean version of the Consortium to Establish a Registry for Alzheimer’s Disease Assessment Packet (CERAD-K) Clinical Assessment Battery^21^ and the Korean version of the Mini International Neuropsychiatric Interview (MINI-K),^22^ as well as physical and neurological examinations and laboratory tests including complete blood counts, chemistry profiles, and serological tests for syphilis. Neuropsychologists or trained research nurses administered the Korean version of Mini-Mental State Examination (MMSE) and the Korean version of Geriatric Depression Scale (GDS).^23^

All participants underwent ^18^F-florbetaben PET imaging to confirm amyloid positivity. Images were acquired 90-110 minutes after injection of 300 MBq ^18^F-florbetaben. Quantitative analysis was performed to calculate the global standardized uptake value ratio (SUVR) using the cerebellar cortex as reference region, and brain amyloid plaque load (BAPL) scores were determined on a 1-3 scale. Amyloid positivity was defined as a BAPL score of 2 or 3, as evaluated by a board-certified nuclear medicine physician.

A panel of four geriatric psychiatrists determined diagnoses and Clinical Dementia Rating (CDR) scores through consensus conferences. We diagnosed mild cognitive impairment (MCI) according to the consensus criteria from the International Working Group on MCI,^24^ dementia according to the Diagnostic and Statistical Manual of Mental Disorders, Fourth Edition (DSM-IV) diagnostic criteria,^25^ and Alzheimer’s disease according to the National Institute on Aging - Alzheimer’s Association (NIA-AA) criteria for Alzheimer’s disease.^26^ Inclusion criteria were: (1) age ≥50 years; (2) literacy and ability to provide written informed consent; (3) corrected binocular visual acuity ≥0.5; (4) reliable caregiver available for ≥8 hours per week; (5) diagnosis of MCI due to AD or mild AD dementia with CDR of 0.5-1.0; and (6) evidence of amyloid positivity on ^18^F-florbetaben PET imaging. Exclusion criteria included: (1) retinal or optic nerve conditions contraindicated for light stimulation; (2) major psychiatric, neurological, or physical conditions or severe sensory deficits affecting cognition; (3) baseline EEG entrainment failure (signal-to-noise ratio [SNR] <1.5 at occipital electrodes during frequency scan); and (4) any condition judged to interfere with study participation.

Written informed consent was obtained from all participants or their legal guardians prior to enrollment.

### Intervention Protocol

The intervention consisted of 12 weeks of home-based FLS-CF delivered through custom-built eyewear containing OLED panels. Prior to intervention, we determined each participant’s optimal stimulation frequency during baseline EEG assessment. We tested gamma entrainment responses at five frequencies (32, 34, 36, 38, and 40 Hz) and selected the frequency yielding the highest event-related synchronization (ERS) across occipital channels (Oz, POz, O1, O2) as the individualized CF. ^17^

Participants self-administered FLS-CF twice daily (30 minutes per session) for at least 5 days per week, with device parameters and safety monitoring detailed below.

### OLED-based FLS device system and Operation

We developed a custom eyewear device using organic light-emitting diode (OLED) technology (A88MA2B; Konica Minolta Inc., Tokyo, Japan). The OLED panels were mounted in a 3D-printed frame positioned 2 cm from each pupil, delivering square-wave modulated white light (5700K color temperature, 1120 cd/m² luminance, 50% duty cycle) at programmable frequencies (32, 34, 36, 38, and 40 Hz). The device incorporated multiple safety features including thermal management, battery backup, and automatic power-off functionality. All 20 devices demonstrated consistent performance within ±2.5% luminance variation. (**Supplementary Appendix 1)**.

For the home-based intervention, each device was pre-programmed with the participant’s individualized frequency. A tablet application connected via Bluetooth Low Energy (BLE) initiated 30-minute stimulation sessions and automatically logged usage data including session timing, duration, and completion status, enabling remote monitoring of adherence.

For baseline and post-intervention EEG assessments, the device operation was modified to enable precise experimental control. A function generator (TG 5012A, Aim & Thurlby Thandar Instruments, Huntingdon, UK) replaced the standard AC circuit to ensure exact timing for steady-state visual evoked potential (SSVEP) measurements. During these assessments, all five frequencies were tested in randomized order with voltage fluctuations controlled within 5 mV. Each assessment session began with a 5-minute resting-state recording, followed by a 3-minute break, then 10 experimental blocks containing 10 trials each. Individual trials lasted 2 seconds with randomized inter-trial intervals of 3-6 seconds to minimize habituation effects. This protocol enabled systematic identification of each participant’s optimal entrainment frequency based on maximal ERS across occipital channels (Oz, POz, O1, O2).^17, 27, 28^

### Safety and Tolerability Assessment

Participants rated six potential adverse effects after each session (fatigue, headache, dizziness, dazzling sensation, asthenopia, ocular pain) on a 7-point Likert scale (0 = not at all, 6 = extremely severe). Moderate-to-severe adverse effects were defined as severity scores ≥3. Research staff conducted weekly phone calls to monitor adherence and safety.

### Outcome Measures

The present analysis focused on three main outcome categories from the registered clinical trial protocol (CRIS Registration No. KCT0010618). Electrophysiological changes were evaluated as primary outcomes using quantitative EEG (QEEG) and event-related spectral perturbation (ERSP) measurements. Pre- and post-intervention assessments included the following: (1) resting-state low gamma power (32 - 40 Hz); (2) gamma-band ERS during flickering light stimulation; (3) directional functional connectivity measured by changes in spectral Granger causality (ΔsGC); and (4) CF of gamma oscillations. These measures were derived from 64-channel EEG recordings collected before and after the 12-week personalized FLS-CF intervention.

Cognitive function was assessed as a secondary outcome using the Mini-Mental State Examination (MMSE), administered by trained neuropsychologists following standardized protocols. Assessments were conducted at baseline and immediately after the 12-week intervention period. Treatment adherence was calculated as the percentage of completed sessions relative to the 120 prescribed sessions, monitored through automated device logs.

### Recording, Preprocessing and Analysis of EEG

EEG data were recorded using 64-channel Ag–AgCl electrodes caps (Easycap, EASYCAP GmbH, Munich, Germany) positioned according to the extended International 10–20 System. FCz served as the reference electrode, with the ground electrode placed on the forehead. Vertical electrooculogram (VEOG) was recorded using a pair of electrodes positioned above and below the left eye to monitor ocular artifacts. Electrode impedances were maintained below 10 kΩ throughout all recording sessions. Signals were amplified and digitized using a 24-bit ActiCHamp DC amplifier and recorded with BrainVision Recorder software (Brain Products GmbH, Gilching, Germany) at a sampling rate of 1,000 Hz without online filtering. Stimulus markers from the FLS control system were synchronized with the EEG recording for precise temporal alignment.

Data preprocessing and analysis were performed using MATLAB (The MathWorks Inc., Natick, MA, USA) with EEGLAB toolbox and BSMART toolbox for connectivity analysis. The recorded signals underwent several preprocessing steps including application of a 1-Hz high-pass finite impulse response filter to remove slow drifts and a 60-Hz notch filter to eliminate line noise. The data were then re-referenced to the common average of all channels. Independent component analysis (ICA) was performed to identify and remove components related to eye blinks and other ocular artifacts.

Following preprocessing, the continuous data were segmented for different analyses. For resting-state analysis, the 5-minute recordings were divided into 1,500-ms epochs, from which 20 artifact-free epochs were randomly selected. For FLS response analysis, we extracted 4,000-ms epochs spanning from 1,000 ms before stimulus onset to 1,000 ms after stimulus offset, yielding 20 epochs per frequency condition. To analyze steady-state responses while avoiding onset transients, we selected a 1,500-ms window from 501 to 2,000 ms post-stimulus for spectral Granger causality (sGC) analysis, ensuring stable entrainment while excluding initial evoked responses.

We determined each participant’s optimal stimulation frequency through systematic testing of gamma entrainment responses. Signal-to-noise ratio (SNR) was calculated as the ratio of steady-state visual evoked potential (SSVEP) power at the stimulus frequency to mean background power (1-60 Hz) on a logarithmic scale. Participants with SNR <1.5 at any occipital electrode (POz, Oz, O1, O2) were excluded as non-responders.^17, 28^

For frequency optimization, we computed ERSP during 2-second FLS at five frequencies (32, 34, 36, 38, 40 Hz). The frequency producing maximal ERS averaged across occipital channels was selected as the participant’s individualized CF.^17, 28^ This CF served as the stimulation frequency throughout the 12-week intervention.

Four primary electrophysiological measures were assessed pre- and post-intervention: (1) Resting-state gamma power (RSP; 32-40 Hz): Quantified using fast Fourier transform on 20 artifact-free 1.5-second epochs from the 5-minute resting-state recording; (2) Gamma-band ERS: Computed as the normalized power change during FLS relative to pre-stimulus baseline, analyzing the 501-2000 ms window when steady-state entrainment was established; (3) CF: Re-assessed post-intervention using identical procedures, with CF change (ΔCF) calculated as post-minus pre-intervention values; (4) Directional functional connectivity (ΔsGC): Measured using spectral Granger causality between all electrode pairs during both resting-state and FLS conditions; ΔsGC quantified FLS-induced connectivity modulation.

For connectivity analyses, electrodes were grouped into 13 regions^29^: prefrontal (pF), left/right frontal (LF/RF), frontal midline (F), left/right temporal (LT/RT), left/right central (LC/RC), central midline (C), left/right parietal (LP/RP), parietal midline (P), and occipital (O). All analyses were performed using MATLAB with EEGLAB and BSMART toolboxes, focusing on hypothesis-driven regions and frequency bands to minimize multiple comparison issues while maintaining sensitivity to detect clinically meaningful changes.

## Statistical Analysis

Participants were stratified post-hoc based on CF change: CF-increased (ICF+, n = 7) versus CF-not-increased (ICF-, n = 9). Baseline demographic and clinical characteristics were compared between groups using Mann-Whitney U test for continuous variables and Fisher’s exact test for categorical variables. Exploratory comparisons between enrolled (n = 16) and excluded participants (n = 11, insufficient entrainment) used Welch’s t-test and Fisher’s exact test.

Outcomes were analyzed using repeated-measures ANOVA with time (pre vs. post-intervention) as within-subject factor and group (ICF+ vs. ICF-) as between-subject factor. Main effects of time, group, and time × group interactions were evaluated for resting-state gamma power (RSP), gamma-band ERS, directional functional connectivity (ΔsGC), and MMSE score. Effect sizes were calculated using omega squared (ω²; small > 0.01, medium > 0.06, large > 0.14) for ANOVA and Cohen’s d for pairwise comparisons. For significant interactions, simple main effects were examined using paired t-tests (within-group) and independent t-tests (between-group).

The relationship between CF change and cognitive outcomes was evaluated using multiple approaches: (1) Logistic regression assessed the association between CF change (ICF+ vs. ICF-) and cognitive maintenance/improvement (defined as MMSE change ≥ 0); and (2) Receiver operating characteristic (ROC) analysis determined the optimal CF change cutoff for predicting cognitive preservation, with bootstrapped 95% confidence intervals (1000 resamples).

To identify baseline predictors of treatment response, univariate logistic regression with leave-one-out cross-validation (LOOCV) was performed for each EEG feature independently. Model performance was evaluated using ROC analysis with the optimal threshold determined by the Youden Index. Performance metrics included AUC, sensitivity, specificity, and odds ratios (ORs) with 95% confidence intervals from 1000 bootstrap resamplings. Spearman correlation examined relationships between treatment adherence and outcomes.

All statistical analyses were performed using MATLAB (The MathWorks Inc., Natick, MA, USA) with a significance threshold of p < 0.05. Given the exploratory nature of this pilot study and the emphasis on effect sizes to inform future trial designs, we applied false discovery rate (FDR) correction using the Benjamini–Hochberg procedure for electrode- or region-wise comparisons. Both uncorrected p-values and FDR-adjusted q-values are reported throughout the text and figures, with results that did not survive correction explicitly described as exploratory. For the LOOCV-based logistic regression analysis, statistical significance of odds ratios was also assessed using both uncorrected and FDR-adjusted p-values. For all other analyses, nominal p-values are reported unless otherwise specified.

## Data availability

The data that support the findings of this study are available from the corresponding author upon reasonable request.

## Results

### Participants

Of the 37 individuals who were screened for eligibility, 17 participants were excluded due to insufficient baseline gamma entrainment (n = 11, defined as SNR < 1.5 at occipital electrodes), ophthalmologic contraindications (n = 1), withdrawal of consent (n = 4), or protocol noncompliance (n = 1; refusal to undergo required MRI). Of the 20 participants who met the inclusion criteria and enrolled in the study, 16 successfully completed the 12-week intervention (**Figure 1**). Four participants were discontinued due to withdrawal of consent (n = 2) or unrelated medical events (nephrectomy, n = 1; wrist surgery, n = 1).

**Figure. 1.**
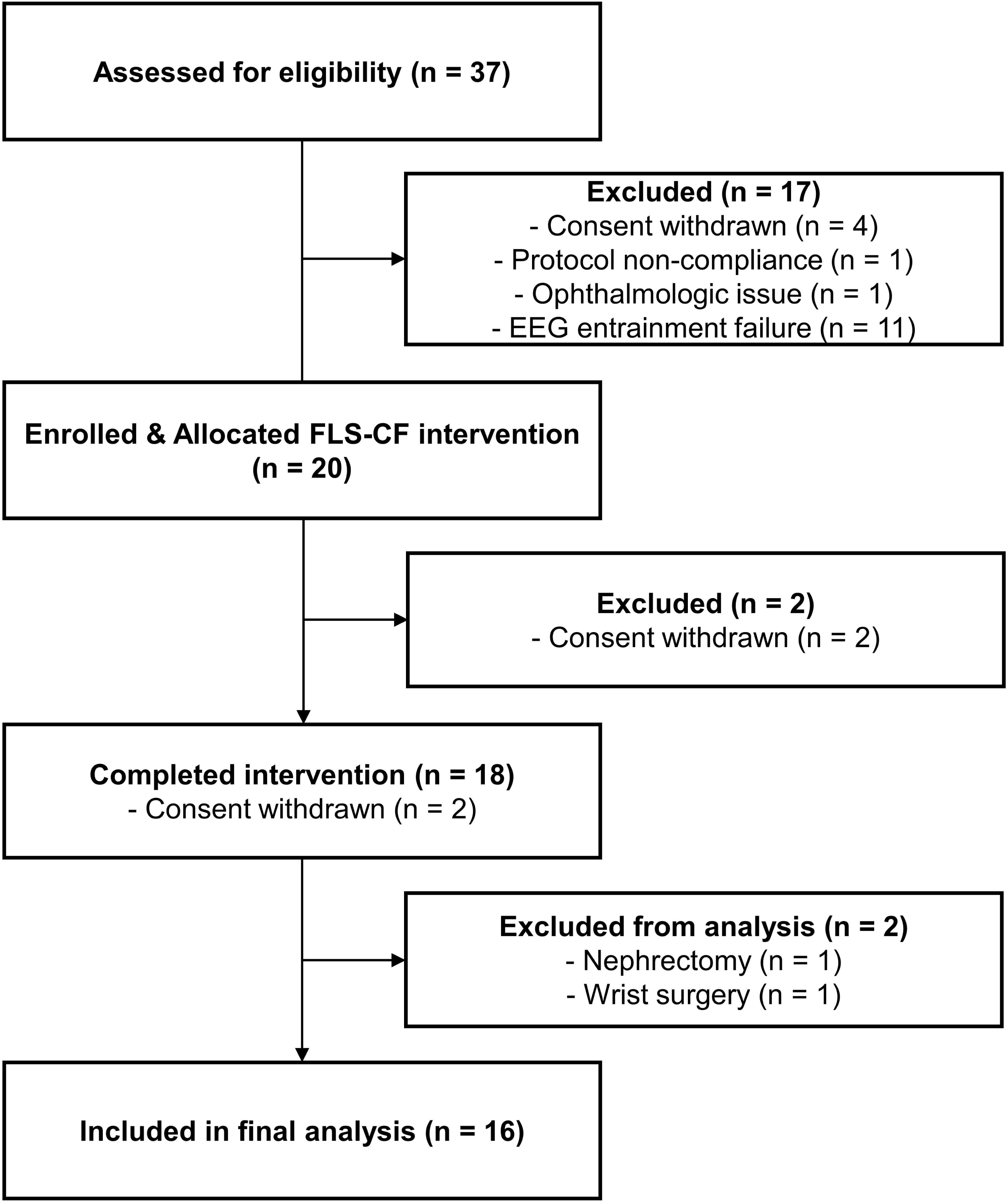
CONSORT flow diagram of participant enrollment, allocation, and analysis.

Baseline demographic and clinical characteristics are presented in **Table 1**. The 16 participants who completed the 12-week intervention comprised 9 women and 7 men. Their mean age was 74.8 ± 6.8 years and the mean years of education was 13.0 ± 3.8 years. All participants met the criteria for early-stage AD, as indicated by a mean MMSE score of 22.0 ± 4.9, a CDR global score of 0.47 ± 0.13, and confirmed amyloid positivity on ¹⁸F-florbetaben PET (mean global SUVR: 1.38 ± 0.26; mean BAPL score: 2.69 ± 0.60). Among them, 43.8% (seven participants) carried the APOE ε4 allele.

**Table 1.**
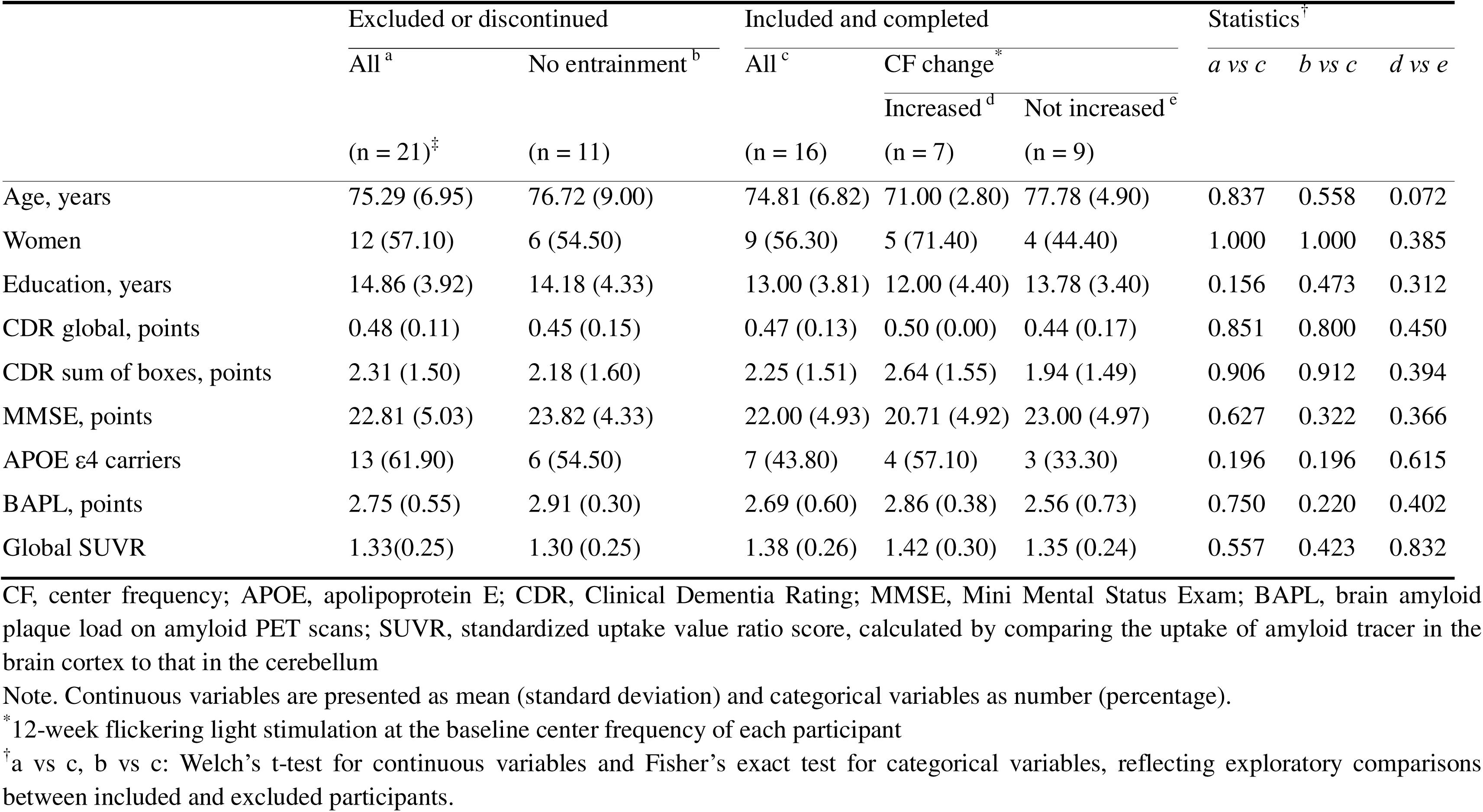
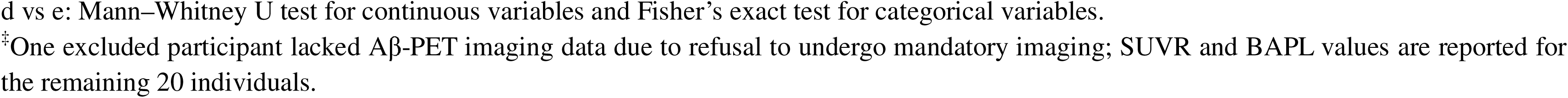
Baseline demographic and clinical characteristics of study participants stratified by enrollment status and center frequency response.

|  | Excluded or discontinued |  | Included and completed |  |  | Statistics<sup>†</sup> |  |  |
| --- | --- | --- | --- | --- | --- | --- | --- | --- |
|  | All<sup>a</sup> | No entrainment<sup>b</sup> | All<sup>c</sup> | CF change<sup>*</sup> |  | *a* vs *c* | *b* vs *c* | *d* vs *e* |
|  | (n = 21)<sup>‡</sup> | (n = 11) | (n = 16) | Increased<sup>d</sup> | Not increased<sup>e</sup> |  |  |  |
|  |  |  |  | (n = 7) | (n = 9) |  |  |  |
| Age, years | 75.29 (6.95) | 76.72 (9.00) | 74.81 (6.82) | 71.00 (2.80) | 77.78 (4.90) | 0.837 | 0.558 | 0.072 |
| Women | 12 (57.10) | 6 (54.50) | 9 (56.30) | 5 (71.40) | 4 (44.40) | 1.000 | 1.000 | 0.385 |
| Education, years | 14.86 (3.92) | 14.18 (4.33) | 13.00 (3.81) | 12.00 (4.40) | 13.78 (3.40) | 0.156 | 0.473 | 0.312 |
| CDR global, points | 0.48 (0.11) | 0.45 (0.15) | 0.47 (0.13) | 0.50 (0.00) | 0.44 (0.17) | 0.851 | 0.800 | 0.450 |
| CDR sum of boxes, points | 2.31 (1.50) | 2.18 (1.60) | 2.25 (1.51) | 2.64 (1.55) | 1.94 (1.49) | 0.906 | 0.912 | 0.394 |
| MMSE, points | 22.81 (5.03) | 23.82 (4.33) | 22.00 (4.93) | 20.71 (4.92) | 23.00 (4.97) | 0.627 | 0.322 | 0.366 |
| APOE ε4 carriers | 13 (61.90) | 6 (54.50) | 7 (43.80) | 4 (57.10) | 3 (33.30) | 0.196 | 0.196 | 0.615 |
| BAPL, points | 2.75 (0.55) | 2.91 (0.30) | 2.69 (0.60) | 2.86 (0.38) | 2.56 (0.73) | 0.750 | 0.220 | 0.402 |
| Global SUVR | 1.33(0.25) | 1.30 (0.25) | 1.38 (0.26) | 1.42 (0.30) | 1.35 (0.24) | 0.557 | 0.423 | 0.832 |

CF, center frequency; APOE, apolipoprotein E; CDR, Clinical Dementia Rating; MMSE, Mini Mental Status Exam; BAPL, brain amyloid plaque load on amyloid PET scans; SUVR, standardized uptake value ratio score, calculated by comparing the uptake of amyloid tracer in the brain cortex to that in the cerebellum
Note. Continuous variables are presented as mean (standard deviation) and categorical variables as number (percentage).
<sup>\*</sup> 12-week flickering light stimulation at the baseline center frequency of each participant
<sup>†</sup> *a* vs *c*, *b* vs *c*: Welch's t-test for continuous variables and Fisher's exact test for categorical variables, reflecting exploratory comparisons between included and excluded participants.
d vs e: Mann–Whitney U test for continuous variables and Fisher’s exact test for categorical variables.
<sup>‡</sup>One excluded participant lacked A $\beta$ -PET imaging data due to refusal to undergo mandatory imaging; SUVR and BAPL values are reported for the remaining 20 individuals.

Exploratory comparisons between enrolled and excluded participants revealed no significant differences in age, sex, education, APOE ε4 status, or baseline MMSE scores (all p > 0.05). The subset excluded specifically for insufficient gamma entrainment (n = 11) also exhibited comparable baseline characteristics, suggesting that entrainment capacity may reflect underlying neurophysiological factors rather than overt clinical features.

### Treatment Adherence and Safety

Participants completed a mean of 94.9 ± 13.4% of prescribed sessions (113.8 ± 16.0 of 120 sessions). No participants discontinued due to adverse effects. Across 1,920 total sessions, moderate-to-severe adverse effects (severity ≥ 3) were infrequent: asthenopia (1.8%), dazzling sensation (1.4%), fatigue (1.4%), dizziness (0.7%), ocular pain (0.6%), and headache (0.5%). When stratified by CF response (Table 2), groups showed distinct adverse effect patterns. The ICF-group reported higher frequencies of dizziness (12.59% vs 1.79%, p < 0.001) and dazzling sensation (13.98% vs 5.36%, p < 0.001), whereas the ICF+ group experienced more frequent fatigue (5.83% vs 1.02%, p < 0.001) and asthenopia (7.38% vs 2.87%, p < 0.001).

**Table 2.** Adverse effect profiles during the 12-week intervention stratified by center frequency response.

|  | All | Center frequency after intervention<sup>*</sup> |  |  |
| --- | --- | --- | --- | --- |
|  |  | Increased | Not increased | p<sup>†</sup> |
|  | (n = 16) | (n = 7) | (n = 9) |  |
| Scores, mean (SD)<sup>‡</sup> |  |  |  |  |
| Fatigue | 0.766 (0.827) | 0.541 (0.648) | 0.942 (0.943) | 0.536 |
| Headache | 0.254 (0.300) | 0.241 (0.298) | 0.265 (0.319) | 1.000 |
| Dizziness | 0.744 (1.095) | 0.432 (0.412) | 0.987 (1.403) | 0.758 |
| Dazzling | 1.014 (1.184) | 0.743 (0.697) | 1.226 (1.466) | 0.607 |
| Asthenopia | 0.706 (0.734) | 0.702 (0.909) | 0.709 (0.626) | 0.681 |
| Ocular pain | 0.240 (0.345) | 0.183 (0.376) | 0.287 (0.318) | 0.607 |
| Frequencies, n (%)<sup>§</sup> |  |  |  |  |
| Fatigue | 60 (3.125) | 49 (5.833) | 11 (1.019) | < 0.001 |
| Headache | 13 (0.677) | 11 (1.310) | 2 (0.185) | 0.004 |
| Dizziness | 151 (7.865) | 15 (1.786) | 136 (12.593) | < 0.001 |
| Dazzling | 196 (10.208) | 45 (5.357) | 151 (13.981) | < 0.001 |
| Asthenopia | 93 (4.844) | 62 (7.381) | 31 (2.870) | < 0.001 |
| Ocular pain | 18 (0.938) | 12 (1.429) | 6 (0.556) | 0.045 |

SD, standard deviation
<sup>\*</sup> 12-week flickering light stimulation at the baseline center frequency of each participant
<sup>†</sup> Mann-Whitney U test for continuous variables and Fisher's exact test for categorical variables
<sup>‡</sup> 7-point Likert scale (0 = not at all, 6 = extremely severe)
<sup>§</sup> The number of treatment sessions in which participants reported adverse effects as severe (scoring 3 or higher) and the proportion of these sessions relative to the total number of sessions (n = 1,920). The ICF+ and ICF- groups contributed 840 and 1,080 sessions, respectively, across 7 and 9 participants.

### CF Change and Cognitive Outcomes

Post-hoc stratification by CF response yielded two groups: CF-increased (ICF+, n = 7; 43.8%) and CF-not-increased (ICF-, n = 9; 56.3%) (Figure 2A). The groups showed no significant baseline differences in demographics, cognitive scores, APOE ε4 status, or amyloid burden (all p > 0.05; **Table 1**). Individual data are provided in **Supplementary Table 4.** Treatment adherence was comparable between groups (ICF+: 99.5 ± 1.2%; ICF-: 98.9 ± 7.4%; U = 34.00, p = 0.785).). Treatment adherence was uniformly high in both groups (ICF+: 99.5 ± 1.2%; ICF−: 98.9 ± 7.4%; U = 34.00, p = 0.785), ruling out compliance as a confounding factor.

**Figure 2.**
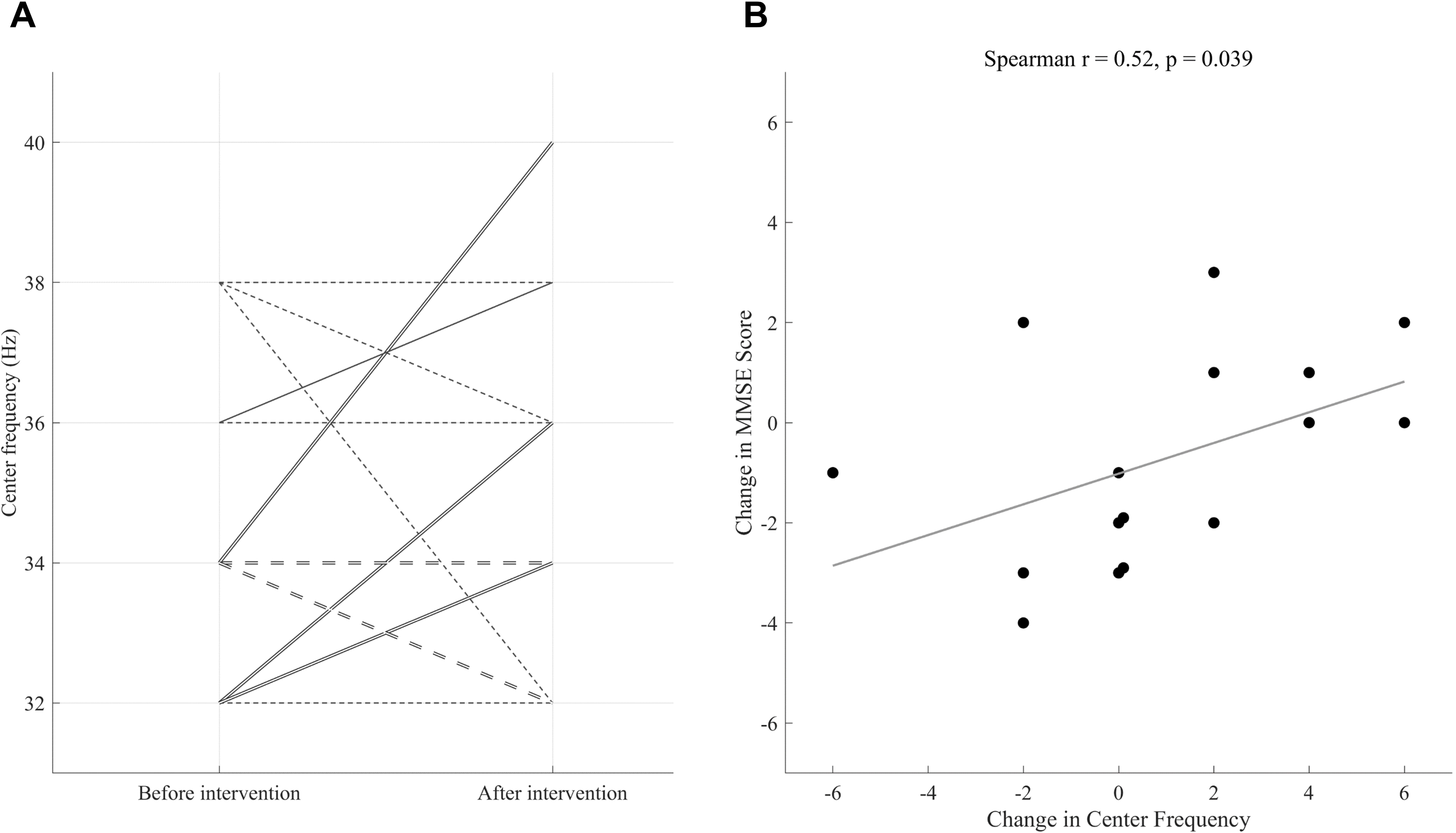
Center frequency restoration and cognitive preservation following personalized gamma entrainment. (A) Individual CF trajectories following 12-week personalized FLS-CF intervention. Solid lines represent participants who exhibited an increase in CF, while dashed lines indicate no change or a decrease. Line thickness reflects the number of participants represented (double line = two participants; single line = one participant). **(B)** Scatterplot illustrating the association between CF change and Mini Mental Status Exam score change (r = 0.52, p = 0.039). Each dot corresponds to an individual participant. MMSE, Mini Mental Status Exam *12-week flickering light stimulation at the baseline center frequency of each participant

Repeated-measures ANOVA revealed a significant time × group interaction for MMSE scores (F₁,₁₄ = 9.27, p = 0.009, ω² = 0.40; post-hoc power = 85.6%). The ICF+ group demonstrated cognitive maintenance (ΔMMSE: +0.71 ± 1.60; Cohen’s d = 0.45), while the ICF-group showed decline (ΔMMSE: -1.89 ± 1.76; Cohen’s d = -1.07), yielding a 2.6-point between-group difference over 12 weeks.

CF change correlated positively with MMSE change (r = 0.52, p = 0.039; Figure 2B). Logistic regression revealed that ICF+ participants were significantly more likely to maintain or improve cognition (OR = 48.00, 95% CI: 2.47-932.85, p = 0.010; wide CI reflects small sample). ROC analysis demonstrated good discriminative ability (AUC = 0.880, 95% CI: 0.631-1.000), with an optimal cutoff of +2 Hz yielding 86.5% sensitivity and 88.8% specificity. Neither adherence (r = 0.02, p = 0.94) nor baseline MMSE (r = 0.33, p = 0.21) correlated with CF change, suggesting that response variations reflect neurobiological rather than behavioral factors.

### Baseline Predictors of Treatment Response

Baseline EEG characteristics distinguished future responders (**Figure 3**). The ICF+ group demonstrated significantly higher resting-state gamma power, with the most pronounced differences in bilateral temporal regions (t = 2.16-2.75, Cohen’s d = 0.70-0.92, uncorrected p < 0.05). The ICF+ group also exhibited stronger gamma-band ERS during stimulation, particularly in right frontotemporal regions (t = 2.28-2.47, Cohen’s d = 1.12-1.23, uncorrected p < 0.05). Network-level analysis revealed the most robust differences. The ICF+ group showed significantly greater stimulus-induced connectivity increases (ΔsGC) from frontotemporal regions to multiple cortical areas (t = 2.19-2.92, Cohen’s d = 1.11-1.47, uncorrected p < 0.05), with enhanced interhemispheric communication originating from right occipital cortex. The ICF-group exhibited only weak, localized ΔsGC changes. Full statistical details are provided in **Supplementary Tables 5A-C**.

**Figure 3.**
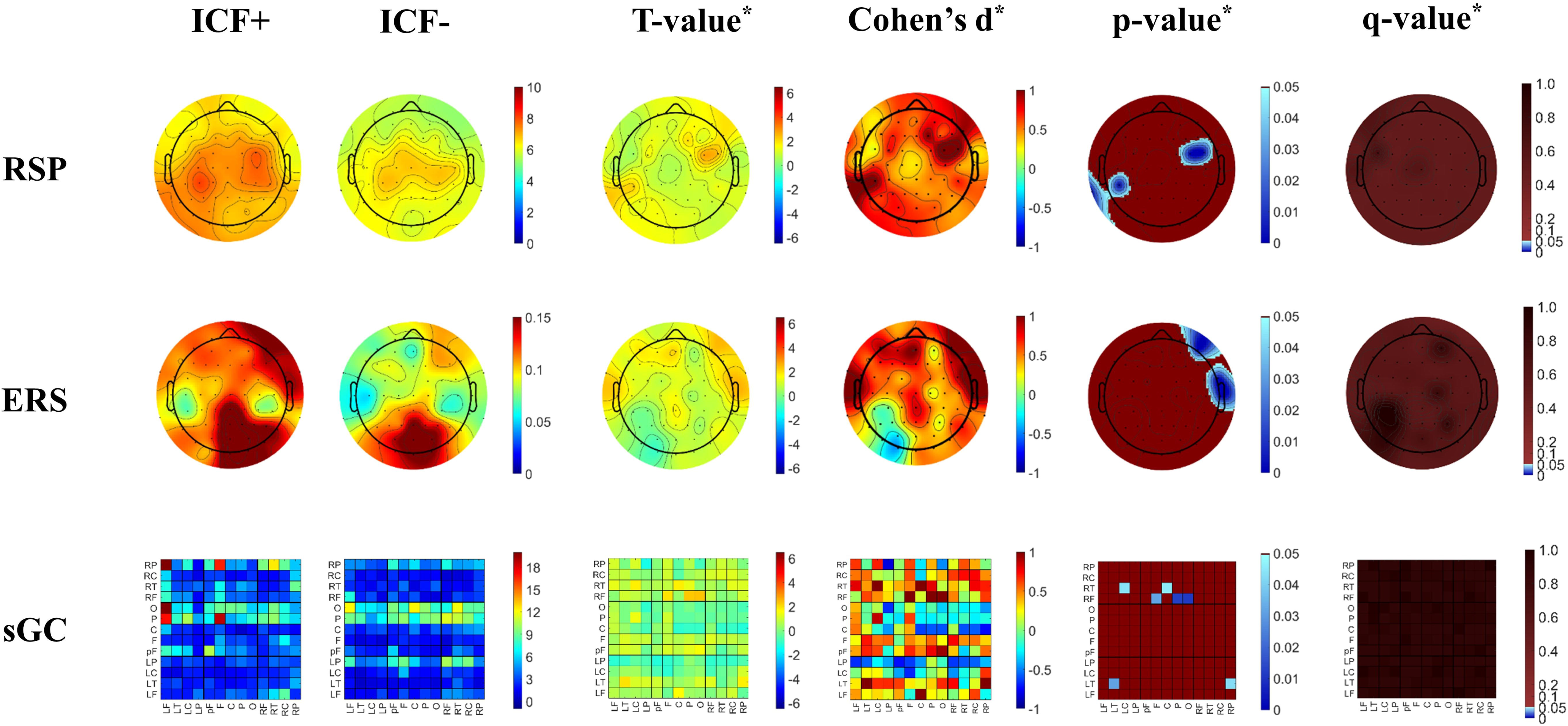
Baseline electrophysiological signatures distinguishing future treatment responders. ICF+, participants who showed an increase in center frequency after the intervention; ICF-, participants who did not show an increase in center frequency after the intervention; RSP, resting state power of gamma band; ERS, event-related synchronization of gamma band; sGC, spectral Granger causality of gamma band; PF, prefrontal; LF, left frontal; F, frontal; RF, right frontal; LT, left temporal; LC, left central; C, central; RC, right central; RT, right temporal; LP, left parietal; P, parietal; RP, right parietal; O, occipital. *Comparisons between the ICF+ and ICF-groups using Student’s t-test for independent samples. Both p- and q-value maps use blue scale to indicate regions with values < 0.05. The q-value maps represent FDR-corrected p-values (Benjamini–Hochberg), highlighting regions that remained significant after multiple comparisons correction.

LOOCV analysis identified specific features with predictive value (**Figure 4**; **Supplementary Tables 6A-C**). Among RSP features, channel TP9 achieved the highest accuracy (81.3%; AUC = 0.714; OR = 3.71, 95% CI: 1.02-13.54). For ERS, channel FT9 showed equivalent performance (OR = 4.45, 95% CI: 0.76-26.15). Among connectivity features, the F→RF connection demonstrated optimal performance (accuracy = 81.3%; AUC = 0.762; OR = 6.75, 95% CI: 0.66-69.48). Wide confidence intervals reflect the limited sample size. The concentration of predictive features in frontotemporal and occipital regions suggests these areas are critical for treatment responsiveness.

**Figure 4.**
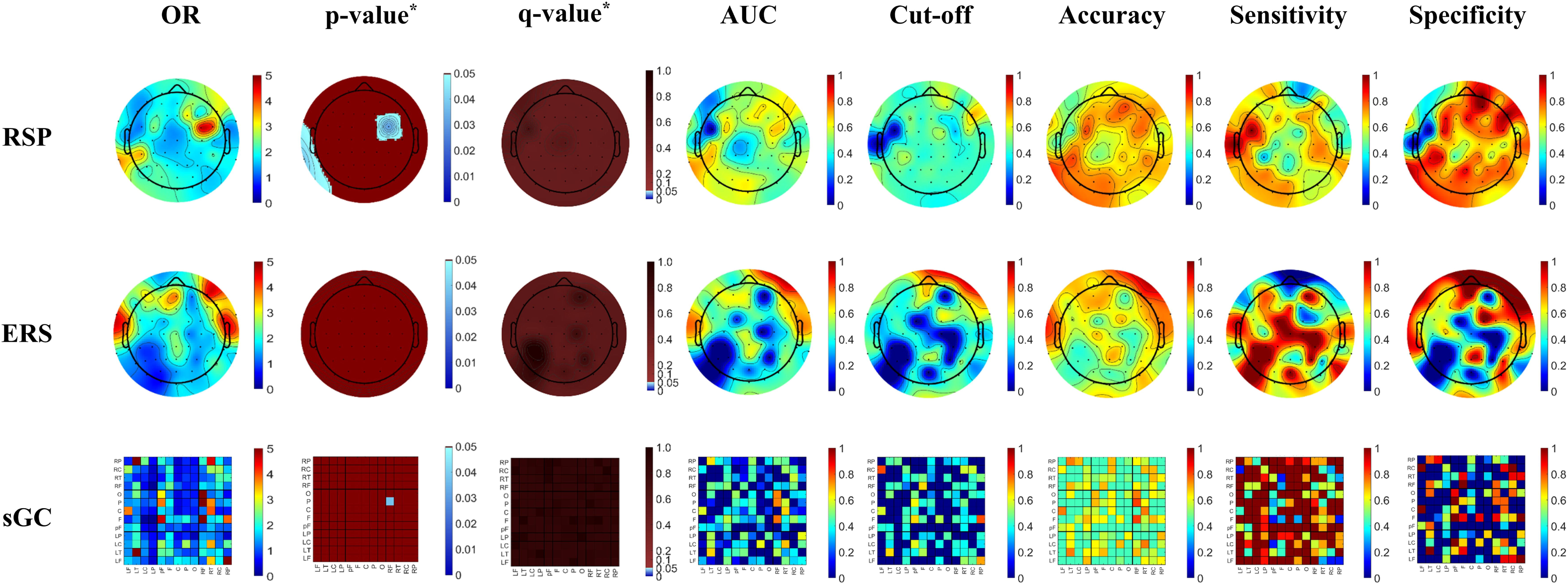
Predictive performance of baseline EEG features for treatment response: LOOCV analysis. RSP, resting state power of gamma band; ERS, event-related synchronization of gamma band; sGC, spectral Granger causality of gamma band, OR, odds ratio; CI, confidence interval; AUC, area under the curve; Cut-off, optimal threshold for classification; q value, false discovery rate–adjusted p value using the Benjamini–Hochberg procedure. PF, prefrontal; LF, left frontal; F, frontal; RF, right frontal; LT, left temporal; LC, left central; C, central; RC, right central; RT, right temporal; LP, left parietal; P, parietal; RP, right parietal; O, occipital. *Statistical significance of ORs was evaluated using uncorrected p-values and FDR-corrected q-values (Benjamini–Hochberg). The p- and q-value maps show regions with values < 0.05 in blue scale, indicating statistical significance before and after multiple comparisons correction.

### Post-Intervention Electrophysiological Changes

The intervention induced divergent electrophysiological changes between groups (**Figure 5**). Significant time × group interactions emerged for RSP in frontal and temporal regions (F₁,₁₄ = 5.34-12.10, ω² = 0.21-0.41, uncorrected p < 0.05; **Supplementary Tables 5A-C**). Post-hoc analyses revealed divergent trajectories (**Figure 6).** The ICF+ group demonstrated gamma power decreases in fronto-central areas (t = -5.55 to -2.58, d = -2.10 to -0.97, uncorrected p < 0.05) with simultaneous increases in parieto-occipital regions (t = 2.31-2.50, d = 0.77-0.83, uncorrected p < 0.05), suggesting anterior-posterior redistribution. The ICF-group showed the inverse pattern (increased frontal and decreased parietal gamma power). Both groups showed reduced ERS post-intervention (main effect of time: F₁,₁₄ = 4.78-49.17, ω² = 0.19-0.75, uncorrected p < 0.05), consistent with habituation. However, the ICF+ group retained greater responsiveness, with smaller ERS reductions in frontotemporal and midline regions (main effect of group: F₁,₁₄ = 4.89-10.25, ω² = 0.30-0.65, uncorrected p < 0.05).

**Figure 5.**
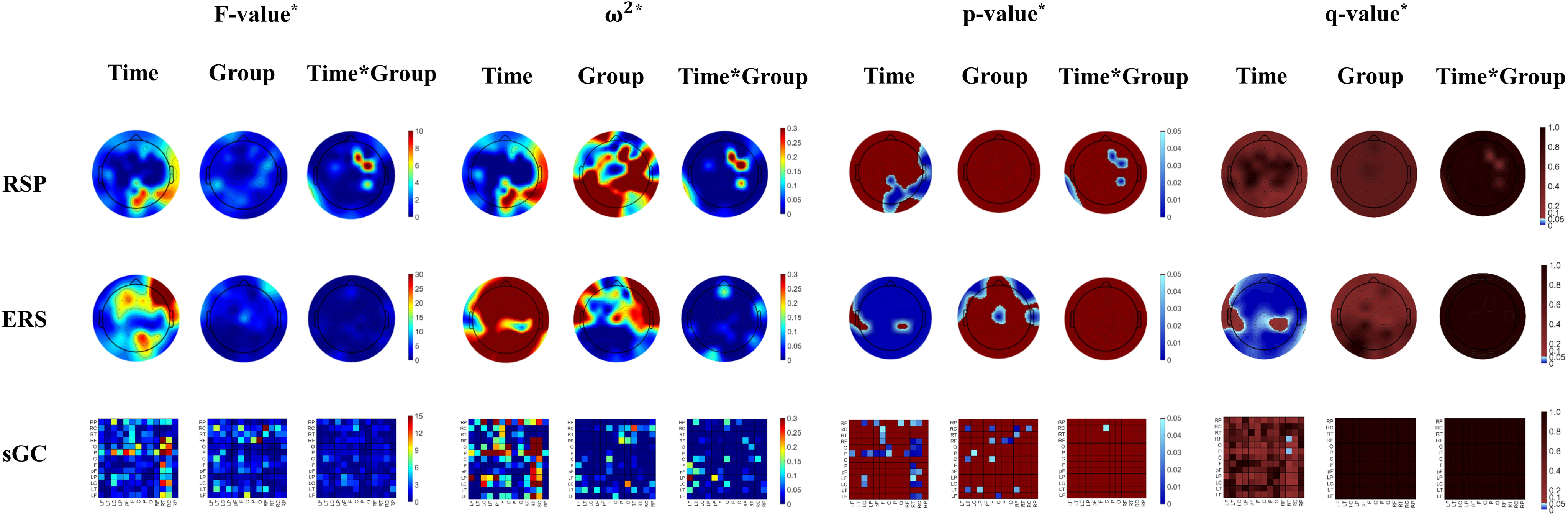
Post-intervention electrophysiological changes: Time × Group interaction effects. RSP, resting state power of gamma band; ERS, event-related synchronization of gamma band; sGC, spectral Granger causality of gamma band; PF, prefrontal; LF, left frontal; F, frontal; RF, right frontal; LT, left temporal; LC, left central; C, central; RC, right central; RT, right temporal; LP, left parietal; P, parietal; RP, right parietal; O, occipital *Repeated-measures analysis of variance examining the main effects of time (pre-intervention versus post-intervention) and group (participants who showed an increase in center frequency after the intervention versus those who did not show an increase in center frequency after intervention) and their interaction. Both p- and q-value maps use blue scale to indicate regions with values < 0.05. The q-value maps represent FDR-corrected p-values (Benjamini–Hochberg), highlighting regions that remained significant after multiple comparisons correction.

**Figure 6.**
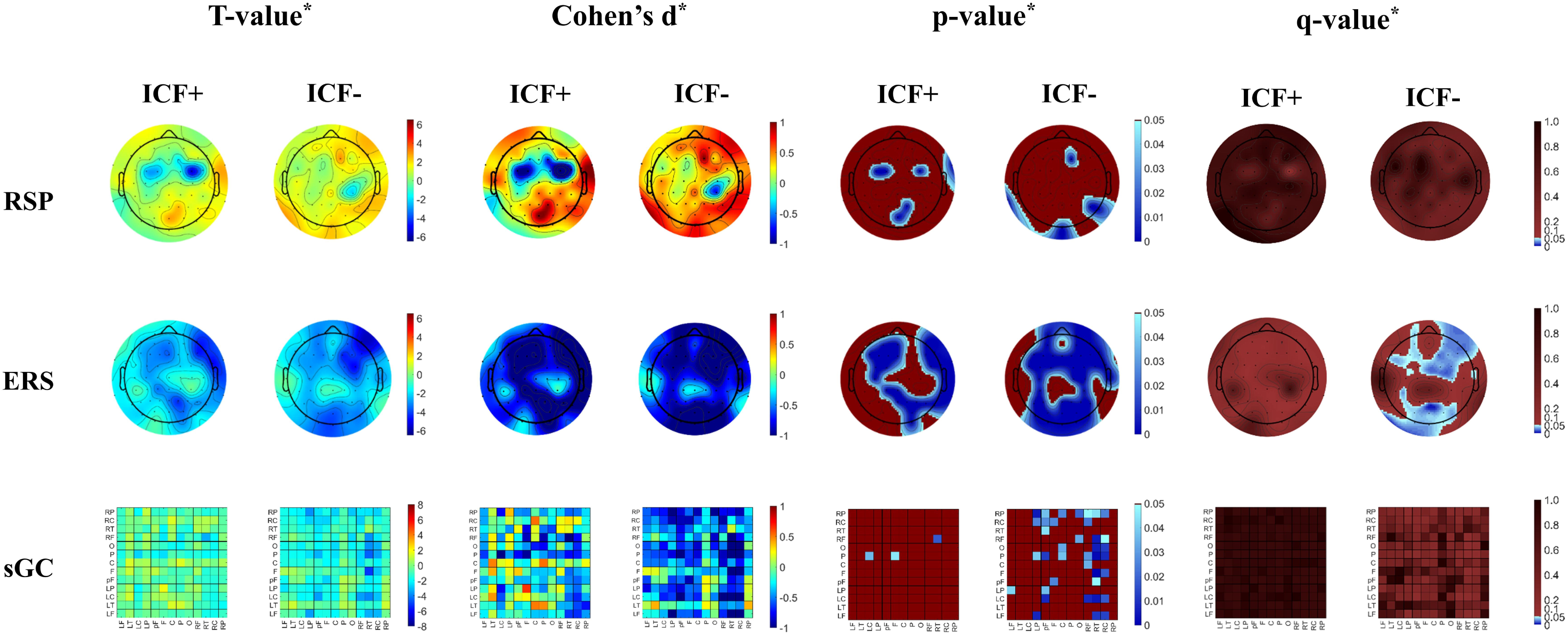
Divergent electrophysiological trajectories in responders versus non-responders. ICF+, participants who showed an increase in center frequency after the intervention; ICF-, participants who did not show an increase in center frequency after the intervention; RSP, resting state power of gamma band; ERS, event-related synchronization of gamma band; sGC, spectral Granger causality of gamma band; PF, prefrontal; LF, left frontal; F, frontal; RF, right frontal; LT, left temporal; LC, left central; C, central; RC, right central; RT, right temporal; LP, left parietal; P, parietal; RP, right parietal; O, occipital. *Paired t test examining within-group changes. Both p- and q-value maps use blue scale to indicate regions with values < 0.05. The q-value maps represent FDR-corrected p-values (Benjamini–Hochberg), highlighting regions that remained significant after multiple comparisons correction.

Functional connectivity showed the most profound group differences. The ICF+ group maintained and strengthened ΔsGC across widespread regions, whereas the ICF-group demonstrated widespread reductions, particularly in right hemisphere-to-frontal connections (t = -5.18 to -1.32, d = -1.73 to -0.78, uncorrected p < 0.05). These divergent patterns indicate that the ICF+ group underwent beneficial network reorganization while the ICF-group experienced progressive deterioration of directional information flow.

## Discussion

This pilot study provides the first mechanistic evidence that restoration of gamma CF serves as a promising biomarker for cognitive preservation in early AD. Our findings reveal that response to personalized gamma entrainment is not uniform but stratified by baseline neurophysiological characteristics. Participants demonstrating CF increase (ICF+) maintained cognitive function (MMSE change: +0.71 ± 1.60), whereas those without CF increase (ICF−) experienced significant decline (−1.89 ± 1.76), yielding a clinically meaningful 2.6-point divergence over 12 weeks. This dichotomous response pattern, coupled with the strong correlation between CF change and cognitive outcomes (r = 0.52, p = 0.039), suggests that CF restoration identifies a distinct biological subtype capable of adaptive plasticity.

### Neurobiological Significance of CF Modulation

Our findings suggest that CF restoration reflects genuine circuit-level recovery rather than mere frequency entrainment. In AD, progressive CF decline correlates with parvalbumin-positive (PV+) interneuron dysfunction.^30, 31^ Our demonstration that personalized gamma entrainment can reverse this CF decline suggests partial restoration of interneuron function. The observed CF increase in responders likely signals restoration of inhibitory-excitatory balance, consistent with findings in motor learning paradigms where CF upshifts mark functional gains.^20^ This mechanistic interpretation supports our hypothesis that CF modulation capacity serves as a proxy for neural reserve - the remaining functional integrity of inhibitory circuits capable of metabolic recovery under rhythmic stimulation.

The anterior-posterior redistribution of gamma power observed exclusively in the ICF+ group (decreased frontal activity coupled with increased parieto-occipital power) suggests adaptive circuit reorganization rather than passive frequency following. This interpretation aligns with recent evidence that 40Hz stimulation promotes glymphatic clearance^4^ and the “dynamic disequilibrium” model proposed by Harris et al., wherein external stimulation can rebalance disrupted circuits.^35^ We hypothesize a potential multi-stage therapeutic process: (1) baseline hyperactivity representing compensatory neural reserve, (2) activity normalization through enhanced clearance mechanisms, and (3) CF increase reflecting genuine circuit restoration. However, our two-timepoint design of the current study precludes definitive establishment of this temporal sequence; longitudinal studies with intermediate assessments are needed to confirm these putative therapeutic milestones.

### Baseline Electrophysiological Signatures as Predictive Biomarkers

A key translational finding is that baseline electrophysiological profiles reliably distinguished future responders from non-responders. The ICF+ group exhibited elevated baseline gamma power and enhanced functional connectivity, which we interpret as compensatory neural reserve. ^33, 34^ While often viewed as pathological, this hyperactivity paradoxically indicates that the underlying circuitry retains the metabolic capital to respond to stimulation. Critically, this “hyperactive” baseline signature appears to represent intact neural substrate capable of normalization, rather than irreversible pathological change.

Network-level analyses revealed the most robust predictive markers. The ICF+ group demonstrated significantly greater stimulus-induced connectivity changes (ΔsGC; Cohen’s d = 1.11–1.47), particularly in frontotemporal-to-cortical pathways. The LOOCV analysis identified specific topographical features with clinical utility: TP9 (left temporoparietal) resting-state gamma power achieved 81.3% classification accuracy, while the F→RF connectivity metric showed optimal predictive performance (AUC = 0.762). These findings suggest that preserved network-level responsiveness (the capacity for dynamic reorganization) may be more prognostically informative than static power measures alone.

This framework may address why the OVERTURE trial, despite failing to meet primary endpoints in the overall population, showed benefits in certain subgroups.^7^ Our results demonstrate that electrophysiological profiling could prospectively identify these responsive subgroups, potentially transforming our understanding of the heterogeneous outcomes in gamma entrainment trials.^5, 8^ In contrast, the “hypoactive” baseline profile observed in non-responders likely reflects a system that has crossed the threshold of recoverable function, where insufficient neural reserve precludes adaptive reorganization.

### The “Non-Entrainer” Phenotype and Trial Design Implications

A critical finding with immediate translational relevance is that 29.7% of screened individuals (11/37) failed to demonstrate baseline gamma entrainment. These “non-entrainers” likely represent a distinct biological subtype with advanced circuit degeneration, possibly involving severe PV+ interneuron loss or white matter dysconnectivity, that precludes the propagation of entrained rhythms. This finding directly parallels Soula et al.’s observation that only 25% of visual cortex neurons showed entrainment in AD model mice,^9^ suggesting that entrainment capacity varies substantially even within disease populations. The identification and prospective exclusion of this “non-entrainer” subtype is essential for future trial design to avoid diluting efficacy signals - a factor that likely contributed to the heterogeneous results in previous fixed-frequency trials.^5, 7, 8^

### Differential Functional Network Trajectories

The functional connectivity analyses revealed fundamentally divergent trajectories between groups. Post-intervention, the ICF+ group maintained and strengthened directional connectivity (ΔsGC) across widespread cortical regions, indicating preserved capacity for adaptive network reorganization. In contrast, the ICF− group demonstrated widespread reductions in ΔsGC, particularly in right hemisphere-to-frontal pathways (Cohen’s d = −0.78 to −1.73), consistent with progressive deterioration of directional information flow. This pattern aligns with Corriveau-Lecavalier et al.’s description of maladaptive hyperactivity that perpetuates excitotoxicity.^37^ These findings underscore that successful gamma entrainment requires not merely the ability to follow external rhythms, but sufficient neural reserve to undergo beneficial plasticity.

### Bridging the Translational Gap

Our results bridge the translational gap between the dramatic preclinical success of gamma entrainment^1, 3, 32^ and modest human trial outcomes.^5, 7, 8^ Our individualized approach determining optimal stimulation frequency for each participant based on baseline CF represents a fundamental departure from fixed 40Hz protocols. The convergence between our 29.7% entrainment failure rate and the limited neuronal entrainment observed in animal models^9^ underscores that patient stratification and personalized frequency selection are prerequisites for therapeutic success. The heterogeneity in human gamma responses likely reflects complex interactions between genetic factors, disease stage, and individual circuit integrity. While our study found no baseline differences in APOE ε4 carrier status between groups, the small sample size precludes definitive conclusions about genetic modifiers, warranting adequately powered studies to characterize these interactions.^38^

### Differential Adverse Effect Profiles as Mechanistic Clues

The divergent adverse effect profiles between groups (Table 2) offer unexpected mechanistic insights and potential clinical utility. The ICF+ group’s predominant fatigue (5.8% vs 1.0%) and asthenopia (7.4% vs 2.9%) may reflect active cortical engagement - the ‘work’ of successful neural entrainment and reorganization. Conversely, the ICF-group’s higher incidence of dizziness (12.6% vs 1.8%) and dazzling sensations (14.0% vs 5.4%) suggests sensory-integration failure and network instability when stimulation cannot be properly integrated. These symptom profiles might serve as early, clinically accessible indicators of treatment responsiveness.

### Clinical Translation and Precision Medicine Implications

The identification of CF change as a treatment response biomarker has immediate translational implications. The optimal CF change cutoff of +2 Hz (86.5% sensitivity, 88.8% specificity; AUC = 0.880) provides a clinically actionable metric for treatment monitoring. Practical applications include: (1) early identification of non-responders within weeks of treatment initiation; (2) real-time optimization of stimulation parameters based on CF trajectory; and (3) patient stratification for combination therapy approaches. This finding is particularly timely given the recent approvals of anti-amyloid antibodies such as lecanemab^39^ and donanemab^40^ with heterogeneous response rates, suggesting potential for combination therapy guided by electrophysiological biomarkers. The non-invasive nature of gamma entrainment, combined with biomarker-guided personalization, positions it as an ideal adjunctive therapy to pharmacological interventions. These findings advance the precision medicine framework for AD, demonstrating that electrophysiological phenotyping could transform both clinical trial design and therapeutic practice.^41^

### Limitations and Future Directions

Several limitations warrant consideration. First, the modest sample size (n = 16) and open-label design preclude definitive efficacy conclusions; however, as a proof-of-concept study, our primary aim was mechanistic insight and biomarker identification rather than clinical efficacy demonstration. The robust correlation between objective electrophysiological changes and cognitive outcomes provides compelling preliminary evidence for the proposed framework. Second, the 12-week duration cannot address long-term sustainability or disease modification. Third, the absence of a sham control precludes definitive exclusion of placebo effects, although the correlation between objective EEG measures and cognitive outcomes argues against purely expectation-driven responses. Fourth, the post-hoc stratification based on CF change, while necessary for hypothesis generation, cannot establish causality, as unmeasured confounders could explain group differences despite comparable measured characteristics. Prospective validation with a priori CF-based stratification is essential.

These findings establish clear research priorities:. (1) phase II randomized controlled trials with CF change as a primary endpoint and sham-controlled design; (2) development of point-of-care EEG systems for clinical implementation; (3) investigation of combination approaches with anti-amyloid therapies; and (4) characterization of non-entrainers to develop enhancement strategies such as pharmacological priming or alternative stimulation modalities. The 29.7% entrainment failure rate represents a critically understudied population. This failure likely reflects severe circuit disruption (extensive white matter pathology, profound PV+ interneuron loss, or genetic factors) that precludes gamma propagation. Understanding whether pharmacological priming or alternative modalities can restore entrainment capacity in these individuals represents an important direction for expanding therapeutic reach.

## Conclusions

This pilot study establishes CF restoration as a promising biomarker for identifying responders to personalized gamma entrainment in early AD. By demonstrating that individualized frequency selection and baseline electrophysiological profiling can predict treatment response, we provide a roadmap for precision neuromodulation in neurodegenerative disease. The divergent electrophysiological and cognitive trajectories between responders and non-responders reveal fundamental differences in neural reserve and plasticity capacity. While validation in larger, sham-controlled trials is essential, these findings suggest that biomarker-guided patient selection could substantially improve clinical trial success rates and therapeutic outcomes. Personalized gamma entrainment, guided by electrophysiological biomarkers, represents a safe, non-invasive intervention with potential for integration into multimodal therapeutic paradigms for AD.

## Data Access Statement

Data are not publicly available, but access may be considered upon reasonable request and subject to institutional approval.

## Data Availability Statement

The dataset is not available for public sharing due to ethical and regulatory restrictions.

## Supplementary material

Supplementary material is available at online.

