## Supplementary for "Restoration of Gamma Center Frequency via Personalized Entrainment Marks Cognitive Preservation in Early Alzheimer’s Disease"

Manuscript title:

**Supplementary Appendix 1. OLED-based FLS Device System**

***Device Design and Configuration***

The FLS device was designed as eyewear containing organic light-emitting diode (OLED) panels positioned 2 cm from each pupil. The glasses-like frame was fabricated using a 3D printer (SLA-type with ABS-like resin; HY3D Co., Ltd., Ansan, Korea). Each eye received light from two OLED panels (A88MA2B; Konica Minolta Inc., Tokyo, Japan) with individual active areas of 43.4 mm × 15.9 mm, arranged along the short axis to create a combined active area of 43.4 mm × 31.8 mm per eye. Due to bus electrode constraints, a 7 mm gap existed between the panels. However, geometric simulation using LightTools™ software confirmed uniform light distribution across the pupil despite this gap (Supplementary Fig. 1).

***Optical Characteristics***

The OLED panels were characterized using a spectroradiometer (CS2000, Konica Minolta Inc.) and source measure unit (Keithley 2400, Tektronix Inc.). To achieve the target illuminance of 1385 lm/m² at a 2 mm pupil diameter and 2 cm viewing distance, we set the OLED luminance to 1120 cd/m². The emission spectrum ranged from 400-700 nm with a primary peak at 446 nm and secondary peaks at 437 nm and 562 nm, resulting in a color temperature of 5700 K similar to daylight (Supplementary Fig. 2).

***Electronic Control System***

We developed a custom AC circuit powered by a standard 12V/2A household adapter. The circuit supported five flickering frequencies (32, 34, 36, 38, and 40 Hz) operating at 50% duty cycle. Low RC delay ensured precise on/off transitions, which were verified by oscilloscope and power meter measurements (Supplementary Fig. 3A-B). The circuit included a current control dial that enabled linear luminance adjustment, with approximately 180 mA required to achieve 1120 cd/m². All 20 devices used in the clinical trial operated within ±2.5% luminance variation (Supplementary Fig. 3C-D).

***Thermal Management and Safety Features***

Thermal imaging (U5856A; Keysight Technologies Inc.) showed that panel temperature stabilized at 37°C after 20 minutes of continuous operation, remaining stable for extended periods up to 90 minutes (Supplementary Fig. 4). To ensure safety and reliability, the device incorporated several protective features. A battery backup system prevented unintended shutdowns during use, while an automatic power-off relay timer ensured complete shutdown after the designated period. The entire system was housed in a UL94 HB-rated ABS plastic enclosure (AC 3220; Caseforyou Inc., Daejeon, Korea) to meet fire safety standards (Supplementary Fig. 5).

***Data Logging and User Interface***

The device incorporated Bluetooth Low Energy (BLE) communication for wireless control and data logging. A custom mobile application managed device operation, recorded usage data, and collected post-session surveys (Supplementary Fig. 6). The application featured a simplified one-way interface designed specifically for ease of use by elderly participants. All control signals and operation logs were automatically recorded according to protocols detailed in Supplementary Tables 1-3, ensuring comprehensive documentation of device usage throughout the clinical trial.

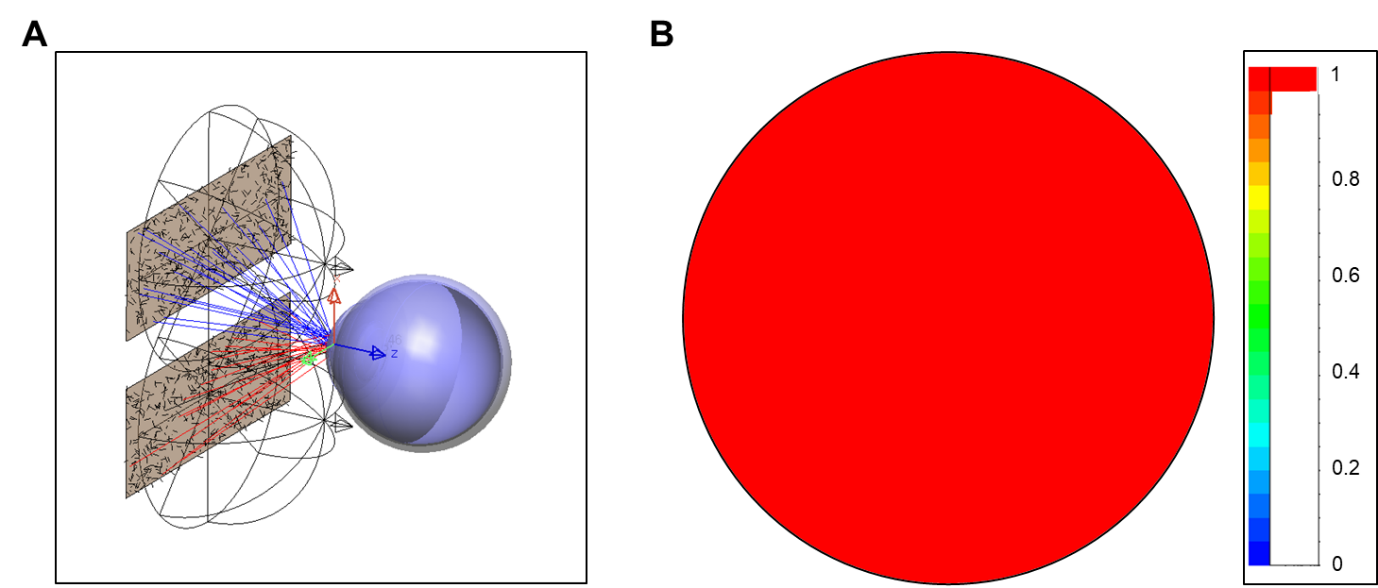

**Supplementary Figure S1**. Light distribution delivered to the pupil by two panels based on geometric optics simulation. (A) A schematic diagram of the eye model (LightTools^TM^) and OLED panels separated by 2 cm. (B) A result of the illuminance distribution at the pupil

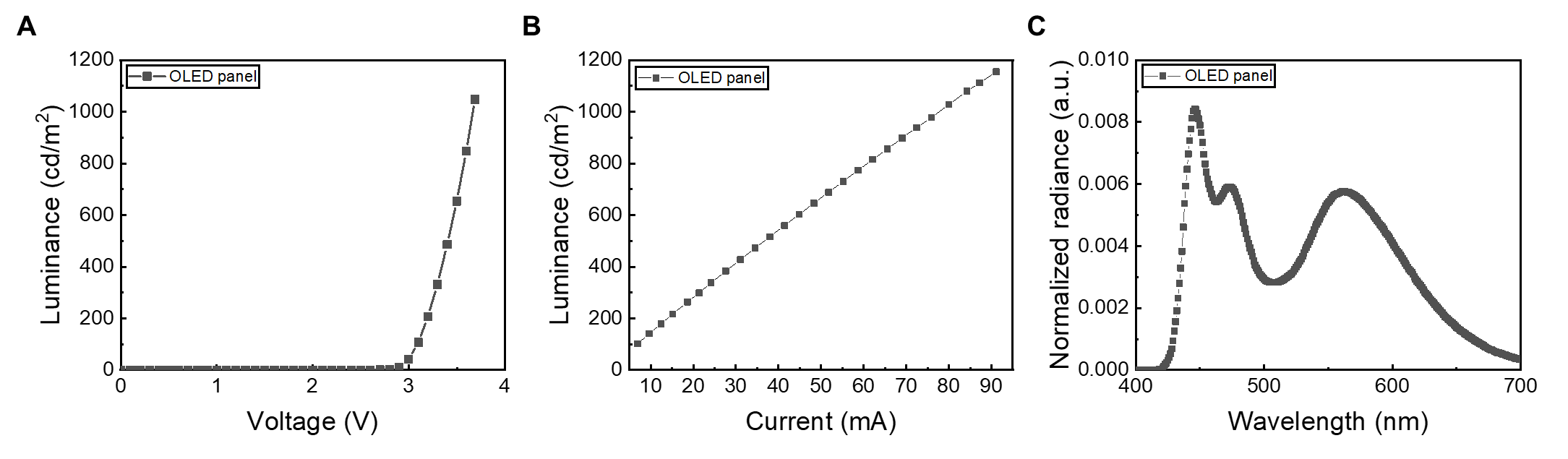

**Supplementary Figure S2**. Electrical and optical characteristics of a single OLED panel: Relationships between luminance and (A) Voltage, (B) Current. (C) Normalized spectrum of white OLED panels with 5700K.

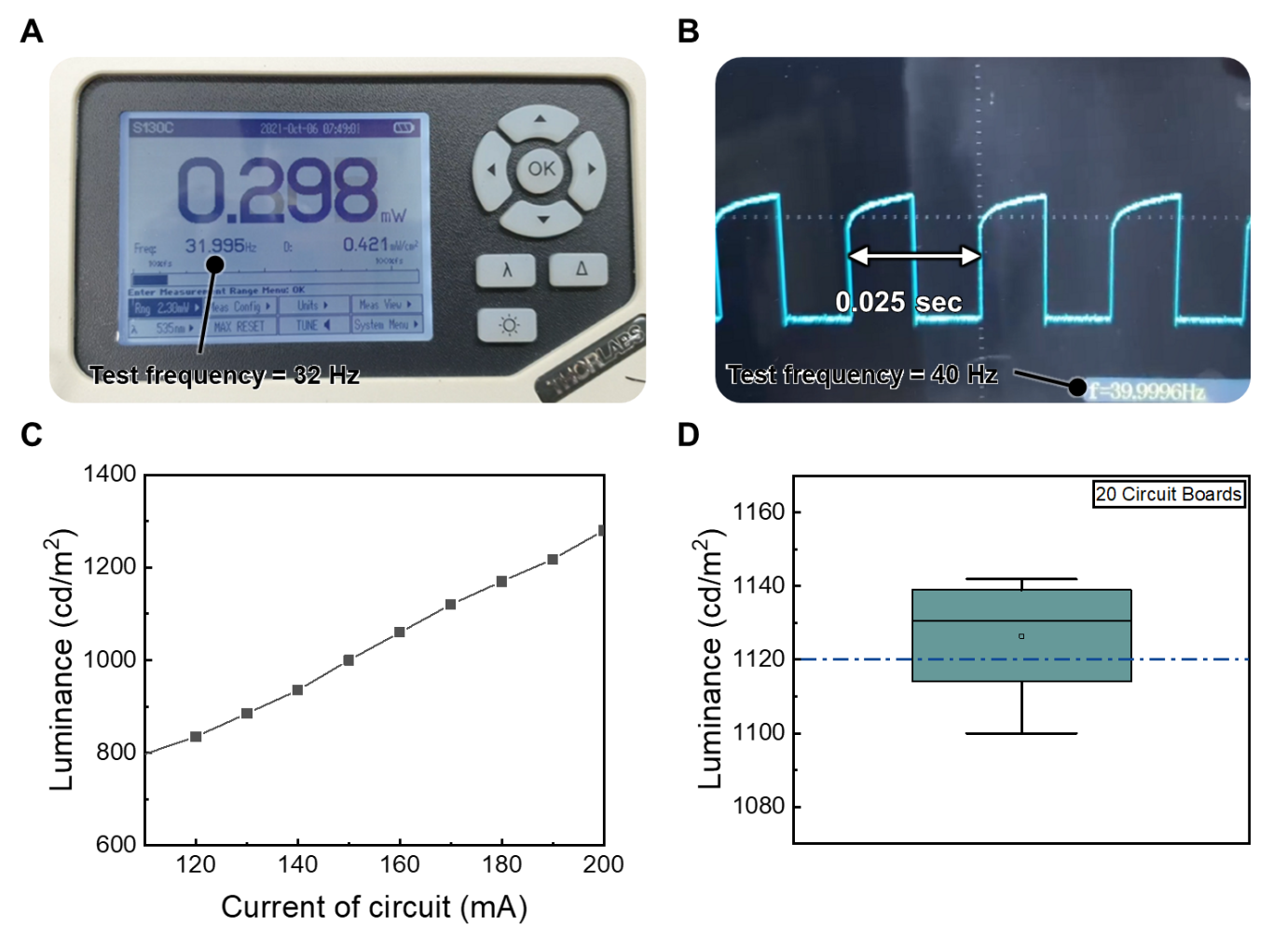

**Supplementary Figure S3**. Characteristics of two OLED panels driven by an in-house developed circuit: (A) Testing AC operation of the circuit board using a power meter (e.g., 32 Hz) and (B) an oscilloscope (e.g., 40 Hz). (C) Final luminance of the OLED glasses according to changes in the circuit's current dial. (D) Final luminance (target: 1120 cd/m²) variation across clinical trial devices (20 units).

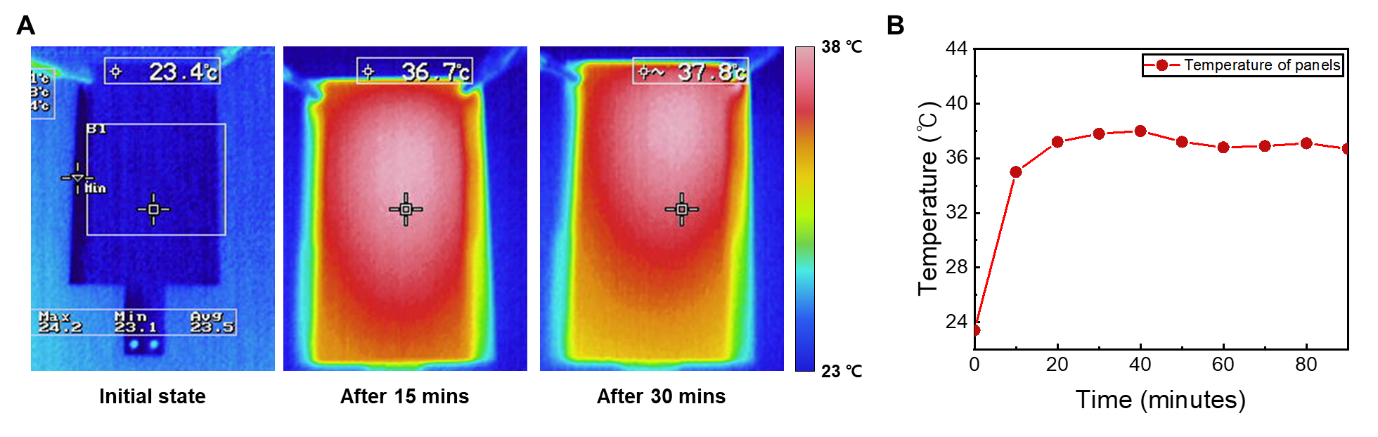

**Supplementary Figure S4**. Temperature changes of the OLED panel during continuous operation at 1120 cd/m²: (A) Thermal camera images over driving time. (B) Temperature variation graph during continuous operation for 90 minutes (3 times of actual experiment duration).

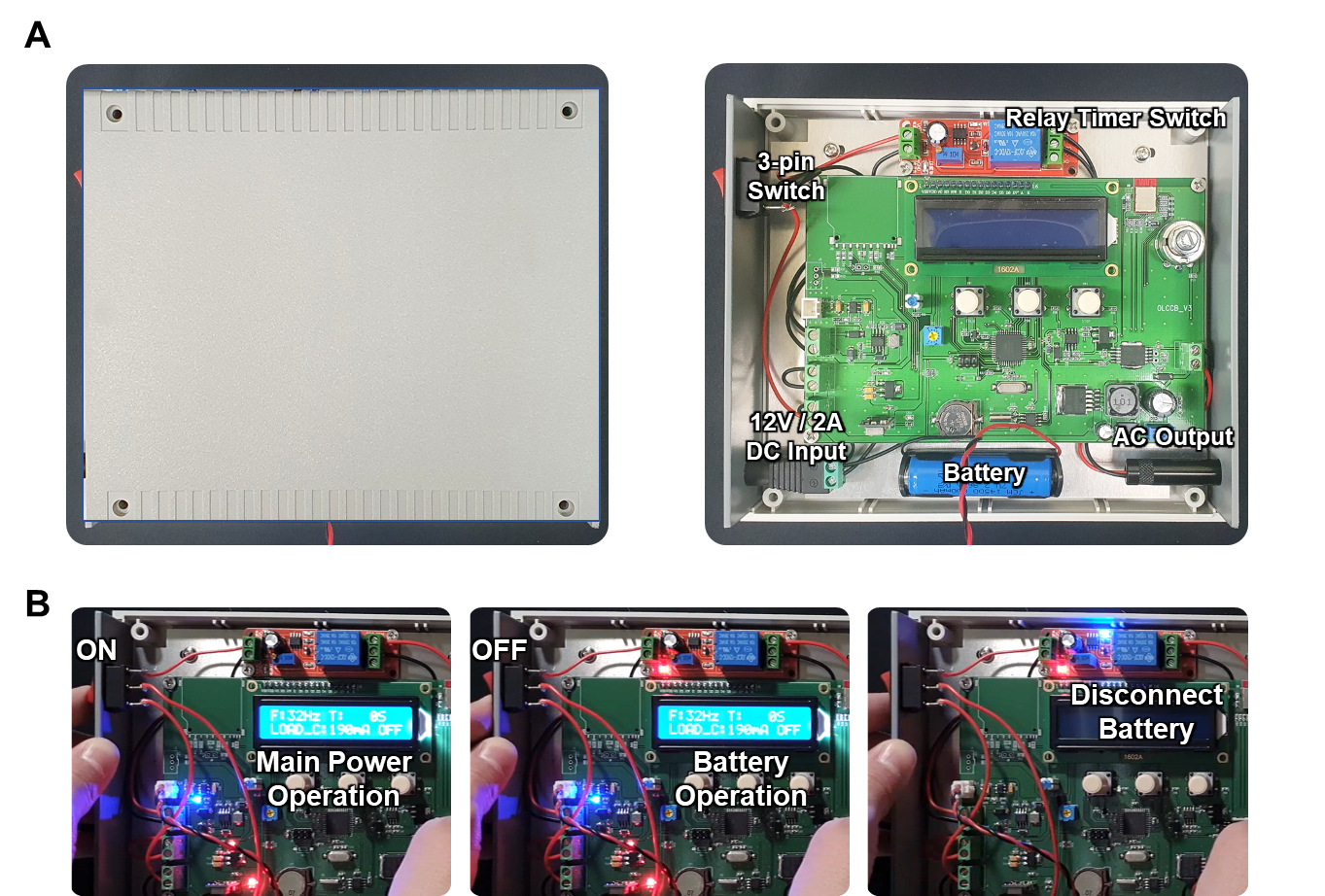

**Supplementary Figure S5**. Photographs of the in-house developed circuit system: (A) Main circuit board and components inside the enclosure (battery, relay timer switch, etc.). (B) Example of operation using the timer switch: main power, battery power, and turn off sequence.

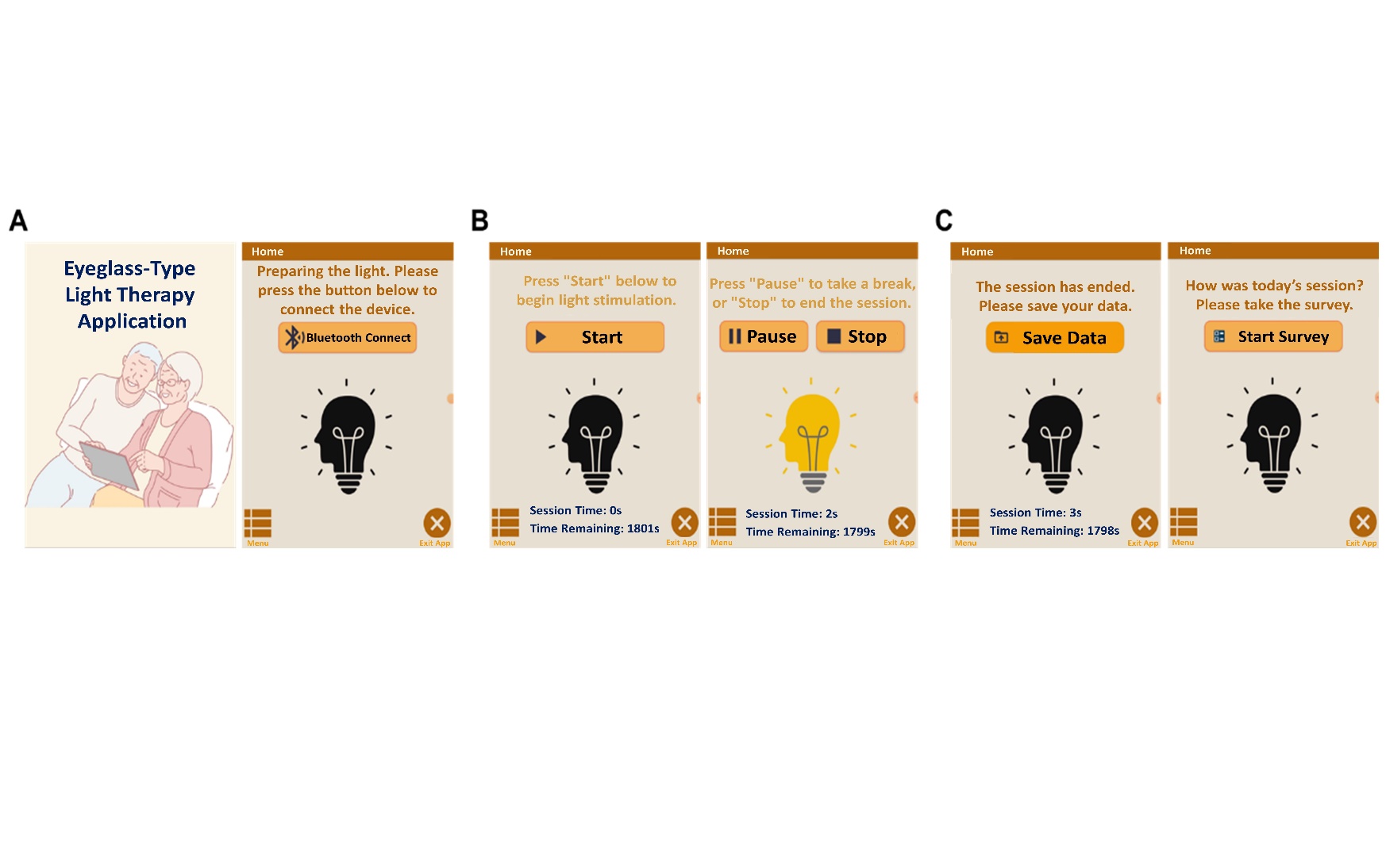
**Supplementary Figure S6**. Example screens of the in-house developed Android app for controlling the circuit system: **(A)** Communication with the main circuit board, **(B)** Communication with the patient, **(C)** Communication with the hospital.

Supplementary Table 1. Action signals transmitted from the app to the board.

| **STX ^a)^** | **FUNC ^b)^** | **CMD ^c)^** | | **CHECKSUM ^d)^** | **ETX ^e)^** |
| --- | --- | --- | --- | --- | --- |
| 0xf1 | 0x81 | BLE Connection | 0x10 | (STX^FUNC^CMD)+1 | 0x02 |
|  |  | START | 0x20 |  |  |
|  |  | PAUSE | 0x30 |  |  |
|  |  | STOP | 0x40 |  |  |

a) Start byte. b) Data classification byte sent from the app to the board. c) Action signal. d) Data confirmation signal, where ^ represents "and" logical operation. e) End byte.

Supplementary Table 2. Data transmitted from the board to the app when START is transmitted from the app to the board.

| **STX** | **FUNC** | **CURR ^a)^** | **FREQ ^b)^** | **STAT ^c)^** | | **CHECKSUM** | **ETX** |
| --- | --- | --- | --- | --- | --- | --- | --- |
| 0xfe | 0x82 | 0xXX | 0xXX | START | 0x10 | (STX^FUNC^CURR^FREQ^STAT)+1 | 0x02 |
|  |  |  |  | PAUSE | 0x20 |  |  |
|  |  |  |  | STOP | 0x30 |  |  |
|  |  |  |  | Battery RUN | 0x40 |  |  |
|  |  |  |  | No Load | 0x50 |  |  |
|  |  |  |  | No Current | 0x60 |  |  |

a) Hex value of the operating current. b) Hex value of the operating frequency. c) Operating states of the circuit where normal operations correspond to START, PAUSE, and STOP, while Battery RUN indicates the operation of a Li-ion battery.

Supplementary Table 3. Data transmitted from the board to the app when one trial ends or when PAUSE/STOP is transmitted from the app to the board.

| **STX** | **FUNC** | **Yr. ^a)^** | **Mo. ^a)^** | **Day ^a)^** | **Hr. ^a)^** | **Min. ^a)^** | **Sec. ^a)^** | **Op. Time ^b)^** | | **Total**  **Time ^b)^** | | **STAT** | **CHECK**  **SUM** | **ETX** |
| --- | --- | --- | --- | --- | --- | --- | --- | --- | --- | --- | --- | --- | --- | --- |
| 0xfe | 0x83 | 0xXX (Date) | | | | | | H | L | H | L | 0x(10 ~ 60) | AND | 0x02 |

a) Transmitting values for each byte corresponding to the date, utilizing only the last two digits for YEAR. b) As the operation time exceeds the size of one byte with a maximum of 1800 seconds, it is divided into two bytes, High (H) and Low (L), for transmission.

Supplementary Table 4. Individual participant-level outcomes before and after the intervention.

| **ID** | **Age range** | **Sex** | **Education**  **(years)** | **Adherence (%)** | **Group** | **CF (Baseline)** | **CF**  **(Post)** | **ΔCF** | **CDR (Baseline)** | **CDR**  **(Post)** | **ΔCDR** | **CDR SOB (Baseline)** | **CDR SOB (Post)** | **ΔCDR SOB** | **MMSE (Baseline)** | **MMSE (Post)** | **ΔMMSE** | **APOE genotype** | **SUVR (Baseline)** | **SUVR (Post)** | **ΔSUVR** | **BAPL (Baseline)** | **BAPL (Post)** |
| --- | --- | --- | --- | --- | --- | --- | --- | --- | --- | --- | --- | --- | --- | --- | --- | --- | --- | --- | --- | --- | --- | --- | --- |
| 01 | 71-75 | F | 12 | 100 | + | 32 | 36 | 4 | 0.5 | 0.5 | 0 | 2.5 | 2.5 | 0 | 20 | 21 | 1 | ε3/ε4 | 1.5 | 1.46 | -0.04 | 3 | 3 |
| 02 | 71-75 | M | 16 | 100 | + | 34 | 40 | 6 | 0.5 | 0.5 | 0 | 2.5 | 2 | -0.5 | 23 | 25 | 2 | ε3/ε3 | 1.55 | 1.55 | 0 | 3 | 3 |
| 03 | 71-75 | M | 14 | 100 | − | 34 | 34 | 0 | 0.5 | 0.5 | 0 | 1.5 | 2.5 | 1 | 23 | 21 | -2 | ε3/ε3 | 1.31 | 1.33 | 0.02 | 1 | 1 |
| 04 | 71-75 | F | 12 | 96.7 | + | 32 | 34 | 2 | 0.5 | 0.5 | 0 | 3.5 | 2.5 | -1 | 19 | 20 | 1 | ε3/ε4 | 1.11 | 1.13 | 0.02 | 3 | 3 |
| 05 | 71-75 | F | 6 | 80 | − | 32 | 32 | 0 | 0.5 | 0.5 | 0 | 3 | 2.5 | -0.5 | 17 | 14 | -3 | ε3/ε3 | 1.19 | 1.19 | 0 | 3 | 3 |
| 06 | 66-70 | M | 16 | 75 | − | 38 | 38 | 0 | 0.5 | 0.5 | 0 | 0.5 | 0.5 | 0 | 27 | 24 | -3 | ε3/ε3 | 1.13 | 1.15 | 0.02 | 2 | 2 |
| 07 | 76-80 | F | 16 | 100 | − | 38 | 36 | -2 | 0.5 | 1 | 0.5 | 4 | 5 | 1 | 19 | 15 | -4 | ε3/ε4 | 1.57 | 1.47 | -0.1 | 3 | 3 |
| 08 | 76-80 | F | 12 | 100 | − | 38 | 32 | -6 | 0.5 | 0.5 | 0 | 0.5 | 0.5 | 0 | 28 | 27 | -1 | ε3/ε3 | 1.2 | 1.18 | -0.02 | 3 | 3 |
| 09 | 81-85 | F | 3 | 96.7 | + | 32 | 36 | 4 | 0.5 | 0.5 | 0 | 3.5 | 4.5 | 1 | 15 | 15 | 0 | ε4/ε4 | 0.91 | 0.95 | 0.04 | 2 | 2 |
| 10 | 71-75 | M | 16 | 100 | − | 34 | 34 | 0 | 0.5 | 0.5 | 0 | 2 | 1 | -1 | 27 | 26 | -1 | ε3/ε4 | 1.15 | 1.16 | 0.01 | 2 | 2 |
| 11 | 61-65 | F | 13 | 100 | + | 34 | 40 | 6 | 0.5 | 0.5 | 0 | 1 | 1 | 0 | 25 | 25 | 0 | ε3/ε4 | 1.75 | 1.87 | 0.12 | 3 | 3 |
| 12 | 81-85 | M | 16 | 100 | − | 36 | 36 | 0 | 0.5 | 0.5 | 0 | 4 | 4 | 0 | 16 | 14 | -2 | ε3/ε3 | 1.57 | 1.6 | 0.03 | 3 | 3 |
| 13 | 61-65 | F | 12 | 100 | + | 36 | 38 | 2 | 0.5 | 0.5 | 0 | 5 | 3.5 | -1.5 | 15 | 18 | 3 | ε3/ε3 | 1.59 | 1.52 | -0.07 | 3 | 3 |
| 14 | 81-85 | M | 16 | 97.5 | − | 34 | 32 | -2 | 0 | 0 | 0 | 0 | 0.5 | 0.5 | 29 | 26 | -3 | ε3/ε3 | 1.34 | 1.35 | 0.01 | 3 | 3 |
| 15 | 76-80 | M | 16 | 100 | + | 32 | 34 | 2 | 0.5 | 0.5 | 0 | 0.5 | 1.5 | 1 | 28 | 26 | -2 | ε3/ε3 | 1.37 | 1.34 | -0.03 | 3 | 3 |
| 16 | 81-85 | F | 12 | 100 | − | 34 | 32 | -2 | 0.5 | 0.5 | 0 | 2 | 1 | -1 | 21 | 23 | 2 | ε3/ε4 | 1.88 | 1.88 | 0 | 3 | 3 |

APOE, apolipoprotein E; CDR, Clinical Dementia Rating; CDR SOB, CDR Sum of Boxes; MMSE, Mini-Mental State Examination; BAPL, brain amyloid plaque load on amyloid PET scans; SUVR, standardized uptake value ratio, calculated as cortical uptake relative to cerebellar uptake; Δ, change from baseline to post-intervention; CF, center frequency; ICF+, participants who showed an increase in center frequency after the intervention; ICF-, participants who did not show an increase in center frequency after the intervention.

Supplementary Table 5A. Effect sizes and confidence intervals for resting-state gamma power

| **Electrode** | **ω² (Visit)** | **95% CI (Visit)** | **ω² (Group)** | **95% CI (Group)** | **ω² (Visit×**  **Group)** | **95% CI (Visit×Group)** | **Cohen’s d (Group Baseline)** | **95% CI (Group Baseline)** | **Cohen’s d (ICF+ Pre vs Post)** | **95% CI**  **(ICF+ Pre vs Post)** | **Cohen’s d (ICF- Pre vs Post)** | **95% CI (ICF- Pre vs Post)** |
| --- | --- | --- | --- | --- | --- | --- | --- | --- | --- | --- | --- | --- |
| Fp1 | 0.079 | 0.000-0.419 | 0.266 | 0.000-0.708 | 0.000 | 0.000-0.266 | 0.633 | -1.156-1.095 | 0.480 | -0.393-1.220 | 0.338 | -0.442-1.053 |
| Fz | 0.000 | 0.000-0.272 | 0.154 | 0.000-0.736 | 0.000 | 0.000-0.375 | 0.585 | -1.119-1.087 | -0.076 | -1.550-0.628 | 0.267 | -0.553-1.119 |
| F3 | 0.000 | 0.000-0.283 | 0.174 | 0.000-0.593 | 0.000 | 0.000-0.277 | 0.695 | -1.047-1.203 | 0.175 | -1.104-0.789 | 0.307 | -0.416-0.959 |
| F7 | 0.017 | 0.000-0.306 | 0.055 | 0.000-0.538 | 0.000 | 0.000-0.276 | 0.291 | -1.116-1.134 | 0.362 | -0.526-1.048 | 0.193 | -0.937-0.754 |
| FT9 | 0.064 | 0.000-0.401 | 0.170 | 0.000-0.610 | 0.000 | 0.000-0.305 | 0.673 | -1.085-1.100 | 0.300 | -0.749-1.025 | 0.435 | -0.146-1.291 |
| FC5 | 0.000 | 0.000-0.271 | 0.036 | 0.000-0.454 | 0.000 | 0.000-0.274 | 0.198 | -1.043-1.199 | 0.299 | -0.682-0.984 | 0.008 | -0.896-0.698 |
| FC1 | 0.000 | 0.000-0.408 | 0.093 | 0.000-0.868 | 0.012 | 0.000-0.513 | 0.419 | -1.155-1.048 | -1.017 | -3.404--0.267 | 0.029 | -0.601-0.938 |
| C3 | 0.000 | 0.000-0.279 | 0.182 | 0.000-0.875 | 0.000 | 0.000-0.453 | 0.498 | -1.070-0.981 | -0.362 | -2.280-0.332 | 0.255 | -0.368-1.218 |
| T7 | 0.054 | 0.000-0.535 | 0.144 | 0.000-0.729 | 0.000 | 0.000-0.288 | 0.284 | -1.021-1.070 | 0.605 | 0.114-1.461 | 0.173 | -0.464-1.655 |
| TP9 | 0.041 | 0.000-0.490 | 0.433 | 0.000-0.812 | 0.213 | 0.000-0.635 | 1.173 | -1.199-1.083 | -0.295 | -1.233-0.674 | 0.833 | 0.227-2.553 |
| CP5 | 0.049 | 0.000-0.570 | 0.530 | 0.000-0.845 | 0.000 | 0.000-0.423 | 1.065 | -1.097-1.138 | 0.288 | -0.475-1.365 | 0.407 | -0.198-2.063 |
| CP1 | 0.000 | 0.000-0.425 | 0.204 | 0.000-0.830 | 0.000 | 0.000-0.337 | 0.382 | -1.254-1.061 | 0.076 | -0.559-1.349 | 0.174 | -0.728-0.880 |
| Pz | 0.048 | 0.000-0.448 | 0.237 | 0.000-0.790 | 0.000 | 0.000-0.281 | 0.494 | -1.087-1.036 | 0.430 | -0.314-1.893 | 0.276 | -0.406-1.027 |
| P3 | 0.001 | 0.000-0.456 | 0.443 | 0.000-0.922 | 0.000 | 0.000-0.285 | 0.715 | -1.112-1.114 | 0.303 | -0.457-2.198 | 0.261 | -0.355-1.185 |
| P7 | 0.055 | 0.000-0.473 | 0.390 | 0.000-0.868 | 0.054 | 0.000-0.402 | 0.709 | -1.025-1.005 | 0.002 | -0.833-1.525 | 0.667 | 0.085-1.393 |
| O1 | 0.013 | 0.000-0.303 | 0.484 | 0.000-0.905 | 0.000 | 0.000-0.299 | 0.685 | -1.103-1.045 | 0.107 | -0.876-1.004 | 0.411 | -0.295-1.009 |
| Oz | 0.228 | 0.000-0.607 | 0.358 | 0.000-0.919 | 0.034 | 0.000-0.324 | 0.677 | -1.153-1.162 | 0.401 | -0.359-2.326 | 0.785 | 0.566-1.706 |
| O2 | 0.099 | 0.000-0.402 | 0.109 | 0.000-0.820 | 0.000 | 0.000-0.283 | 0.541 | -1.081-1.041 | 0.361 | -0.671-1.109 | 0.513 | -0.213-1.055 |
| P4 | 0.161 | 0.000-0.508 | 0.183 | 0.000-0.785 | 0.000 | 0.000-0.249 | 0.487 | -1.242-1.078 | 0.456 | -0.309-1.927 | 0.568 | 0.022-1.305 |
| P8 | 0.305 | 0.039-0.618 | 0.235 | 0.000-0.849 | 0.000 | 0.000-0.229 | 0.503 | -1.218-1.048 | 0.682 | 0.025-2.098 | 0.772 | 0.410-1.471 |
| TP10 | 0.255 | 0.000-0.643 | 0.381 | 0.000-0.871 | 0.000 | 0.000-0.370 | 0.578 | -1.038-1.020 | 0.524 | -0.270-1.399 | 0.763 | 0.055-2.719 |
| CP6 | 0.056 | 0.000-0.392 | 0.559 | 0.000-0.921 | 0.000 | 0.000-0.347 | 0.694 | -1.076-1.080 | 0.246 | -0.977-0.907 | 0.485 | -0.111-1.493 |
| CP2 | 0.000 | 0.000-0.476 | 0.519 | 0.000-0.914 | 0.012 | 0.000-0.557 | 0.514 | -1.087-1.108 | 0.305 | -0.448-2.017 | -0.227 | -0.934-0.579 |
| Cz | 0.000 | 0.000-0.451 | 0.054 | 0.000-0.772 | 0.000 | 0.000-0.287 | 0.302 | -1.131-1.115 | 0.123 | -0.566-2.068 | 0.263 | -0.402-1.349 |
| C4 | 0.000 | 0.000-0.239 | 0.400 | 0.000-0.891 | 0.000 | 0.000-0.361 | 0.731 | -1.103-1.036 | -0.043 | -0.631-1.741 | -0.208 | -1.087-0.539 |
| T8 | 0.183 | 0.000-0.509 | 0.054 | 0.000-0.752 | 0.000 | 0.000-0.254 | 0.382 | -1.041-1.077 | 0.746 | 0.187-3.579 | 0.450 | -0.135-1.185 |
| FT10 | 0.245 | 0.000-0.574 | 0.105 | 0.000-0.726 | 0.000 | 0.000-0.311 | 0.431 | -1.052-1.025 | 0.972 | 0.380-5.050 | 0.473 | -0.137-1.158 |
| FC6 | 0.000 | 0.000-0.295 | 0.518 | 0.000-0.907 | 0.080 | 0.000-0.589 | 1.148 | -1.069-1.027 | -0.900 | -2.852--0.205 | 0.313 | -0.294-1.263 |
| FC2 | 0.000 | 0.000-0.279 | 0.126 | 0.000-0.814 | 0.119 | 0.000-0.498 | 0.651 | -1.105-1.138 | -0.439 | -1.205-0.364 | 0.465 | -0.201-1.566 |
| F4 | 0.000 | 0.000-0.550 | 0.094 | 0.000-0.807 | 0.000 | 0.000-0.538 | 0.591 | -1.161-1.119 | 0.047 | -0.926-1.024 | 0.302 | -0.333-1.926 |
| F8 | 0.172 | 0.000-0.626 | 0.318 | 0.000-0.771 | 0.062 | 0.000-0.539 | 0.962 | -1.031-1.074 | 0.270 | -0.463-2.017 | 0.729 | 0.115-1.949 |
| Fp2 | 0.085 | 0.000-0.533 | 0.045 | 0.000-0.636 | 0.000 | 0.000-0.358 | 0.707 | -1.040-1.066 | 0.175 | -0.477-2.615 | 0.596 | -0.061-1.435 |
| AF7 | 0.087 | 0.000-0.507 | 0.094 | 0.000-0.559 | 0.000 | 0.000-0.385 | 0.321 | -1.246-1.140 | 0.457 | -0.250-2.003 | 0.314 | -0.455-0.910 |
| AF3 | 0.090 | 0.000-0.519 | 0.044 | 0.000-0.556 | 0.000 | 0.000-0.414 | 0.597 | -1.000-1.036 | 0.213 | -0.578-1.148 | 0.563 | -0.047-1.459 |
| AFz | 0.058 | 0.000-0.432 | 0.000 | 0.000-0.539 | 0.026 | 0.000-0.398 | 0.518 | -1.002-1.049 | 0.075 | -0.687-1.311 | 0.559 | -0.066-1.361 |
| F1 | 0.000 | 0.000-0.342 | 0.265 | 0.000-0.840 | 0.000 | 0.000-0.329 | 0.568 | -1.147-1.141 | -0.355 | -2.372-0.436 | 0.066 | -0.924-0.763 |
| F5 | 0.000 | 0.000-0.320 | 0.100 | 0.000-0.596 | 0.000 | 0.000-0.229 | 0.493 | -1.059-0.924 | 0.245 | -1.071-0.811 | 0.212 | -0.782-0.813 |
| FT7 | 0.006 | 0.000-0.317 | 0.083 | 0.000-0.529 | 0.000 | 0.000-0.235 | 0.418 | -1.097-1.098 | 0.295 | -0.742-0.932 | 0.228 | -0.702-0.908 |
| FC3 | 0.000 | 0.000-0.266 | 0.239 | 0.000-0.857 | 0.059 | 0.000-0.465 | 0.729 | -1.019-1.044 | -0.974 | -2.151--0.396 | 0.268 | -0.507-0.919 |
| C1 | 0.000 | 0.000-0.263 | 0.073 | 0.000-0.793 | 0.000 | 0.000-0.328 | 0.346 | -1.104-1.100 | -0.176 | -1.019-0.776 | 0.134 | -0.531-1.020 |
| C5 | 0.103 | 0.000-0.458 | 0.434 | 0.000-0.838 | 0.000 | 0.000-0.252 | 0.617 | -1.211-1.139 | 0.377 | -0.511-1.268 | 0.472 | -0.121-1.401 |
| TP7 | 0.000 | 0.000-0.357 | 0.417 | 0.000-0.815 | 0.033 | 0.000-0.548 | 1.065 | -1.133-1.173 | -0.256 | -1.371-0.560 | 0.372 | -0.280-2.178 |
| CP3 | 0.000 | 0.000-0.250 | 0.201 | 0.000-0.734 | 0.000 | 0.000-0.252 | 0.543 | -1.083-1.005 | -0.125 | -0.806-0.888 | 0.113 | -0.575-0.886 |
| P1 | 0.109 | 0.000-0.459 | 0.311 | 0.000-0.824 | 0.000 | 0.000-0.257 | 0.576 | -1.003-1.074 | 0.437 | -0.416-1.429 | 0.427 | -0.297-1.194 |
| P5 | 0.022 | 0.000-0.417 | 0.425 | 0.000-0.868 | 0.000 | 0.000-0.279 | 0.647 | -1.086-1.106 | 0.182 | -0.686-1.172 | 0.392 | -0.271-1.250 |
| PO7 | 0.051 | 0.000-0.418 | 0.401 | 0.000-0.870 | 0.000 | 0.000-0.328 | 0.714 | -1.078-1.174 | 0.124 | -0.672-1.315 | 0.548 | -0.158-1.252 |
| PO3 | 0.023 | 0.000-0.386 | 0.439 | 0.000-0.899 | 0.000 | 0.000-0.246 | 0.727 | -1.088-1.150 | 0.260 | -0.588-1.189 | 0.339 | -0.415-1.003 |
| POz | 0.319 | 0.103-0.600 | 0.400 | 0.000-0.910 | 0.000 | 0.000-0.339 | 0.620 | -1.149-1.080 | 1.051 | 0.693-2.115 | 0.576 | 0.174-1.162 |
| PO4 | 0.203 | 0.000-0.515 | 0.193 | 0.000-0.839 | 0.000 | 0.000-0.261 | 0.592 | -1.211-1.106 | 0.634 | -0.013-1.526 | 0.601 | 0.155-1.190 |
| PO8 | 0.153 | 0.000-0.492 | 0.154 | 0.000-0.807 | 0.000 | 0.000-0.233 | 0.432 | -0.980-1.101 | 0.449 | -0.262-1.680 | 0.558 | -0.023-1.199 |
| P6 | 0.291 | 0.000-0.631 | 0.231 | 0.000-0.836 | 0.000 | 0.000-0.295 | 0.462 | -1.054-1.196 | 0.586 | -0.127-1.810 | 0.801 | 0.409-1.525 |
| P2 | 0.324 | 0.025-0.676 | 0.286 | 0.000-0.840 | 0.000 | 0.000-0.364 | 0.444 | -1.048-0.990 | 0.954 | 0.200-3.292 | 0.602 | 0.074-1.329 |
| CPz | 0.202 | 0.000-0.764 | 0.382 | 0.000-0.908 | 0.000 | 0.000-0.590 | 0.393 | -1.134-1.065 | 0.549 | -0.167-3.723 | 0.593 | -0.030-1.263 |
| CP4 | 0.000 | 0.000-0.291 | 0.671 | 0.000-0.941 | 0.236 | 0.000-0.623 | 0.487 | -1.062-1.045 | 0.561 | -0.155-1.485 | -0.693 | -2.265--0.061 |
| TP8 | 0.245 | 0.000-0.729 | 0.376 | 0.000-0.891 | 0.000 | 0.000-0.451 | 0.567 | -1.130-1.007 | 0.483 | -0.231-1.594 | 0.765 | 0.088-3.422 |
| C6 | 0.000 | 0.000-0.322 | 0.286 | 0.000-0.783 | 0.000 | 0.000-0.337 | 0.431 | -1.083-1.107 | 0.445 | -0.630-1.170 | -0.055 | -0.813-0.774 |
| C2 | 0.000 | 0.000-0.469 | 0.208 | 0.000-0.827 | 0.000 | 0.000-0.259 | 0.522 | -0.995-1.104 | 0.157 | -0.505-1.374 | 0.328 | -0.346-1.534 |
| FC4 | 0.000 | 0.000-0.245 | 0.512 | 0.000-0.892 | 0.409 | 0.110-0.791 | 1.351 | -1.053-1.018 | -2.096 | -5.045--1.622 | 0.630 | 0.041-1.958 |
| FT8 | 0.213 | 0.000-0.639 | 0.220 | 0.000-0.734 | 0.000 | 0.000-0.257 | 0.411 | -1.133-1.063 | 0.786 | 0.128-2.655 | 0.423 | -0.225-1.597 |
| F6 | 0.112 | 0.000-0.748 | 0.109 | 0.000-0.774 | 0.036 | 0.000-0.650 | 0.780 | -1.160-1.124 | 0.329 | -0.481-1.251 | 0.589 | -0.035-2.387 |
| AF8 | 0.116 | 0.000-0.618 | 0.052 | 0.000-0.639 | 0.019 | 0.000-0.337 | 0.791 | -1.100-1.041 | 0.148 | -0.477-3.449 | 0.766 | 0.203-1.960 |
| AF4 | 0.138 | 0.000-0.513 | 0.312 | 0.000-0.824 | 0.029 | 0.000-0.402 | 0.781 | -1.108-1.074 | 0.244 | -0.570-1.245 | 0.663 | 0.090-1.596 |
| F2 | 0.114 | 0.000-0.610 | 0.496 | 0.000-0.902 | 0.369 | 0.067-0.807 | 1.005 | -1.053-1.022 | -0.750 | -3.124--0.052 | 1.001 | 0.406-2.663 |

Note. Electrode refers to individual scalp EEG channels analyzed for resting-state gamma power

Supplementary Table 5B. Effect sizes and confidence intervals for event-related synchronization

| **Electrode** | **ω² (Visit)** | **95% CI (Visit)** | **ω²**  **(Group)** | **95% CI (Group)** | **ω² (Visit×**  **Group)** | **95% CI (Visit×Group)** | **Cohen’s d (Group Baseline)** | **95% CI (Group Baseline)** | **Cohen’s d (ICF+ Pre vs Post)** | **95% CI**  **(ICF+ Pre vs Post)** | **Cohen’s d (ICF- Pre vs Post)** | **95% CI (ICF- Pre vs Post)** |
| --- | --- | --- | --- | --- | --- | --- | --- | --- | --- | --- | --- | --- |
| Fp1 | 0.373 | 0.067-0.753 | 0.012 | 0.000-0.419 | 0.000 | 0.000-0.390 | 0.499 | -0.569-1.763 | -0.707 | -2.081--0.101 | -1.083 | -1.872--0.741 |
| Fz | 0.509 | 0.202-0.802 | 0.102 | 0.000-0.272 | 0.001 | 0.000-0.348 | 0.770 | -0.132-1.705 | -1.367 | -2.787--0.935 | -0.778 | -2.766--0.128 |
| F3 | 0.526 | 0.213-0.808 | 0.071 | 0.000-0.283 | 0.000 | 0.000-0.312 | 0.686 | -0.306-1.759 | -1.146 | -3.007--0.414 | -1.002 | -2.488--0.375 |
| F7 | 0.254 | 0.000-0.618 | 0.117 | 0.000-0.306 | 0.000 | 0.000-0.292 | 0.531 | -0.592-1.528 | -0.533 | -1.584-0.189 | -0.763 | -1.976--0.203 |
| FT9 | 0.090 | 0.000-0.465 | 0.316 | 0.000-0.401 | 0.000 | 0.000-0.309 | 1.026 | 0.230-2.214 | -0.379 | -1.231-0.528 | -0.438 | -1.191-0.200 |
| FC5 | 0.305 | 0.000-0.723 | 0.061 | 0.000-0.271 | 0.000 | 0.000-0.310 | 0.562 | -0.451-1.737 | -0.622 | -2.256-0.051 | -0.857 | -2.186--0.264 |
| FC1 | 0.473 | 0.204-0.729 | 0.004 | 0.000-0.408 | 0.000 | 0.000-0.236 | 0.359 | -0.856-1.253 | -1.096 | -2.384--0.667 | -0.892 | -2.004--0.369 |
| C3 | 0.463 | 0.234-0.690 | 0.041 | 0.000-0.279 | 0.000 | 0.000-0.259 | 0.444 | -0.552-1.580 | -1.033 | -2.687--0.769 | -0.907 | -1.833--0.386 |
| T7 | 0.038 | 0.000-0.414 | 0.337 | 0.000-0.535 | 0.000 | 0.000-0.314 | 1.057 | 0.213-2.820 | -0.680 | -1.582--0.183 | -0.128 | -0.881-0.701 |
| TP9 | 0.214 | 0.000-0.620 | 0.095 | 0.000-0.490 | 0.000 | 0.000-0.410 | 0.574 | -0.396-1.806 | -0.635 | -1.552-0.047 | -0.491 | -2.595-0.119 |
| CP5 | 0.119 | 0.000-0.450 | 0.001 | 0.000-0.570 | 0.002 | 0.000-0.315 | -0.001 | -1.014-1.222 | -0.253 | -1.388-0.489 | -0.614 | -1.273--0.186 |
| CP1 | 0.267 | 0.016-0.628 | 0.000 | 0.000-0.425 | 0.020 | 0.000-0.468 | 0.380 | -0.551-1.589 | -1.294 | -2.659--0.889 | -0.317 | -1.035-0.378 |
| Pz | 0.524 | 0.262-0.782 | 0.000 | 0.000-0.448 | 0.008 | 0.000-0.490 | 0.689 | -0.196-1.982 | -1.248 | -2.757--0.638 | -0.880 | -1.743--0.524 |
| P3 | 0.385 | 0.105-0.703 | 0.000 | 0.000-0.456 | 0.000 | 0.000-0.316 | -0.053 | -1.299-1.083 | -0.788 | -2.001--0.184 | -0.885 | -1.738--0.419 |
| P7 | 0.282 | 0.012-0.595 | 0.000 | 0.000-0.473 | 0.000 | 0.000-0.243 | 0.186 | -1.052-1.244 | -0.680 | -1.786--0.046 | -0.685 | -1.420--0.140 |
| O1 | 0.299 | 0.000-0.684 | 0.000 | 0.000-0.303 | 0.115 | 0.000-0.537 | -0.431 | -1.707-0.547 | -0.245 | -1.337-0.616 | -1.216 | -2.643--0.664 |
| Oz | 0.427 | 0.077-0.861 | 0.043 | 0.000-0.607 | 0.000 | 0.000-0.367 | 0.466 | -0.461-1.801 | -0.696 | -3.532--0.017 | -1.361 | -2.487--0.892 |
| O2 | 0.542 | 0.241-0.804 | 0.000 | 0.000-0.402 | 0.000 | 0.000-0.378 | 0.403 | -0.682-1.641 | -1.206 | -4.925--0.424 | -1.020 | -1.977--0.595 |
| P4 | 0.457 | 0.204-0.722 | 0.000 | 0.000-0.508 | 0.000 | 0.000-0.401 | 0.154 | -0.900-1.283 | -1.078 | -2.212--0.574 | -0.792 | -1.892--0.341 |
| P8 | 0.384 | 0.089-0.668 | 0.053 | 0.039-0.618 | 0.089 | 0.000-0.469 | 0.707 | -0.284-1.788 | -1.062 | -2.478--0.575 | -0.506 | -1.498-0.129 |
| TP10 | 0.191 | 0.000-0.575 | 0.123 | 0.000-0.643 | 0.000 | 0.000-0.578 | 0.475 | -0.630-1.575 | -1.258 | -2.940--0.683 | -0.256 | -0.754-0.572 |
| CP6 | 0.266 | 0.000-0.639 | 0.229 | 0.000-0.392 | 0.000 | 0.000-0.242 | 0.472 | -0.514-1.814 | -0.688 | -2.160--0.094 | -0.653 | -2.212--0.029 |
| CP2 | 0.155 | 0.000-0.577 | 0.021 | 0.000-0.476 | 0.000 | 0.000-0.279 | 0.549 | -0.430-1.859 | -0.680 | -2.244-0.038 | -0.403 | -1.349-0.273 |
| Cz | 0.228 | 0.000-0.641 | 0.304 | 0.000-0.451 | 0.000 | 0.000-0.400 | 0.760 | -0.149-1.834 | -0.533 | -1.489-0.215 | -0.678 | -2.081--0.064 |
| C4 | 0.308 | 0.098-0.595 | 0.253 | 0.000-0.239 | 0.000 | 0.000-0.461 | 0.436 | -0.818-1.333 | -0.434 | -1.196-0.267 | -1.207 | -2.061--0.876 |
| T8 | 0.602 | 0.405-0.798 | 0.447 | 0.000-0.509 | 0.044 | 0.000-0.488 | 1.189 | 0.321-3.003 | -1.453 | -3.343--0.939 | -1.014 | -1.964--0.640 |
| FT10 | 0.511 | 0.237-0.775 | 0.274 | 0.000-0.574 | 0.072 | 0.000-0.559 | 1.034 | 0.165-2.418 | -1.340 | -3.263--0.667 | -0.726 | -1.541--0.226 |
| FC6 | 0.482 | 0.161-0.757 | 0.263 | 0.000-0.295 | 0.000 | 0.000-0.324 | 0.896 | -0.031-2.433 | -1.056 | -2.322--0.514 | -0.928 | -1.843--0.387 |
| FC2 | 0.479 | 0.183-0.780 | 0.194 | 0.000-0.279 | 0.000 | 0.000-0.332 | 0.549 | -0.484-1.714 | -0.802 | -1.975--0.214 | -1.274 | -4.863--0.584 |
| F4 | 0.528 | 0.182-0.821 | 0.128 | 0.000-0.550 | 0.000 | 0.000-0.284 | 0.402 | -0.572-1.795 | -0.887 | -2.966--0.160 | -1.347 | -3.647--0.704 |
| F8 | 0.720 | 0.509-0.878 | 0.353 | 0.000-0.626 | 0.000 | 0.000-0.295 | 0.917 | -0.012-2.412 | -1.661 | -3.306--1.169 | -1.594 | -4.178--0.978 |
| Fp2 | 0.454 | 0.069-0.826 | 0.162 | 0.000-0.533 | 0.000 | 0.000-0.392 | 0.545 | -0.479-2.492 | -0.844 | -7.499--0.108 | -1.097 | -2.479--0.504 |
| AF7 | 0.336 | 0.039-0.684 | 0.302 | 0.000-0.507 | 0.000 | 0.000-0.374 | 0.933 | -0.036-2.205 | -0.809 | -2.311--0.207 | -0.675 | -1.660--0.085 |
| AF3 | 0.532 | 0.220-0.797 | 0.148 | 0.000-0.519 | 0.010 | 0.000-0.442 | 0.852 | -0.158-2.038 | -1.090 | -2.743--0.487 | -1.118 | -2.437--0.593 |
| AFz | 0.466 | 0.198-0.709 | 0.202 | 0.000-0.432 | 0.156 | 0.000-0.538 | 1.022 | 0.163-2.166 | -1.206 | -2.791--0.693 | -0.597 | -1.347-0.019 |
| F1 | 0.576 | 0.291-0.848 | 0.106 | 0.000-0.342 | 0.000 | 0.000-0.345 | 0.758 | -0.117-1.670 | -1.261 | -3.200--0.759 | -1.096 | -3.788--0.336 |
| F5 | 0.459 | 0.104-0.756 | 0.075 | 0.000-0.320 | 0.000 | 0.000-0.328 | 0.723 | -0.239-1.707 | -0.996 | -2.872--0.352 | -0.901 | -2.588--0.249 |
| FT7 | 0.263 | 0.005-0.575 | 0.265 | 0.000-0.317 | 0.000 | 0.000-0.327 | 0.923 | 0.085-2.292 | -0.677 | -1.512--0.148 | -0.610 | -1.531--0.106 |
| FC3 | 0.510 | 0.326-0.745 | 0.092 | 0.000-0.266 | 0.000 | 0.000-0.285 | 0.550 | -0.479-1.399 | -1.132 | -3.895--0.942 | -0.980 | -1.951--0.485 |
| C1 | 0.322 | 0.026-0.692 | 0.115 | 0.000-0.263 | 0.000 | 0.000-0.265 | 0.599 | -0.405-1.985 | -0.893 | -2.063--0.315 | -0.672 | -1.810-0.024 |
| C5 | 0.195 | 0.000-0.541 | 0.217 | 0.000-0.458 | 0.000 | 0.000-0.390 | 0.641 | -0.340-1.622 | -0.634 | -1.448--0.033 | -0.418 | -1.018-0.243 |
| TP7 | 0.174 | 0.000-0.538 | 0.000 | 0.000-0.357 | 0.067 | 0.000-0.453 | 0.518 | -0.423-1.698 | -0.653 | -1.565--0.001 | -0.268 | -1.883-0.323 |
| CP3 | 0.302 | 0.000-0.679 | 0.000 | 0.000-0.250 | 0.000 | 0.000-0.292 | -0.019 | -1.255-0.950 | -0.555 | -2.127-0.212 | -0.865 | -2.063--0.251 |
| P1 | 0.314 | 0.035-0.629 | 0.000 | 0.000-0.459 | 0.000 | 0.000-0.265 | 0.389 | -0.673-1.823 | -0.781 | -1.640--0.241 | -0.653 | -1.379--0.118 |
| P5 | 0.329 | 0.009-0.641 | 0.000 | 0.000-0.417 | 0.000 | 0.000-0.318 | -0.054 | -1.433-0.945 | -0.672 | -1.462--0.112 | -0.848 | -1.998--0.264 |
| PO7 | 0.368 | 0.027-0.705 | 0.000 | 0.103-0.600 | 0.000 | 0.000-0.310 | -0.122 | -1.288-0.982 | -0.782 | -2.065--0.065 | -0.859 | -1.899--0.298 |
| PO3 | 0.351 | 0.050-0.695 | 0.000 | 0.000-0.515 | 0.000 | 0.000-0.369 | -0.303 | -1.671-0.630 | -0.503 | -1.499-0.214 | -1.114 | -2.294--0.580 |
| POz | 0.489 | 0.092-0.860 | 0.000 | 0.000-0.492 | 0.000 | 0.000-0.366 | 0.276 | -0.744-1.477 | -0.850 | -4.342--0.098 | -1.298 | -2.541--0.795 |
| PO4 | 0.569 | 0.258-0.840 | 0.000 | 0.000-0.631 | 0.000 | 0.000-0.425 | 0.379 | -0.575-1.695 | -1.340 | -11.006--0.537 | -1.042 | -2.125--0.558 |
| PO8 | 0.394 | 0.111-0.724 | 0.084 | 0.025-0.676 | 0.000 | 0.000-0.345 | 0.461 | -0.672-1.769 | -0.712 | -1.660--0.105 | -1.111 | -2.587--0.625 |
| P6 | 0.517 | 0.284-0.749 | 0.062 | 0.000-0.764 | 0.028 | 0.000-0.426 | 0.598 | -0.430-1.825 | -1.140 | -2.540--0.659 | -0.949 | -1.843--0.516 |
| P2 | 0.599 | 0.395-0.831 | 0.000 | 0.000-0.291 | 0.043 | 0.000-0.506 | 0.528 | -0.447-1.619 | -1.618 | -3.542--1.142 | -0.914 | -2.407--0.440 |
| CPz | 0.234 | 0.000-0.607 | 0.000 | 0.000-0.729 | 0.047 | 0.000-0.548 | 0.844 | -0.090-2.320 | -0.966 | -2.250--0.375 | -0.270 | -0.979-0.431 |
| CP4 | 0.077 | 0.000-0.416 | 0.127 | 0.000-0.322 | 0.000 | 0.000-0.489 | 0.361 | -0.749-1.472 | -0.147 | -0.831-1.142 | -0.807 | -1.707--0.280 |
| TP8 | 0.364 | 0.102-0.663 | 0.117 | 0.000-0.469 | 0.014 | 0.000-0.500 | 0.509 | -0.491-1.747 | -1.092 | -2.840--0.638 | -0.521 | -1.140-0.104 |
| C6 | 0.236 | 0.000-0.688 | 0.258 | 0.000-0.245 | 0.000 | 0.000-0.273 | 0.503 | -0.606-1.516 | -0.519 | -2.093-0.224 | -0.723 | -1.595--0.165 |
| C2 | 0.257 | 0.000-0.710 | 0.142 | 0.000-0.639 | 0.022 | 0.000-0.672 | 0.202 | -1.206-1.070 | -0.388 | -0.873-0.625 | -0.877 | -2.801--0.130 |
| FC4 | 0.532 | 0.246-0.784 | 0.262 | 0.000-0.748 | 0.000 | 0.000-0.360 | 0.424 | -0.795-1.508 | -0.787 | -1.887--0.244 | -1.646 | -3.211--1.124 |
| FT8 | 0.629 | 0.456-0.807 | 0.417 | 0.000-0.618 | 0.120 | 0.000-0.574 | 1.233 | 0.337-2.723 | -1.857 | -4.028--1.508 | -0.863 | -1.822--0.390 |
| F6 | 0.724 | 0.556-0.877 | 0.304 | 0.000-0.513 | 0.000 | 0.000-0.334 | 0.833 | -0.116-2.037 | -1.730 | -3.540--1.295 | -1.553 | -4.226--0.883 |
| AF8 | 0.751 | 0.592-0.885 | 0.645 | 0.000-0.610 | 0.000 | 0.000-0.282 | 1.119 | 0.171-2.699 | -1.665 | -3.516--1.170 | -1.870 | -4.390--1.229 |
| AF4 | 0.486 | 0.193-0.784 | 0.005 | 0.000-0.902 | 0.000 | 0.000-0.335 | 0.152 | -1.058-1.299 | -0.821 | -4.641--0.108 | -1.228 | -2.571--0.735 |
| F2 | 0.547 | 0.245-0.839 | 0.166 | 0.000-0.879 | 0.000 | 0.000-0.325 | 0.452 | -0.685-1.631 | -0.903 | -2.889--0.241 | -1.495 | -4.933--0.785 |

Note. Electrode refers to individual scalp EEG channels analyzed for gamma-band event-related synchronization

Supplementary Table 5C. Effect sizes and confidence intervals for spectral Granger causality

| **ROI** | **ω² (Visit)** | **95% CI (Visit)** | **ω² (Group)** | **95% CI (Group)** | **ω² (Visit×**  **Group)** | **95% CI (Visit×Group)** | **Cohen’s d (Group Baseline)** | **95% CI (Group Baseline)** | **Cohen’s d (ICF+ Pre vs Post)** | **95% CI**  **(ICF+ Pre vs Post)** | **Cohen’s d (ICF- Pre vs Post)** | **95% CI (ICF- Pre vs Post)** |
| --- | --- | --- | --- | --- | --- | --- | --- | --- | --- | --- | --- | --- |
| LF_LF | 0.000 | 0.000-0.272 | 0.000 | 0.000-0.706 | 0.012 | 0.000-0.447 | 0.439 | -0.976-1.152 | -0.276 | -1.464-0.485 | -0.204 | -0.86-0.879 |
| LF_LT | 0.000 | 0.000-0.348 | 0.000 | 0.000-0.532 | 0.000 | 0.000-0.353 | 0.249 | -1.055-1.038 | 0.119 | -0.900-0.822 | -0.216 | -1.16-0.444 |
| LF_LC | 0.067 | 0.000-0.427 | 0.000 | 0.000-0.431 | 0.000 | 0.000-0.624 | 0.314 | -0.973-1.18 | -0.298 | -1.247-0.466 | -0.397 | -0.98-0.411 |
| LF_LP | 0.135 | 0.000-0.508 | 0.001 | 0.000-0.234 | 0.030 | 0.000-0.587 | 0.195 | -0.915-1.042 | -0.138 | -0.823-0.931 | -1.178 | -2.795--0.599 |
| LF_pF | 0.000 | 0.000-0.409 | 0.000 | 0.000-0.632 | 0.000 | 0.000-0.281 | 0.419 | -1.082-1.165 | -0.073 | -1.142-0.595 | -0.570 | -1.286--0.008 |
| LF_F | 0.135 | 0.000-0.450 | 0.000 | 0.000-0.175 | 0.000 | 0.000-0.298 | -0.182 | -1.042-1.089 | -0.281 | -1.104-0.651 | -0.562 | -3.018--0.135 |
| LF_C | 0.131 | 0.000-0.544 | 0.119 | 0.000-0.447 | 0.000 | 0.000-0.371 | 0.985 | -1.042-1.051 | -0.363 | -1.910-0.329 | -0.691 | -1.73--0.097 |
| LF_P | 0.050 | 0.000-0.544 | 0.000 | 0.000-0.119 | 0.000 | 0.000-0.496 | 0.251 | -1.104-1.123 | -0.372 | -1.043-0.530 | -0.291 | -1.391-0.406 |
| LF_O | 0.000 | 0.000-0.436 | 0.000 | 0.000-0.346 | 0.000 | 0.000-0.370 | 0.186 | -1.044-0.957 | -0.237 | -1.991-0.482 | -0.014 | -0.95-0.739 |
| LF_RF | 0.149 | 0.000-0.411 | 0.000 | 0.000-0.132 | 0.000 | 0.000-0.305 | -0.005 | -1.025-1.126 | -0.187 | -1.007-0.751 | -0.673 | -1.378--0.406 |
| LF_RT | 0.339 | 0.109-0.819 | 0.000 | 0.000-0.297 | 0.000 | 0.000-0.542 | 0.253 | -1.054-0.991 | -0.655 | -1.532--0.456 | -1.098 | -3.366--0.393 |
| LF_RC | 0.220 | 0.091-0.714 | 0.000 | 0.000-0.314 | 0.000 | 0.000-0.269 | 0.621 | -0.913-0.978 | -0.667 | -1.844--0.503 | -1.055 | -2.494--0.561 |
| LF_RP | 0.000 | 0.000-0.450 | 0.010 | 0.000-0.292 | 0.000 | 0.000-0.376 | -0.234 | -0.964-1.121 | -0.448 | -1.261-0.325 | -0.099 | -0.875-0.595 |
| LT_LF | 0.000 | 0.000-0.466 | 0.074 | 0.000-0.446 | 0.141 | 0.000-0.555 | 0.282 | -1.008-1.091 | -0.167 | -1.949-0.524 | 0.089 | -1.253-0.55 |
| LT_LT | 0.000 | 0.000-0.312 | 0.234 | 0.000-0.746 | 0.000 | 0.000-0.386 | 1.204 | -1.032-1.158 | -0.187 | -1.279-0.713 | 0.393 | -0.242-1.737 |
| LT_LC | 0.000 | 0.000-0.274 | 0.000 | 0.000-0.216 | 0.012 | 0.000-0.305 | 0.662 | -0.904-1.146 | -0.476 | -1.766-0.192 | -0.091 | -0.664-0.916 |
| LT_LP | 0.000 | 0.000-0.380 | 0.060 | 0.000-0.536 | 0.000 | 0.000-0.184 | 0.666 | -1.01-1.062 | 0.116 | -1.426-0.784 | -0.059 | -1.321-0.543 |
| LT_pF | 0.000 | 0.000-0.275 | 0.000 | 0.000-0.222 | 0.000 | 0.000-0.348 | 0.628 | -1.028-1.055 | -0.319 | -1.114-0.588 | -0.187 | -0.857-0.637 |
| LT_F | 0.000 | 0.000-0.248 | 0.032 | 0.000-0.418 | 0.048 | 0.000-0.470 | 0.284 | -1.066-1.067 | -0.199 | -0.978-0.710 | -0.234 | -0.779-0.793 |
| LT_C | 0.000 | 0.000-0.250 | 0.082 | 0.000-0.538 | 0.014 | 0.000-0.412 | 0.188 | -1.037-1.064 | 0.501 | -0.243-1.933 | -0.271 | -0.987-0.561 |
| LT_P | 0.019 | 0.000-0.314 | 0.002 | 0.000-0.421 | 0.000 | 0.000-0.320 | 0.336 | -1.02-1.098 | 0.471 | -0.391-1.123 | 0.251 | -1.28-0.719 |
| LT_O | 0.000 | 0.000-0.215 | 0.000 | 0.000-0.264 | 0.000 | 0.000-0.131 | 0.667 | -0.921-1.133 | 0.097 | -0.839-0.823 | 0.090 | -0.997-0.592 |
| LT_RF | 0.035 | 0.000-0.492 | 0.036 | 0.000-0.326 | 0.046 | 0.000-0.456 | 0.755 | -1.068-1.178 | -0.398 | -1.851-0.308 | -0.480 | -2.857-0.169 |
| LT_RT | 0.068 | 0.000-0.296 | 0.000 | 0.000-0.087 | 0.000 | 0.000-0.142 | -0.249 | -0.843-1.05 | -0.415 | -1.222-0.558 | -0.410 | -1.05-0.018 |
| LT_RC | 0.128 | 0.000-0.445 | 0.025 | 0.000-0.438 | 0.000 | 0.000-0.439 | 0.180 | -0.996-1.096 | -0.617 | -1.471-0.001 | -0.483 | -1.304--0.003 |
| LT_RP | 0.000 | 0.000-0.271 | 0.009 | 0.000-0.606 | 0.064 | 0.000-0.512 | 1.122 | -0.997-1.039 | -0.726 | -1.756--0.212 | 0.387 | -1.067-0.784 |
| LC_LF | 0.000 | 0.000-0.422 | 0.000 | 0.000-0.246 | 0.000 | 0.000-0.326 | -0.003 | -1.102-1.219 | 0.054 | -1.089-0.898 | -0.228 | -1.315-0.435 |
| LC_LT | 0.000 | 0.000-0.254 | 0.058 | 0.000-0.455 | 0.135 | 0.000-0.523 | 0.235 | -0.99-1.123 | 0.449 | -0.280-1.356 | -0.274 | -0.831-0.853 |
| LC_LC | 0.278 | 0.023-0.582 | 0.000 | 0.000-0.423 | 0.000 | 0.000-0.233 | 0.253 | -1.121-1.155 | -0.600 | -2.492-0.106 | -0.696 | -1.361--0.25 |
| LC_LP | 0.123 | 0.000-0.471 | 0.001 | 0.000-0.274 | 0.032 | 0.000-0.301 | -0.068 | -1.017-1.175 | -0.031 | -0.769-0.981 | -0.637 | -1.485--0.069 |
| LC_pF | 0.029 | 0.000-0.624 | 0.000 | 0.000-0.391 | 0.000 | 0.000-0.244 | -0.435 | -0.951-1.033 | 0.190 | -0.930-0.911 | -1.167 | -2.107--0.764 |
| LC_F | 0.077 | 0.000-0.368 | 0.000 | 0.000-0.241 | 0.000 | 0.000-0.315 | -0.684 | -0.889-0.998 | 0.252 | -0.836-1.064 | -0.573 | -1.157--0.383 |
| LC_C | 0.153 | 0.000-0.459 | 0.000 | 0.000-0.290 | 0.122 | 0.000-0.498 | -0.220 | -0.977-1.058 | 0.076 | -0.893-1.100 | -0.692 | -1.348--0.271 |
| LC_P | 0.000 | 0.000-0.392 | 0.000 | 0.000-0.381 | 0.000 | 0.000-0.242 | 0.019 | -1.047-1.198 | -0.039 | -1.939-0.735 | 0.044 | -1.172-0.561 |
| LC_O | 0.034 | 0.000-0.313 | 0.106 | 0.000-0.506 | 0.000 | 0.000-0.183 | 0.149 | -1.004-1.171 | -0.097 | -1.167-0.795 | -0.498 | -1.015--0.027 |
| LC_RF | 0.108 | 0.000-0.669 | 0.000 | 0.000-0.279 | 0.000 | 0.000-0.245 | -0.462 | -0.982-1.148 | -0.007 | -1.485-0.681 | -1.172 | -3.542--0.577 |
| LC_RT | 0.431 | 0.111-0.732 | 0.000 | 0.000-0.255 | 0.000 | 0.000-0.279 | -0.203 | -0.99-1.058 | -0.569 | -1.714-0.134 | -1.309 | -3.074--0.682 |
| LC_RC | 0.382 | 0.078-0.677 | 0.000 | 0.000-0.220 | 0.000 | 0.000-0.282 | -0.028 | -1.021-1.022 | -0.681 | -3.018--0.269 | -0.999 | -2.125--0.517 |
| LC_RP | 0.000 | 0.000-0.279 | 0.040 | 0.000-0.373 | 0.045 | 0.000-0.421 | 0.650 | -0.995-1.105 | -0.414 | -1.324-0.351 | 0.110 | -0.756-0.638 |
| LP_LF | 0.406 | 0.000-0.799 | 0.200 | 0.000-0.793 | 0.340 | 0.000-0.841 | -0.668 | -1.025-1.067 | -0.149 | -1.086-0.565 | -0.808 | -1.745--0.283 |
| LP_LT | 0.000 | 0.000-0.487 | 0.030 | 0.000-0.447 | 0.001 | 0.000-0.301 | -0.568 | -0.988-1.136 | 0.283 | -0.619-0.935 | -0.429 | -1.474-0.198 |
| LP_LC | 0.220 | 0.000-0.597 | 0.000 | 0.000-0.208 | 0.000 | 0.000-0.229 | -0.263 | -1.027-1.105 | -0.776 | -2.028--0.181 | -0.579 | -1.472-0.044 |
| LP_LP | 0.196 | 0.000-0.488 | 0.114 | 0.000-0.674 | 0.000 | 0.000-0.285 | -0.423 | -1.064-1.058 | -0.314 | -1.066-0.614 | -0.697 | -1.275--0.24 |
| LP_pF | 0.326 | 0.019-0.579 | 0.091 | 0.000-0.540 | 0.198 | 0.000-0.567 | -0.727 | -1.132-1.181 | -0.030 | -1.095-0.840 | -0.836 | -1.582--0.442 |
| LP_F | 0.114 | 0.000-0.463 | 0.000 | 0.000-0.273 | 0.051 | 0.000-0.392 | -0.634 | -0.881-0.95 | 0.650 | 0.003-2.042 | -0.541 | -1.405--0.341 |
| LP_C | 0.131 | 0.000-0.410 | 0.000 | 0.000-0.099 | 0.000 | 0.000-0.108 | -0.446 | -0.944-1.078 | -0.328 | -0.972-0.579 | -0.568 | -1.249--0.019 |
| LP_P | 0.000 | 0.000-0.447 | 0.000 | 0.000-0.088 | 0.000 | 0.000-0.116 | 0.123 | -1.018-1.147 | -0.496 | -1.232-0.354 | 0.224 | -1.033-0.605 |
| LP_O | 0.000 | 0.000-0.251 | 0.000 | 0.000-0.072 | 0.000 | 0.000-0.123 | -0.134 | -1.107-1.096 | 0.062 | -1.540-0.707 | -0.079 | -1.035-0.605 |
| LP_RF | 0.110 | 0.000-0.444 | 0.000 | 0.000-0.281 | 0.065 | 0.000-0.377 | -0.642 | -0.928-1.033 | 0.207 | -1.355-0.777 | -0.561 | -1.264--0.299 |
| LP_RT | 0.282 | 0.000-0.609 | 0.040 | 0.000-0.364 | 0.060 | 0.000-0.345 | -0.582 | -0.917-1.075 | -0.433 | -2.659-0.311 | -0.704 | -1.572--0.366 |
| LP_RC | 0.221 | 0.000-0.549 | 0.000 | 0.000-0.277 | 0.000 | 0.000-0.289 | -0.569 | -0.906-1.133 | -0.547 | -1.620-0.183 | -0.648 | -1.686--0.384 |
| LP_RP | 0.000 | 0.000-0.572 | 0.000 | 0.000-0.256 | 0.000 | 0.000-0.163 | -0.387 | -1.049-1.191 | -0.083 | -3.081-0.507 | 0.160 | -1.316-0.6 |
| pF_LF | 0.013 | 0.000-0.431 | 0.000 | 0.000-0.885 | 0.099 | 0.000-0.587 | 0.428 | -0.914-1.068 | -0.401 | -0.886-0.841 | -0.621 | -1.528--0.011 |
| pF_LT | 0.000 | 0.000-0.249 | 0.000 | 0.000-0.525 | 0.149 | 0.000-0.697 | 0.686 | -0.965-1.156 | -0.187 | -1.031-0.825 | -0.053 | -0.992-0.484 |
| pF_LC | 0.000 | 0.000-0.270 | 0.000 | 0.000-0.497 | 0.092 | 0.000-0.545 | 0.614 | -0.837-1.027 | -0.278 | -0.803-1.134 | -0.021 | -1.421-0.6 |
| pF_LP | 0.016 | 0.000-0.464 | 0.000 | 0.000-0.220 | 0.000 | 0.000-0.332 | -0.186 | -1.005-1.127 | 0.095 | -0.878-1.068 | -0.620 | -1.476--0.048 |
| pF_pF | 0.066 | 0.000-0.454 | 0.000 | 0.000-0.697 | 0.000 | 0.000-0.604 | 0.287 | -0.892-0.971 | -0.393 | -1.025-0.461 | -0.547 | -1.595--0.42 |
| pF_F | 0.036 | 0.000-0.584 | 0.070 | 0.000-0.544 | 0.059 | 0.000-0.466 | 0.676 | -0.938-0.981 | -0.399 | -0.897-0.388 | -0.908 | -2.385--0.235 |
| pF_C | 0.008 | 0.000-0.425 | 0.000 | 0.000-0.370 | 0.000 | 0.000-0.324 | 0.490 | -0.996-1.124 | -0.221 | -1.437-0.518 | -0.486 | -1.54-0.195 |
| pF_P | 0.000 | 0.000-0.235 | 0.000 | 0.000-0.176 | 0.000 | 0.000-0.161 | 0.778 | -0.879-1.102 | -0.397 | -1.070-0.751 | 0.258 | -0.446-1.086 |
| pF_O | 0.000 | 0.000-0.270 | 0.015 | 0.000-0.473 | 0.000 | 0.000-0.231 | 0.940 | -1.001-1.087 | -0.049 | -0.779-1.227 | 0.092 | -0.638-0.844 |
| pF_RF | 0.000 | 0.000-0.708 | 0.000 | 0.000-0.759 | 0.028 | 0.000-0.624 | 0.359 | -0.848-1.015 | -0.411 | -0.865-0.951 | -0.134 | -2.386-0.419 |
| pF_RT | 0.283 | 0.000-0.713 | 0.000 | 0.000-0.459 | 0.000 | 0.000-0.624 | -0.337 | -0.996-1.133 | -0.501 | -1.059-0.519 | -0.775 | -1.828--0.228 |
| pF_RC | 0.210 | 0.000-0.547 | 0.000 | 0.000-0.322 | 0.000 | 0.000-0.481 | 0.071 | -0.987-1.179 | -0.558 | -2.013--0.279 | -0.639 | -1.328--0.116 |
| pF_RP | 0.000 | 0.000-0.230 | 0.000 | 0.000-0.154 | 0.000 | 0.000-0.151 | 0.531 | -0.858-0.984 | -0.404 | -1.010-0.342 | 0.123 | -0.857-0.616 |
| F_LF | 0.009 | 0.000-0.332 | 0.140 | 0.000-0.623 | 0.173 | 0.000-0.646 | 0.230 | -1.056-1.126 | -0.057 | -0.843-1.099 | 0.189 | -0.848-0.646 |
| F_LT | 0.031 | 0.000-0.303 | 0.000 | 0.000-0.672 | 0.000 | 0.000-0.591 | 0.707 | -1.003-1.035 | 0.406 | -0.611-1.090 | 0.182 | -0.886-0.672 |
| F_LC | 0.000 | 0.000-0.266 | 0.000 | 0.000-0.297 | 0.000 | 0.000-0.381 | 0.520 | -0.989-1.043 | -0.158 | -0.985-1.032 | -0.007 | -0.945-0.604 |
| F_LP | 0.000 | 0.000-0.230 | 0.113 | 0.000-0.471 | 0.000 | 0.000-0.118 | 0.552 | -1.062-1.123 | 0.203 | -1.137-0.905 | -0.163 | -0.856-0.718 |
| F_pF | 0.000 | 0.000-0.204 | 0.000 | 0.000-0.305 | 0.000 | 0.000-0.164 | -0.042 | -0.985-1.066 | 0.346 | -0.537-1.085 | -0.376 | -0.956-0.214 |
| F_F | 0.000 | 0.000-0.211 | 0.056 | 0.000-0.439 | 0.000 | 0.000-0.288 | 0.478 | -0.93-1.040 | -0.089 | -0.898-0.852 | -0.167 | -0.636-0.755 |
| F_C | 0.077 | 0.000-0.613 | 0.000 | 0.000-0.272 | 0.000 | 0.000-0.293 | 0.365 | -1.033-1.111 | -0.364 | -2.740-0.378 | -0.367 | -1.769-0.236 |
| F_P | 0.037 | 0.000-0.350 | 0.074 | 0.000-0.458 | 0.000 | 0.000-0.249 | 0.545 | -0.977-1.061 | -0.365 | -0.993-0.606 | -0.459 | -1.04-0.242 |
| F_O | 0.067 | 0.000-0.405 | 0.000 | 0.000-0.413 | 0.000 | 0.000-0.246 | 0.237 | -1.037-1.118 | -0.343 | -1.367-0.413 | -0.412 | -0.959-0.27 |
| F_RF | 0.000 | 0.000-0.261 | 0.200 | 0.000-0.662 | 0.000 | 0.000-0.347 | 0.754 | -1.023-1.075 | 0.191 | -0.577-1.609 | -0.351 | -1.209-0.304 |
| F_RT | 0.439 | 0.204-0.773 | 0.107 | 0.000-0.791 | 0.000 | 0.000-0.504 | 0.468 | -0.984-1.055 | -0.545 | -1.863-0.135 | -1.729 | -3.047--1.253 |
| F_RC | 0.098 | 0.020-0.661 | 0.000 | 0.000-0.268 | 0.000 | 0.000-0.616 | 0.517 | -0.829-0.987 | -0.435 | -1.137-0.212 | -0.987 | -1.956--0.533 |
| F_RP | 0.021 | 0.000-0.297 | 0.000 | 0.000-0.336 | 0.000 | 0.000-0.343 | 0.329 | -0.99-1.026 | -0.337 | -0.891-0.857 | -0.370 | -0.926-0.334 |
| C_LF | 0.089 | 0.000-0.532 | 0.000 | 0.000-0.216 | 0.000 | 0.000-0.366 | 0.522 | -0.915-1.04 | -0.402 | -1.089-0.478 | -0.527 | -1.272-0.067 |
| C_LT | 0.000 | 0.000-0.251 | 0.070 | 0.000-0.429 | 0.105 | 0.000-0.512 | 0.186 | -1.057-1.059 | 0.499 | -0.181-2.321 | -0.519 | -1.035-0.189 |
| C_LC | 0.084 | 0.000-0.486 | 0.000 | 0.000-0.185 | 0.087 | 0.000-0.448 | -0.172 | -1.089-1.02 | 0.172 | -0.632-1.212 | -0.549 | -1.178-0.092 |
| C_LP | 0.000 | 0.000-0.242 | 0.000 | 0.000-0.160 | 0.000 | 0.000-0.186 | -0.447 | -0.842-1.006 | 0.259 | -0.631-1.334 | -0.310 | -0.94-0.37 |
| C_pF | 0.037 | 0.000-0.408 | 0.000 | 0.000-0.301 | 0.000 | 0.000-0.135 | 0.118 | -1.072-1.04 | -0.198 | -2.032-0.549 | -0.433 | -1.559-0.144 |
| C_F | 0.121 | 0.000-0.484 | 0.070 | 0.000-0.348 | 0.000 | 0.000-0.189 | 0.410 | -1.048-1.071 | -0.277 | -1.354-0.522 | -0.676 | -1.265--0.322 |
| C_C | 0.142 | 0.000-0.430 | 0.000 | 0.000-0.133 | 0.136 | 0.000-0.479 | -0.630 | -0.984-1.105 | 0.185 | -0.763-1.217 | -0.630 | -1.373--0.076 |
| C_P | 0.000 | 0.000-0.318 | 0.000 | 0.000-0.025 | 0.000 | 0.000-0.238 | -0.579 | -0.996-1.125 | 0.528 | -0.244-1.514 | 0.021 | -0.851-0.781 |
| C_O | 0.000 | 0.000-0.373 | 0.000 | 0.000-0.170 | 0.000 | 0.000-0.208 | -0.588 | -0.98-1.016 | 0.144 | -1.065-0.948 | -0.295 | -1.213-0.392 |
| C_RF | 0.111 | 0.000-0.527 | 0.018 | 0.000-0.319 | 0.000 | 0.000-0.171 | 0.085 | -1.059-1.072 | -0.110 | -1.131-0.733 | -0.849 | -1.548--0.435 |
| C_RT | 0.052 | 0.000-0.444 | 0.000 | 0.000-0.140 | 0.058 | 0.000-0.370 | -0.599 | -0.964-1.116 | 0.108 | -1.001-1.000 | -0.426 | -1.422-0.194 |
| C_RC | 0.245 | 0.000-0.511 | 0.000 | 0.000-0.178 | 0.122 | 0.000-0.486 | -0.574 | -1.058-1.166 | -0.071 | -0.837-1.678 | -0.729 | -1.42--0.302 |
| C_RP | 0.000 | 0.000-0.384 | 0.000 | 0.000-0.163 | 0.000 | 0.000-0.181 | -0.739 | -1.02-1.044 | 0.217 | -0.824-0.847 | -0.296 | -1.157-0.405 |
| P_LF | 0.201 | 0.062-0.477 | 0.000 | 0.000-0.269 | 0.000 | 0.000-0.152 | 0.475 | -0.903-1.087 | -0.648 | -1.487--0.385 | -0.556 | -1.584--0.446 |
| P_LT | 0.119 | 0.000-0.431 | 0.000 | 0.000-0.298 | 0.000 | 0.000-0.300 | -0.060 | -1.012-1.04 | -0.135 | -0.930-1.438 | -0.611 | -1.162--0.219 |
| P_LC | 0.323 | 0.058-0.595 | 0.073 | 0.000-0.707 | 0.000 | 0.000-0.456 | 0.872 | -1.039-1.09 | -1.018 | -3.139--0.371 | -0.646 | -1.235--0.39 |
| P_LP | 0.319 | 0.069-0.564 | 0.000 | 0.000-0.392 | 0.019 | 0.000-0.373 | -0.215 | -0.967-1.07 | -0.446 | -1.049-0.330 | -0.817 | -1.489--0.52 |
| P_pF | 0.467 | 0.078-0.784 | 0.001 | 0.000-0.464 | 0.000 | 0.000-0.273 | -0.309 | -1.039-1.157 | -0.753 | -5.460--0.030 | -1.081 | -2.146--0.574 |
| P_F | 0.300 | 0.043-0.571 | 0.052 | 0.000-0.546 | 0.000 | 0.000-0.390 | 0.625 | -1.019-1.016 | -0.955 | -3.226--0.728 | -0.643 | -1.235--0.387 |
| P_C | 0.234 | 0.000-0.514 | 0.000 | 0.000-0.228 | 0.000 | 0.000-0.378 | -0.260 | -1.029-1.107 | -0.301 | -0.899-0.659 | -0.830 | -1.512--0.478 |
| P_P | 0.033 | 0.000-0.381 | 0.000 | 0.000-0.193 | 0.000 | 0.000-0.240 | -0.275 | -0.973-1.053 | -0.444 | -1.241-0.335 | -0.271 | -1.154-0.392 |
| P_O | 0.167 | 0.000-0.466 | 0.000 | 0.000-0.228 | 0.000 | 0.000-0.238 | -0.172 | -0.984-0.991 | -0.595 | -1.650-0.054 | -0.482 | -1.617-0.027 |
| P_RF | 0.320 | 0.013-0.625 | 0.000 | 0.000-0.300 | 0.000 | 0.000-0.176 | -0.270 | -1.053-1.139 | -0.557 | -2.345-0.134 | -0.887 | -1.762--0.412 |
| P_RT | 0.555 | 0.222-0.777 | 0.000 | 0.000-0.205 | 0.003 | 0.000-0.418 | -0.123 | -0.979-1.085 | -0.749 | -1.728--0.175 | -1.501 | -3.077--0.903 |
| P_RC | 0.461 | 0.176-0.709 | 0.000 | 0.000-0.365 | 0.000 | 0.000-0.348 | -0.279 | -0.952-1.116 | -0.710 | -1.950--0.126 | -1.157 | -2.304--0.745 |
| P_RP | 0.123 | 0.000-0.533 | 0.000 | 0.000-0.287 | 0.000 | 0.000-0.271 | -0.199 | -1.04-1.087 | -0.408 | -3.149-0.268 | -0.539 | -1.224-0.067 |
| O_LF | 0.111 | 0.029-0.449 | 0.000 | 0.000-0.461 | 0.000 | 0.000-0.194 | 0.389 | -0.894-1.027 | -0.449 | -1.076-0.235 | -0.602 | -2.095--0.492 |
| O_LT | 0.000 | 0.000-0.374 | 0.000 | 0.000-0.107 | 0.000 | 0.000-0.171 | -0.047 | -0.928-1.062 | 0.181 | -1.362-0.733 | -0.443 | -1.155-0.148 |
| O_LC | 0.150 | 0.000-0.599 | 0.000 | 0.000-0.361 | 0.000 | 0.000-0.121 | 0.095 | -1.012-1.08 | -0.254 | -2.702-0.445 | -0.745 | -1.537--0.529 |
| O_LP | 0.001 | 0.000-0.306 | 0.000 | 0.000-0.200 | 0.000 | 0.000-0.339 | -0.659 | -0.944-1.04 | 0.183 | -0.898-0.928 | -0.342 | -0.972-0.317 |
| O_pF | 0.224 | 0.018-0.522 | 0.000 | 0.000-0.438 | 0.000 | 0.000-0.372 | -0.161 | -1.039-1.05 | -0.309 | -1.143-0.485 | -0.694 | -1.752--0.463 |
| O_F | 0.166 | 0.000-0.476 | 0.000 | 0.000-0.209 | 0.000 | 0.000-0.086 | 0.136 | -0.912-1.044 | -0.463 | -1.680-0.240 | -0.558 | -1.17--0.246 |
| O_C | 0.328 | 0.100-0.567 | 0.000 | 0.000-0.298 | 0.000 | 0.000-0.162 | -0.280 | -1.173-1.085 | -0.705 | -1.666--0.196 | -0.858 | -1.521--0.542 |
| O_P | 0.000 | 0.000-0.422 | 0.000 | 0.000-0.148 | 0.000 | 0.000-0.170 | -0.083 | -0.98-1.053 | -0.761 | -1.960--0.247 | -0.109 | -0.999-0.518 |
| O_O | 0.123 | 0.000-0.545 | 0.000 | 0.000-0.285 | 0.000 | 0.000-0.206 | -0.339 | -1.097-1.077 | -0.306 | -1.512-0.457 | -0.719 | -1.977--0.13 |
| O_RF | 0.021 | 0.000-0.352 | 0.000 | 0.000-0.300 | 0.000 | 0.000-0.313 | -0.151 | -1.005-1.056 | 0.075 | -1.063-0.928 | -0.502 | -1.148--0.046 |
| O_RT | 0.429 | 0.196-0.697 | 0.000 | 0.000-0.244 | 0.094 | 0.000-0.440 | -0.275 | -0.987-1.091 | -0.398 | -1.092-0.423 | -1.240 | -2.395--0.828 |
| O_RC | 0.441 | 0.211-0.756 | 0.000 | 0.000-0.375 | 0.001 | 0.000-0.282 | -0.251 | -1.067-1.022 | -0.686 | -2.437--0.001 | -1.071 | -2.373--0.738 |
| O_RP | 0.000 | 0.000-0.353 | 0.000 | 0.000-0.257 | 0.000 | 0.000-0.152 | -0.449 | -1.011-1.023 | -0.192 | -0.811-0.934 | 0.015 | -1.088-0.594 |
| RF_LF | 0.008 | 0.000-0.451 | 0.000 | 0.000-0.440 | 0.000 | 0.000-0.559 | 0.064 | -1.101-1.032 | -0.411 | -2.146-0.270 | -0.299 | -1.158-0.273 |
| RF_LT | 0.000 | 0.000-0.256 | 0.000 | 0.000-0.362 | 0.000 | 0.000-0.494 | 0.616 | -1.018-1.033 | 0.266 | -0.836-0.846 | 0.029 | -1.134-0.481 |
| RF_LC | 0.000 | 0.000-0.499 | 0.000 | 0.000-0.251 | 0.042 | 0.000-0.573 | 0.474 | -1.005-1.127 | -0.481 | -1.424-0.347 | -0.257 | -1.413-0.385 |
| RF_LP | 0.015 | 0.000-0.346 | 0.065 | 0.000-0.631 | 0.013 | 0.000-0.251 | 0.311 | -1.022-1.203 | 0.246 | -0.735-0.916 | -0.404 | -1.065-0.452 |
| RF_pF | 0.158 | 0.000-0.447 | 0.000 | 0.000-0.438 | 0.000 | 0.000-0.287 | 0.046 | -1.026-1.201 | -0.278 | -1.121-0.674 | -0.700 | -1.559--0.504 |
| RF_F | 0.129 | 0.000-0.523 | 0.067 | 0.000-0.480 | 0.012 | 0.000-0.292 | 1.200 | -1.026-1.14 | -0.887 | -2.065--0.390 | -0.281 | -1.256-0.374 |
| RF_C | 0.000 | 0.000-0.268 | 0.026 | 0.000-0.427 | 0.000 | 0.000-0.178 | 0.158 | -0.991-1.028 | 0.145 | -0.831-1.109 | -0.531 | -1.419--0.389 |
| RF_P | 0.005 | 0.000-0.479 | 0.156 | 0.000-0.575 | 0.000 | 0.000-0.175 | 1.438 | -1.037-1.112 | -0.906 | -2.621--0.306 | -0.038 | -0.976-0.473 |
| RF_O | 0.046 | 0.000-0.551 | 0.227 | 0.014-0.691 | 0.000 | 0.000-0.247 | 1.470 | -1.038-1.142 | -0.320 | -1.871-0.403 | -0.858 | -2.132--0.238 |
| RF_RF | 0.000 | 0.000-0.231 | 0.103 | 0.000-0.519 | 0.099 | 0.000-0.475 | 0.489 | -0.958-1.067 | -0.500 | -1.157-0.186 | -0.153 | -0.688-0.692 |
| RF_RT | 0.588 | 0.361-0.774 | 0.000 | 0.000-0.419 | 0.000 | 0.000-0.351 | 0.026 | -1.178-1.069 | -1.234 | -2.391--0.893 | -1.243 | -2.176--0.905 |
| RF_RC | 0.382 | 0.128-0.627 | 0.000 | 0.000-0.405 | 0.016 | 0.000-0.403 | 0.155 | -1.073-1.110 | -0.749 | -1.888--0.489 | -0.773 | -1.553--0.523 |
| RF_RP | 0.000 | 0.000-0.637 | 0.000 | 0.000-0.316 | 0.000 | 0.000-0.283 | 0.636 | -0.986-1.04 | 0.043 | -5.577-0.595 | -0.300 | -1.667-0.298 |
| RT_LF | 0.018 | 0.000-0.451 | 0.000 | 0.000-0.661 | 0.000 | 0.000-0.354 | 0.802 | -0.891-1.085 | -0.285 | -1.819-0.409 | -0.464 | -1.449-0.21 |
| RT_LT | 0.000 | 0.000-0.292 | 0.000 | 0.000-0.185 | 0.000 | 0.000-0.281 | 0.439 | -1.164-1.104 | -0.053 | -0.883-0.860 | -0.339 | -1.211-0.351 |
| RT_LC | 0.000 | 0.000-0.510 | 0.001 | 0.000-0.306 | 0.000 | 0.000-0.208 | 1.115 | -1.083-1.064 | -0.143 | -2.232-0.479 | -0.542 | -1.434-0.073 |
| RT_LP | 0.094 | 0.000-0.333 | 0.000 | 0.000-0.309 | 0.000 | 0.000-0.146 | 0.269 | -0.971-1.142 | -0.461 | -1.726-0.298 | -0.429 | -1.069-0.229 |
| RT_pF | 0.000 | 0.000-0.457 | 0.000 | 0.000-0.331 | 0.000 | 0.000-0.137 | 0.398 | -1.032-1.167 | 0.211 | -1.341-0.999 | -0.798 | -1.623--0.251 |
| RT_F | 0.212 | 0.039-0.484 | 0.000 | 0.000-0.722 | 0.000 | 0.000-0.358 | 0.495 | -1.03-1.070 | -0.798 | -1.978--0.417 | -0.548 | -0.992--0.202 |
| RT_C | 0.000 | 0.000-0.299 | 0.072 | 0.000-0.526 | 0.000 | 0.000-0.460 | 1.106 | -1.068-1.082 | 0.194 | -1.595-0.831 | -0.249 | -0.897-0.449 |
| RT_P | 0.077 | 0.000-0.593 | 0.162 | 0.000-0.503 | 0.000 | 0.000-0.242 | 0.797 | -0.899-1.046 | -0.530 | -1.821-0.214 | -0.397 | -1.632-0.206 |
| RT_O | 0.103 | 0.000-0.381 | 0.000 | 0.000-0.373 | 0.000 | 0.000-0.153 | 0.124 | -0.917-1.027 | -0.621 | -1.794-0.069 | -0.385 | -0.891-0.327 |
| RT_RF | 0.000 | 0.000-0.325 | 0.020 | 0.000-0.417 | 0.000 | 0.000-0.423 | 0.364 | -1.084-1.104 | 0.240 | -0.850-0.976 | -0.618 | -1.116--0.167 |
| RT_RT | 0.000 | 0.000-0.272 | 0.000 | 0.000-0.197 | 0.000 | 0.000-0.161 | 0.151 | -0.985-1.101 | 0.193 | -1.632-0.745 | 0.150 | -0.893-0.601 |
| RT_RC | 0.009 | 0.000-0.510 | 0.147 | 0.000-0.710 | 0.000 | 0.000-0.259 | 0.769 | -1.046-1.127 | -0.260 | -1.952-0.389 | -0.278 | -1.411-0.331 |
| RT_RP | 0.000 | 0.000-0.454 | 0.049 | 0.000-0.441 | 0.000 | 0.000-0.423 | 0.694 | -0.882-0.966 | -0.199 | -1.547-0.574 | -0.444 | -1.543-0.194 |
| RC_LF | 0.124 | 0.000-0.502 | 0.000 | 0.000-0.200 | 0.000 | 0.000-0.371 | 0.519 | -0.924-1.085 | -0.424 | -1.273-0.352 | -0.557 | -1.615-0.042 |
| RC_LT | 0.000 | 0.000-0.275 | 0.000 | 0.000-0.305 | 0.000 | 0.000-0.397 | 0.530 | -0.972-1.029 | -0.003 | -0.771-1.376 | -0.434 | -1.212-0.196 |
| RC_LC | 0.119 | 0.000-0.533 | 0.000 | 0.000-0.207 | 0.000 | 0.000-0.374 | 0.125 | -1.081-1.166 | -0.111 | -1.359-0.692 | -0.651 | -1.293--0.199 |
| RC_LP | 0.000 | 0.000-0.474 | 0.026 | 0.000-0.304 | 0.020 | 0.000-0.320 | 0.060 | -1.004-1.163 | 0.145 | -0.923-0.934 | -0.912 | -1.761--0.427 |
| RC_pF | 0.136 | 0.000-0.495 | 0.000 | 0.000-0.348 | 0.000 | 0.000-0.173 | 0.097 | -1.127-1.065 | -0.084 | -0.988-0.834 | -0.894 | -1.861--0.617 |
| RC_F | 0.211 | 0.000-0.533 | 0.240 | 0.000-0.715 | 0.000 | 0.000-0.366 | 0.600 | -0.968-1.122 | -0.194 | -0.891-0.918 | -0.904 | -1.792--0.618 |
| RC_C | 0.000 | 0.000-0.260 | 0.005 | 0.000-0.256 | 0.065 | 0.000-0.432 | -0.322 | -1.084-1.059 | 0.526 | -0.179-1.431 | -0.573 | -1.484-0.094 |
| RC_P | 0.000 | 0.000-0.560 | 0.037 | 0.000-0.362 | 0.000 | 0.000-0.316 | 0.239 | -1.01-1.034 | 0.013 | -0.870-0.761 | -0.261 | -3.115-0.315 |
| RC_O | 0.051 | 0.000-0.400 | 0.112 | 0.000-0.470 | 0.000 | 0.000-0.141 | 0.383 | -1.113-1.079 | -0.249 | -1.476-0.572 | -0.471 | -1.162-0.194 |
| RC_RF | 0.000 | 0.000-0.308 | 0.173 | 0.004-0.485 | 0.000 | 0.000-0.322 | 0.638 | -1.014-1.148 | 0.206 | -0.528-1.344 | -0.932 | -1.695--0.497 |
| RC_RT | 0.000 | 0.000-0.258 | 0.183 | 0.000-0.636 | 0.000 | 0.000-0.203 | 0.732 | -1.094-1.066 | 0.298 | -1.027-0.976 | -0.249 | -1.068-0.517 |
| RC_RC | 0.000 | 0.000-0.321 | 0.272 | 0.022-0.767 | 0.000 | 0.000-0.511 | 0.746 | -1.043-1.059 | 0.223 | -0.502-1.247 | -0.586 | -1.722-0.059 |
| RC_RP | 0.000 | 0.000-0.352 | 0.080 | 0.000-0.420 | 0.000 | 0.000-0.359 | 0.463 | -0.875-0.885 | -0.217 | -0.903-0.638 | -0.422 | -2.999-0.217 |
| RP_LF | 0.019 | 0.000-0.433 | 0.000 | 0.000-0.438 | 0.000 | 0.000-0.188 | 0.643 | -0.872-0.962 | -0.428 | -1.329-0.073 | -0.564 | -1.132--0.03 |
| RP_LT | 0.127 | 0.000-0.477 | 0.000 | 0.000-0.318 | 0.133 | 0.000-0.489 | -0.061 | -0.974-1.121 | 0.112 | -0.556-1.475 | -0.685 | -1.610--0.079 |
| RP_LC | 0.341 | 0.060-0.647 | 0.000 | 0.000-0.538 | 0.000 | 0.000-0.414 | 0.691 | -1.052-1.093 | -0.743 | -2.002--0.450 | -0.733 | -1.398--0.256 |
| RP_LP | 0.000 | 0.000-0.594 | 0.000 | 0.000-0.140 | 0.019 | 0.000-0.529 | -0.823 | -1.044-1.136 | 0.372 | -0.559-0.996 | -1.261 | -3.732--0.604 |
| RP_pF | 0.305 | 0.022-0.585 | 0.000 | 0.000-0.398 | 0.013 | 0.000-0.502 | 0.035 | -1.021-1.118 | -0.465 | -1.241-0.236 | -0.830 | -1.375--0.456 |
| RP_F | 0.079 | 0.000-0.538 | 0.000 | 0.000-0.169 | 0.000 | 0.000-0.286 | 0.614 | -0.813-0.997 | -0.497 | -3.567--0.104 | -0.554 | -1.286-0.067 |
| RP_C | 0.298 | 0.000-0.660 | 0.000 | 0.000-0.225 | 0.144 | 0.000-0.492 | -0.291 | -1.066-1.066 | -0.078 | -13.701-0.733 | -0.904 | -1.890--0.428 |
| RP_P | 0.060 | 0.000-0.655 | 0.000 | 0.000-0.250 | 0.000 | 0.000-0.196 | 0.393 | -1.175-1.068 | -0.449 | -2.137-0.335 | -0.277 | -2.122-0.262 |
| RP_O | 0.292 | 0.000-0.637 | 0.000 | 0.000-0.331 | 0.066 | 0.000-0.507 | -0.480 | -1.200-1.037 | -0.215 | -1.161-0.968 | -0.962 | -2.007--0.397 |
| RP_RF | 0.150 | 0.000-0.475 | 0.000 | 0.000-0.125 | 0.000 | 0.000-0.351 | 0.194 | -0.902-1.006 | -0.339 | -1.121-0.484 | -0.775 | -1.739--0.264 |
| RP_RT | 0.188 | 0.034-0.555 | 0.000 | 0.000-0.285 | 0.000 | 0.000-0.490 | 0.587 | -0.951-0.972 | -0.568 | -1.119-0.052 | -0.798 | -1.535--0.342 |
| RP_RC | 0.267 | 0.068-0.632 | 0.000 | 0.000-0.256 | 0.000 | 0.000-0.543 | 0.411 | -0.941-1.056 | -0.518 | -1.105-0.166 | -0.912 | -1.857--0.398 |
| RP_RP | 0.054 | 0.000-0.374 | 0.005 | 0.000-0.460 | 0.000 | 0.000-0.448 | -0.001 | -1.01-1.084 | -0.315 | -1.062-0.592 | -0.291 | -1.044-0.448 |

Note. ROI refers to directional pairs of brain regions analyzed for gamma-band connectivity using spectral Granger causality

Supplementary Table 6A. Predictive performance of resting-state gamma power for CF increase based on leave-one-out cross-validation

| **Electrode** | **OR** | **95% CI for OR** | **p vale** | **q value** | **AUC** | **95% CI for AUC** | **Cut**  **off** | **Accuracy** | **Sensitivity** | **Specificity** |
| --- | --- | --- | --- | --- | --- | --- | --- | --- | --- | --- |
| Fp1 | 2.059 | 0.661 - 6.413 | 0.213 | 0.453 | 0.524 | 0.197 - 0.872 | 0.397 | 0.625 | 0.714 | 0.556 |
| Fz | 1.914 | 0.636 - 5.760 | 0.248 | 0.453 | 0.603 | 0.282 - 0.937 | 0.489 | 0.750 | 0.571 | 0.889 |
| F3 | 2.161 | 0.677 - 6.902 | 0.193 | 0.453 | 0.619 | 0.276 - 0.925 | 0.509 | 0.750 | 0.571 | 0.889 |
| F7 | 1.381 | 0.483 - 3.949 | 0.547 | 0.565 | 0.270 | 0.033 - 0.569 | 0.419 | 0.500 | 0.714 | 0.333 |
| FT9 | 2.109 | 0.684 - 6.505 | 0.194 | 0.453 | 0.556 | 0.231 - 0.889 | 0.417 | 0.688 | 0.714 | 0.667 |
| FC5 | 1.249 | 0.443 - 3.520 | 0.674 | 0.674 | 0.000 | 0.000 - 0.000 | 0.000 | 0.438 | 1.000 | 0.000 |
| FC1 | 1.578 | 0.538 - 4.627 | 0.405 | 0.466 | 0.492 | 0.175 - 0.837 | 0.434 | 0.625 | 0.571 | 0.667 |
| C3 | 1.736 | 0.581 - 5.191 | 0.323 | 0.453 | 0.540 | 0.218 - 0.869 | 0.405 | 0.688 | 0.714 | 0.667 |
| T7 | 1.360 | 0.478 - 3.869 | 0.564 | 0.573 | 0.270 | 0.063 - 0.608 | 0.000 | 0.438 | 1.000 | 0.000 |
| TP9 | 3.707 | 1.015 - 13.542 | 0.047 | 0.453 | 0.714 | 0.364 - 1.000 | 0.516 | 0.813 | 0.714 | 0.889 |
| CP5 | 3.488 | 0.905 - 13.443 | 0.070 | 0.453 | 0.683 | 0.371 - 1.000 | 0.473 | 0.813 | 0.714 | 0.889 |
| CP1 | 1.541 | 0.537 - 4.423 | 0.422 | 0.466 | 0.333 | 0.067 - 0.650 | 0.412 | 0.500 | 0.571 | 0.444 |
| Pz | 1.745 | 0.590 - 5.157 | 0.314 | 0.453 | 0.460 | 0.126 - 0.833 | 0.473 | 0.688 | 0.429 | 0.889 |
| P3 | 2.267 | 0.711 - 7.223 | 0.166 | 0.453 | 0.556 | 0.213 - 0.882 | 0.418 | 0.688 | 0.714 | 0.667 |
| P7 | 2.306 | 0.709 - 7.499 | 0.165 | 0.453 | 0.619 | 0.234 - 0.967 | 0.442 | 0.750 | 0.714 | 0.778 |
| O1 | 2.213 | 0.701 - 6.986 | 0.176 | 0.453 | 0.587 | 0.254 - 0.931 | 0.417 | 0.750 | 0.714 | 0.778 |
| Oz | 2.162 | 0.700 - 6.675 | 0.180 | 0.453 | 0.619 | 0.267 - 0.974 | 0.393 | 0.750 | 0.714 | 0.778 |
| O2 | 1.815 | 0.614 - 5.372 | 0.281 | 0.453 | 0.476 | 0.156 - 0.813 | 0.399 | 0.625 | 0.571 | 0.667 |
| P4 | 1.715 | 0.584 - 5.038 | 0.327 | 0.453 | 0.460 | 0.142 - 0.808 | 0.429 | 0.688 | 0.571 | 0.778 |
| P8 | 1.744 | 0.591 - 5.144 | 0.314 | 0.453 | 0.476 | 0.154 - 0.818 | 0.429 | 0.688 | 0.571 | 0.778 |
| TP10 | 1.912 | 0.629 - 5.815 | 0.253 | 0.453 | 0.571 | 0.236 - 0.905 | 0.418 | 0.688 | 0.714 | 0.667 |
| CP6 | 2.207 | 0.688 - 7.085 | 0.183 | 0.453 | 0.571 | 0.250 - 0.917 | 0.419 | 0.750 | 0.714 | 0.778 |
| CP2 | 1.775 | 0.609 - 5.176 | 0.293 | 0.453 | 0.492 | 0.144 - 0.858 | 0.468 | 0.688 | 0.571 | 0.778 |
| Cz | 1.398 | 0.496 - 3.943 | 0.526 | 0.553 | 0.333 | 0.092 - 0.656 | 0.438 | 0.563 | 0.571 | 0.556 |
| C4 | 2.321 | 0.733 - 7.349 | 0.152 | 0.453 | 0.587 | 0.250 - 0.927 | 0.403 | 0.688 | 0.714 | 0.667 |
| T8 | 1.567 | 0.534 - 4.598 | 0.414 | 0.466 | 0.444 | 0.148 - 0.764 | 0.466 | 0.625 | 0.429 | 0.778 |
| FT10 | 1.651 | 0.561 - 4.861 | 0.363 | 0.466 | 0.460 | 0.167 - 0.780 | 0.472 | 0.625 | 0.429 | 0.778 |
| FC6 | 4.132 | 0.935 - 18.271 | 0.061 | 0.453 | 0.651 | 0.305 - 0.949 | 0.579 | 0.750 | 0.571 | 0.889 |
| FC2 | 2.103 | 0.671 - 6.589 | 0.202 | 0.453 | 0.492 | 0.189 - 0.794 | 0.534 | 0.625 | 0.429 | 0.778 |
| F4 | 1.900 | 0.646 - 5.587 | 0.243 | 0.453 | 0.540 | 0.222 - 0.874 | 0.374 | 0.688 | 0.714 | 0.667 |
| F8 | 3.063 | 0.853 - 10.994 | 0.086 | 0.453 | 0.651 | 0.291 - 0.952 | 0.701 | 0.750 | 0.429 | 1.000 |
| Fp2 | 2.293 | 0.698 - 7.536 | 0.172 | 0.453 | 0.556 | 0.236 - 0.893 | 0.589 | 0.688 | 0.286 | 1.000 |
| AF7 | 1.436 | 0.503 - 4.101 | 0.499 | 0.533 | 0.286 | 0.048 - 0.600 | 0.418 | 0.500 | 0.571 | 0.444 |
| AF3 | 1.942 | 0.639 - 5.902 | 0.242 | 0.453 | 0.524 | 0.218 - 0.845 | 0.452 | 0.688 | 0.571 | 0.778 |
| AFz | 1.786 | 0.594 - 5.370 | 0.302 | 0.453 | 0.524 | 0.200 - 0.855 | 0.471 | 0.688 | 0.571 | 0.778 |
| F1 | 1.885 | 0.618 - 5.751 | 0.265 | 0.453 | 0.524 | 0.190 - 0.869 | 0.403 | 0.688 | 0.714 | 0.667 |
| F5 | 1.726 | 0.574 - 5.192 | 0.331 | 0.453 | 0.524 | 0.206 - 0.857 | 0.408 | 0.625 | 0.714 | 0.556 |
| FT7 | 1.596 | 0.544 - 4.682 | 0.395 | 0.466 | 0.349 | 0.089 - 0.661 | 0.383 | 0.563 | 0.714 | 0.444 |
| FC3 | 2.509 | 0.651 - 9.674 | 0.181 | 0.453 | 0.651 | 0.300 - 1.000 | 0.458 | 0.813 | 0.714 | 0.889 |
| C1 | 1.463 | 0.514 - 4.166 | 0.476 | 0.517 | 0.302 | 0.000 - 0.639 | 0.435 | 0.563 | 0.429 | 0.667 |
| C5 | 1.982 | 0.639 - 6.151 | 0.236 | 0.453 | 0.603 | 0.286 - 0.905 | 0.408 | 0.688 | 0.714 | 0.667 |
| TP7 | 3.298 | 0.928 - 11.725 | 0.065 | 0.453 | 0.667 | 0.333 - 0.953 | 0.439 | 0.750 | 0.714 | 0.778 |
| CP3 | 1.839 | 0.614 - 5.508 | 0.276 | 0.453 | 0.444 | 0.144 - 0.800 | 0.549 | 0.625 | 0.286 | 0.889 |
| P1 | 1.946 | 0.636 - 5.950 | 0.243 | 0.453 | 0.492 | 0.167 - 0.857 | 0.456 | 0.688 | 0.571 | 0.778 |
| P5 | 2.129 | 0.671 - 6.750 | 0.199 | 0.453 | 0.587 | 0.252 - 0.917 | 0.475 | 0.750 | 0.714 | 0.778 |
| PO7 | 2.265 | 0.716 - 7.161 | 0.164 | 0.453 | 0.571 | 0.234 - 0.913 | 0.432 | 0.750 | 0.714 | 0.778 |
| PO3 | 2.286 | 0.728 - 7.174 | 0.156 | 0.453 | 0.603 | 0.254 - 0.922 | 0.438 | 0.750 | 0.714 | 0.778 |
| POz | 2.021 | 0.665 - 6.145 | 0.215 | 0.453 | 0.619 | 0.270 - 0.967 | 0.439 | 0.750 | 0.714 | 0.778 |
| PO4 | 1.928 | 0.639 - 5.822 | 0.244 | 0.453 | 0.556 | 0.215 - 0.883 | 0.444 | 0.688 | 0.571 | 0.778 |
| PO8 | 1.621 | 0.555 - 4.737 | 0.377 | 0.466 | 0.460 | 0.133 - 0.809 | 0.435 | 0.625 | 0.571 | 0.667 |
| P6 | 1.672 | 0.572 - 4.885 | 0.347 | 0.465 | 0.460 | 0.133 - 0.835 | 0.439 | 0.688 | 0.571 | 0.778 |
| P2 | 1.651 | 0.564 - 4.833 | 0.361 | 0.466 | 0.460 | 0.159 - 0.795 | 0.429 | 0.625 | 0.571 | 0.667 |
| CPz | 1.552 | 0.542 - 4.442 | 0.413 | 0.466 | 0.349 | 0.100 - 0.696 | 0.424 | 0.563 | 0.429 | 0.667 |
| CP4 | 1.728 | 0.582 - 5.134 | 0.325 | 0.453 | 0.476 | 0.171 - 0.817 | 0.404 | 0.625 | 0.714 | 0.556 |
| TP8 | 1.896 | 0.624 - 5.760 | 0.259 | 0.453 | 0.571 | 0.244 - 0.909 | 0.431 | 0.688 | 0.714 | 0.667 |
| C6 | 1.620 | 0.551 - 4.766 | 0.381 | 0.466 | 0.397 | 0.127 - 0.762 | 0.413 | 0.625 | 0.714 | 0.556 |
| C2 | 1.804 | 0.613 - 5.310 | 0.284 | 0.453 | 0.444 | 0.144 - 0.783 | 0.453 | 0.625 | 0.571 | 0.667 |
| FC4 | 4.603 | 1.102 - 19.226 | 0.036 | 0.453 | 0.667 | 0.315 - 0.953 | 0.302 | 0.750 | 0.714 | 0.778 |
| FT8 | 1.599 | 0.552 - 4.634 | 0.387 | 0.466 | 0.429 | 0.133 - 0.792 | 0.426 | 0.563 | 0.571 | 0.556 |
| F6 | 2.393 | 0.754 - 7.601 | 0.139 | 0.453 | 0.603 | 0.254 - 0.938 | 0.496 | 0.750 | 0.571 | 0.889 |
| AF8 | 2.603 | 0.739 - 9.170 | 0.137 | 0.453 | 0.619 | 0.266 - 0.927 | 0.390 | 0.688 | 0.714 | 0.667 |
| AF4 | 2.646 | 0.717 - 9.769 | 0.144 | 0.453 | 0.651 | 0.250 - 1.000 | 0.563 | 0.813 | 0.571 | 1.000 |
| F2 | 3.179 | 0.874 - 11.560 | 0.079 | 0.453 | 0.635 | 0.283 - 0.967 | 0.360 | 0.750 | 0.714 | 0.778 |

OR, odds ratio; CI, confidence interval; AUC, area under the curve; Cut-off, optimal threshold for classification; q value, false discovery rate–adjusted p value using the Benjamini–Hochberg procedure.

Note. Electrode refers to individual scalp EEG electrodes used in the classification analysis of baseline resting-state gamma power to predict CF increase.

Supplementary Table 6B. Predictive performance of gamma-band event-related synchronization for CF increase based on leave-one-out cross-validation

| **Electrode** | **OR** | **95% CI for OR** | **p vale** | **q value** | **AUC** | **95% CI for AUC** | **Cut**  **off** | **Accuracy** | **Sensitivity** | **Specificity** |
| --- | --- | --- | --- | --- | --- | --- | --- | --- | --- | --- |
| Fp1 | 1.782 | 0.596 - 5.324 | 0.301 | 0.533 | 0.397 | 0.087 - 0.763 | 0.485 | 0.625 | 0.429 | 0.778 |
| Fz | 2.591 | 0.672 - 9.992 | 0.167 | 0.533 | 0.540 | 0.207 - 0.854 | 0.308 | 0.625 | 0.857 | 0.444 |
| F3 | 2.374 | 0.626 - 8.997 | 0.204 | 0.533 | 0.556 | 0.233 - 0.867 | 0.412 | 0.688 | 0.714 | 0.667 |
| F7 | 1.935 | 0.559 - 6.691 | 0.297 | 0.533 | 0.413 | 0.127 - 0.724 | 0.473 | 0.563 | 0.286 | 0.778 |
| FT9 | 4.449 | 0.757 - 26.151 | 0.099 | 0.533 | 0.714 | 0.381 - 0.974 | 0.457 | 0.813 | 0.714 | 0.889 |
| FC5 | 1.954 | 0.602 - 6.345 | 0.265 | 0.533 | 0.444 | 0.167 - 0.785 | 0.416 | 0.563 | 0.571 | 0.556 |
| FC1 | 1.512 | 0.510 - 4.484 | 0.456 | 0.558 | 0.143 | 0.000 - 0.386 | 0.000 | 0.438 | 1.000 | 0.000 |
| C3 | 1.636 | 0.556 - 4.815 | 0.372 | 0.536 | 0.349 | 0.094 - 0.673 | 0.426 | 0.563 | 0.429 | 0.667 |
| T7 | 3.516 | 0.870 - 14.205 | 0.078 | 0.533 | 0.698 | 0.381 - 0.983 | 0.322 | 0.750 | 0.857 | 0.667 |
| TP9 | 1.917 | 0.636 - 5.782 | 0.248 | 0.533 | 0.381 | 0.065 - 0.748 | 0.453 | 0.625 | 0.429 | 0.778 |
| CP5 | 0.999 | 0.360 - 2.770 | 0.998 | 0.998 | 0.000 | 0.000 - 0.000 | 0.000 | 0.438 | 1.000 | 0.000 |
| CP1 | 1.504 | 0.516 - 4.381 | 0.454 | 0.558 | 0.349 | 0.089 - 0.651 | 0.377 | 0.563 | 0.857 | 0.333 |
| Pz | 2.233 | 0.656 - 7.604 | 0.199 | 0.533 | 0.571 | 0.226 - 0.874 | 0.548 | 0.688 | 0.429 | 0.889 |
| P3 | 0.943 | 0.340 - 2.615 | 0.910 | 0.939 | 0.000 | 0.000 - 0.000 | 0.000 | 0.438 | 1.000 | 0.000 |
| P7 | 1.233 | 0.440 - 3.457 | 0.690 | 0.776 | 0.000 | 0.000 - 0.000 | 0.000 | 0.438 | 1.000 | 0.000 |
| O1 | 0.623 | 0.215 - 1.805 | 0.383 | 0.536 | 0.365 | 0.117 - 0.683 | 0.424 | 0.563 | 0.571 | 0.556 |
| Oz | 1.698 | 0.564 - 5.114 | 0.347 | 0.533 | 0.429 | 0.145 - 0.762 | 0.396 | 0.563 | 0.714 | 0.444 |
| O2 | 1.573 | 0.534 - 4.633 | 0.412 | 0.548 | 0.397 | 0.125 - 0.695 | 0.401 | 0.500 | 0.714 | 0.333 |
| P4 | 1.191 | 0.427 - 3.320 | 0.738 | 0.805 | 0.000 | 0.000 - 0.000 | 0.000 | 0.438 | 1.000 | 0.000 |
| P8 | 2.335 | 0.672 - 8.109 | 0.182 | 0.533 | 0.492 | 0.189 - 0.795 | 0.288 | 0.563 | 0.857 | 0.333 |
| TP10 | 1.727 | 0.567 - 5.262 | 0.336 | 0.533 | 0.365 | 0.095 - 0.661 | 0.408 | 0.500 | 0.571 | 0.444 |
| CP6 | 1.700 | 0.571 - 5.064 | 0.341 | 0.533 | 0.476 | 0.185 - 0.789 | 0.394 | 0.625 | 0.714 | 0.556 |
| CP2 | 1.851 | 0.600 - 5.712 | 0.284 | 0.533 | 0.508 | 0.183 - 0.825 | 0.402 | 0.625 | 0.714 | 0.556 |
| Cz | 2.463 | 0.687 - 8.831 | 0.167 | 0.533 | 0.540 | 0.230 - 0.827 | 0.204 | 0.563 | 1.000 | 0.222 |
| C4 | 1.691 | 0.527 - 5.431 | 0.377 | 0.536 | 0.254 | 0.035 - 0.517 | 0.000 | 0.438 | 1.000 | 0.000 |
| T8 | 4.070 | 0.959 - 17.271 | 0.057 | 0.533 | 0.714 | 0.398 - 0.968 | 0.301 | 0.750 | 0.857 | 0.667 |
| FT10 | 3.282 | 0.881 - 12.226 | 0.077 | 0.533 | 0.683 | 0.384 - 0.938 | 0.579 | 0.750 | 0.571 | 0.889 |
| FC6 | 2.759 | 0.802 - 9.493 | 0.107 | 0.533 | 0.587 | 0.267 - 0.874 | 0.472 | 0.688 | 0.571 | 0.778 |
| FC2 | 1.927 | 0.601 - 6.179 | 0.270 | 0.533 | 0.444 | 0.095 - 0.800 | 0.553 | 0.688 | 0.286 | 1.000 |
| F4 | 1.589 | 0.542 - 4.655 | 0.399 | 0.546 | 0.333 | 0.092 - 0.667 | 0.455 | 0.563 | 0.571 | 0.556 |
| F8 | 3.048 | 0.783 - 11.861 | 0.108 | 0.533 | 0.651 | 0.333 - 0.935 | 0.611 | 0.750 | 0.429 | 1.000 |
| Fp2 | 1.923 | 0.612 - 6.050 | 0.263 | 0.533 | 0.571 | 0.244 - 0.908 | 0.533 | 0.688 | 0.286 | 1.000 |
| AF7 | 3.166 | 0.786 - 12.750 | 0.105 | 0.533 | 0.619 | 0.291 - 0.933 | 0.413 | 0.688 | 0.714 | 0.667 |
| AF3 | 2.911 | 0.730 - 11.603 | 0.130 | 0.533 | 0.603 | 0.254 - 0.891 | 0.428 | 0.688 | 0.714 | 0.667 |
| AFz | 3.738 | 0.808 - 17.302 | 0.092 | 0.533 | 0.571 | 0.236 - 0.867 | 0.660 | 0.688 | 0.286 | 1.000 |
| F1 | 3.059 | 0.554 - 16.886 | 0.200 | 0.533 | 0.524 | 0.182 - 0.873 | 0.501 | 0.688 | 0.571 | 0.778 |
| F5 | 2.646 | 0.596 - 11.756 | 0.201 | 0.533 | 0.492 | 0.156 - 0.833 | 0.388 | 0.625 | 0.714 | 0.556 |
| FT7 | 3.718 | 0.721 - 19.180 | 0.117 | 0.533 | 0.667 | 0.365 - 0.951 | 0.297 | 0.688 | 0.857 | 0.556 |
| FC3 | 1.931 | 0.572 - 6.519 | 0.289 | 0.533 | 0.460 | 0.167 - 0.794 | 0.385 | 0.625 | 0.714 | 0.556 |
| C1 | 1.963 | 0.616 - 6.254 | 0.254 | 0.533 | 0.508 | 0.207 - 0.833 | 0.394 | 0.688 | 0.857 | 0.556 |
| C5 | 2.167 | 0.622 - 7.553 | 0.225 | 0.533 | 0.492 | 0.173 - 0.815 | 0.328 | 0.563 | 0.714 | 0.444 |
| TP7 | 1.807 | 0.588 - 5.547 | 0.301 | 0.533 | 0.413 | 0.121 - 0.746 | 0.438 | 0.625 | 0.429 | 0.778 |
| CP3 | 0.979 | 0.353 - 2.717 | 0.967 | 0.983 | 0.000 | 0.000 - 0.000 | 0.000 | 0.438 | 1.000 | 0.000 |
| P1 | 1.563 | 0.531 - 4.600 | 0.418 | 0.548 | 0.333 | 0.068 - 0.655 | 0.422 | 0.500 | 0.571 | 0.444 |
| P5 | 0.941 | 0.339 - 2.616 | 0.908 | 0.939 | 0.000 | 0.000 - 0.000 | 0.000 | 0.438 | 1.000 | 0.000 |
| PO7 | 0.873 | 0.313 - 2.435 | 0.795 | 0.849 | 0.000 | 0.000 - 0.000 | 0.000 | 0.438 | 1.000 | 0.000 |
| PO3 | 0.716 | 0.254 - 2.020 | 0.528 | 0.628 | 0.302 | 0.049 - 0.650 | 0.420 | 0.500 | 0.571 | 0.444 |
| POz | 1.367 | 0.475 - 3.930 | 0.562 | 0.656 | 0.206 | 0.000 - 0.476 | 0.000 | 0.438 | 1.000 | 0.000 |
| PO4 | 1.536 | 0.515 - 4.578 | 0.441 | 0.558 | 0.365 | 0.087 - 0.667 | 0.425 | 0.500 | 0.571 | 0.444 |
| PO8 | 1.706 | 0.574 - 5.075 | 0.337 | 0.533 | 0.413 | 0.065 - 0.750 | 0.468 | 0.688 | 0.429 | 0.889 |
| P6 | 2.001 | 0.632 - 6.341 | 0.238 | 0.533 | 0.460 | 0.150 - 0.762 | 0.518 | 0.625 | 0.286 | 0.889 |
| P2 | 1.820 | 0.594 - 5.575 | 0.295 | 0.533 | 0.381 | 0.091 - 0.745 | 0.457 | 0.625 | 0.429 | 0.778 |
| CPz | 2.576 | 0.760 - 8.724 | 0.128 | 0.533 | 0.571 | 0.272 - 0.883 | 0.314 | 0.625 | 0.714 | 0.556 |
| CP4 | 1.487 | 0.518 - 4.264 | 0.461 | 0.558 | 0.302 | 0.055 - 0.633 | 0.000 | 0.438 | 1.000 | 0.000 |
| TP8 | 1.807 | 0.590 - 5.540 | 0.300 | 0.533 | 0.413 | 0.094 - 0.763 | 0.493 | 0.625 | 0.429 | 0.778 |
| C6 | 1.825 | 0.565 - 5.894 | 0.315 | 0.533 | 0.365 | 0.070 - 0.667 | 0.391 | 0.563 | 0.714 | 0.444 |
| C2 | 1.257 | 0.445 - 3.553 | 0.666 | 0.763 | 0.000 | 0.000 - 0.000 | 0.000 | 0.438 | 1.000 | 0.000 |
| FC4 | 1.639 | 0.554 - 4.850 | 0.372 | 0.536 | 0.333 | 0.055 - 0.667 | 0.494 | 0.563 | 0.286 | 0.778 |
| FT8 | 4.432 | 0.962 - 20.408 | 0.056 | 0.533 | 0.714 | 0.420 - 0.964 | 0.582 | 0.750 | 0.571 | 0.889 |
| F6 | 2.762 | 0.732 - 10.422 | 0.134 | 0.533 | 0.603 | 0.266 - 0.886 | 0.497 | 0.688 | 0.571 | 0.778 |
| AF8 | 4.387 | 0.858 - 22.443 | 0.076 | 0.533 | 0.714 | 0.383 - 1.000 | 0.643 | 0.813 | 0.571 | 1.000 |
| AF4 | 1.189 | 0.426 - 3.318 | 0.741 | 0.805 | 0.000 | 0.000 - 0.000 | 0.000 | 0.438 | 1.000 | 0.000 |
| F2 | 1.691 | 0.568 - 5.036 | 0.345 | 0.533 | 0.317 | 0.033 - 0.636 | 0.512 | 0.563 | 0.286 | 0.778 |

OR, odds ratio; CI, confidence interval; AUC, area under the curve; Cut-off, optimal threshold for classification; q value, false discovery rate–adjusted p value using the Benjamini–Hochberg procedure.

Note. Electrode refers to individual scalp EEG electrodes used in the classification analysis of baseline gamma band event-related synchronizationto predict CF increase.

Supplementary Table 6C. Predictive performance of stimulus-driven directional connectivity for CF increase based on leave-one-out cross-validation

| **ROI** | **OR** | **95% CI for**  **OR** | **p value** | **q value** | **AUC** | **95% CI for AUC** | **Cut-**  **off** | **Accuracy** | **Sensitivity** | **Specificity** |
| --- | --- | --- | --- | --- | --- | --- | --- | --- | --- | --- |
| LF-LF | 1.624 | 0.553 - 4.770 | 0.378 | 0.873 | 0.302 | 0.059 - 0.632 | 0.000 | 0.438 | 1.000 | 0.000 |
| LF-LT | 1.362 | 0.485 - 3.821 | 0.558 | 0.852 | 0.095 | 0.000 - 0.333 | 0.000 | 0.438 | 1.000 | 0.000 |
| LF-LC | 0.996 | 0.359 - 2.764 | 0.994 | 0.970 | 0.000 | 0.000 - 0.000 | 0.000 | 0.438 | 1.000 | 0.000 |
| LF-LP | 0.382 | 0.081 - 1.801 | 0.224 | 0.969 | 0.492 | 0.167 - 0.857 | 0.419 | 0.688 | 0.857 | 0.556 |
| LF-pF | 1.653 | 0.493 - 5.541 | 0.415 | 0.852 | 0.127 | 0.000 - 0.333 | 0.000 | 0.438 | 1.000 | 0.000 |
| LF-F | 1.286 | 0.461 - 3.589 | 0.631 | 0.852 | 0.000 | 0.000 - 0.000 | 0.000 | 0.438 | 1.000 | 0.000 |
| LF-C | 1.912 | 0.498 - 7.330 | 0.345 | 0.911 | 0.302 | 0.049 - 0.587 | 0.000 | 0.438 | 1.000 | 0.000 |
| LF-P | 1.726 | 0.534 - 5.577 | 0.362 | 0.941 | 0.333 | 0.083 - 0.641 | 0.381 | 0.500 | 0.714 | 0.333 |
| LF-O | 1.569 | 0.487 - 5.056 | 0.451 | 0.852 | 0.000 | 0.000 - 0.000 | 0.000 | 0.438 | 1.000 | 0.000 |
| LF-RF | 1.073 | 0.388 - 2.966 | 0.892 | 0.852 | 0.000 | 0.000 - 0.000 | 0.000 | 0.438 | 1.000 | 0.000 |
| LF-RT | 3.344 | 0.504 - 22.176 | 0.211 | 0.873 | 0.492 | 0.167 - 0.794 | 0.525 | 0.625 | 0.286 | 0.889 |
| LF-RC | 1.852 | 0.532 - 6.451 | 0.333 | 0.976 | 0.333 | 0.073 - 0.667 | 0.465 | 0.563 | 0.143 | 0.889 |
| LF-RP | 17.247 | 0.023 - 12818.978 | 0.399 | 0.897 | 0.381 | 0.063 - 0.717 | 0.715 | 0.625 | 0.143 | 1.000 |
| LT-LF | 1.313 | 0.471 - 3.663 | 0.603 | 0.852 | 0.000 | 0.000 - 0.000 | 0.000 | 0.438 | 1.000 | 0.000 |
| LT-LT | 5.848 | 0.766 - 44.643 | 0.089 | 0.852 | 0.667 | 0.322 - 0.967 | 0.656 | 0.750 | 0.429 | 1.000 |
| LT-LC | 1.294 | 0.460 - 3.641 | 0.625 | 0.874 | 0.000 | 0.000 - 0.000 | 0.000 | 0.438 | 1.000 | 0.000 |
| LT-LP | 0.484 | 0.131 - 1.789 | 0.277 | 0.955 | 0.397 | 0.109 - 0.709 | 0.330 | 0.625 | 0.857 | 0.444 |
| LT-pF | 2.338 | 0.587 - 9.315 | 0.229 | 0.873 | 0.444 | 0.108 - 0.793 | 0.387 | 0.625 | 0.571 | 0.667 |
| LT-F | 2.279 | 0.653 - 7.955 | 0.197 | 0.999 | 0.492 | 0.150 - 0.833 | 0.542 | 0.688 | 0.429 | 0.889 |
| LT-C | 1.226 | 0.440 - 3.416 | 0.696 | 0.852 | 0.000 | 0.000 - 0.000 | 0.000 | 0.438 | 1.000 | 0.000 |
| LT-P | 0.935 | 0.335 - 2.610 | 0.898 | 0.852 | 0.000 | 0.000 - 0.000 | 0.000 | 0.438 | 1.000 | 0.000 |
| LT-O | 0.949 | 0.339 - 2.656 | 0.921 | 0.852 | 0.000 | 0.000 - 0.000 | 0.000 | 0.438 | 1.000 | 0.000 |
| LT-RF | 2.056 | 0.596 - 7.088 | 0.254 | 0.852 | 0.540 | 0.220 - 0.873 | 0.385 | 0.688 | 0.714 | 0.667 |
| LT-RT | 1.637 | 0.542 - 4.939 | 0.382 | 0.852 | 0.286 | 0.033 - 0.600 | 0.396 | 0.500 | 0.571 | 0.444 |
| LT-RC | 1.848 | 0.563 - 6.061 | 0.311 | 0.852 | 0.365 | 0.055 - 0.692 | 0.430 | 0.563 | 0.429 | 0.667 |
| LT-RP | 0.934 | 0.335 - 2.608 | 0.897 | 0.870 | 0.000 | 0.000 - 0.000 | 0.000 | 0.438 | 1.000 | 0.000 |
| LC-LF | 1.411 | 0.495 - 4.020 | 0.519 | 0.852 | 0.063 | 0.000 - 0.234 | 0.000 | 0.438 | 1.000 | 0.000 |
| LC-LT | 2.179 | 0.615 - 7.717 | 0.227 | 0.892 | 0.540 | 0.206 - 0.845 | 0.349 | 0.625 | 0.857 | 0.444 |
| LC-LC | 1.322 | 0.469 - 3.726 | 0.598 | 0.852 | 0.175 | 0.000 - 0.436 | 0.000 | 0.438 | 1.000 | 0.000 |
| LC-LP | 0.740 | 0.251 - 2.184 | 0.586 | 0.852 | 0.079 | 0.000 - 0.236 | 0.000 | 0.438 | 1.000 | 0.000 |
| LC-pF | 2.640 | 0.338 - 20.641 | 0.355 | 0.873 | 0.349 | 0.000 - 0.673 | 0.597 | 0.625 | 0.143 | 1.000 |
| LC-F | 1.790 | 0.585 - 5.474 | 0.307 | 0.852 | 0.349 | 0.078 - 0.683 | 0.426 | 0.563 | 0.429 | 0.667 |
| LC-C | 0.825 | 0.291 - 2.341 | 0.718 | 0.852 | 0.000 | 0.000 - 0.000 | 0.000 | 0.438 | 1.000 | 0.000 |
| LC-P | 2.659 | 0.774 - 9.133 | 0.120 | 0.852 | 0.603 | 0.267 - 0.900 | 0.390 | 0.688 | 0.714 | 0.667 |
| LC-O | 1.110 | 0.401 - 3.069 | 0.841 | 0.852 | 0.000 | 0.000 - 0.000 | 0.000 | 0.438 | 1.000 | 0.000 |
| LC-RF | 1.687 | 0.573 - 4.965 | 0.342 | 0.852 | 0.397 | 0.098 - 0.718 | 0.407 | 0.563 | 0.571 | 0.556 |
| LC-RT | 3.521 | 0.924 - 13.418 | 0.065 | 0.852 | 0.683 | 0.381 - 0.964 | 0.379 | 0.750 | 0.714 | 0.778 |
| LC-RC | 1.147 | 0.414 - 3.177 | 0.792 | 0.873 | 0.000 | 0.000 - 0.000 | 0.000 | 0.438 | 1.000 | 0.000 |
| LC-RP | 2.343 | 0.603 - 9.100 | 0.219 | 0.908 | 0.476 | 0.159 - 0.788 | 0.344 | 0.563 | 0.714 | 0.444 |
| LP-LF | 1.238 | 0.439 - 3.496 | 0.687 | 0.873 | 0.000 | 0.000 - 0.000 | 0.000 | 0.438 | 1.000 | 0.000 |
| LP-LT | 2.101 | 0.677 - 6.523 | 0.199 | 0.999 | 0.540 | 0.236 - 0.846 | 0.436 | 0.625 | 0.571 | 0.667 |
| LP-LC | 0.927 | 0.331 - 2.595 | 0.885 | 0.852 | 0.000 | 0.000 - 0.000 | 0.000 | 0.438 | 1.000 | 0.000 |
| LP-LP | 0.614 | 0.201 - 1.876 | 0.392 | 0.852 | 0.254 | 0.000 - 0.569 | 0.345 | 0.500 | 0.714 | 0.333 |
| LP-pF | 0.810 | 0.282 - 2.326 | 0.696 | 0.852 | 0.000 | 0.000 - 0.000 | 0.000 | 0.438 | 1.000 | 0.000 |
| LP-F | 1.862 | 0.595 - 5.829 | 0.285 | 0.852 | 0.508 | 0.208 - 0.819 | 0.328 | 0.563 | 0.857 | 0.333 |
| LP-C | 0.524 | 0.113 - 2.431 | 0.409 | 0.951 | 0.222 | 0.000 - 0.544 | 0.328 | 0.500 | 0.857 | 0.222 |
| LP-P | 0.783 | 0.270 - 2.269 | 0.652 | 0.852 | 0.000 | 0.000 - 0.000 | 0.000 | 0.438 | 1.000 | 0.000 |
| LP-O | 0.274 | 0.028 - 2.666 | 0.265 | 0.870 | 0.476 | 0.188 - 0.798 | 0.455 | 0.625 | 0.571 | 0.667 |
| LP-RF | 1.409 | 0.495 - 4.009 | 0.521 | 0.852 | 0.206 | 0.000 - 0.488 | 0.000 | 0.438 | 1.000 | 0.000 |
| LP-RT | 1.343 | 0.473 - 3.807 | 0.580 | 0.873 | 0.000 | 0.000 - 0.000 | 0.000 | 0.438 | 1.000 | 0.000 |
| LP-RC | 1.068 | 0.385 - 2.962 | 0.899 | 0.891 | 0.000 | 0.000 - 0.000 | 0.000 | 0.438 | 1.000 | 0.000 |
| LP-RP | 0.368 | 0.100 - 1.359 | 0.134 | 0.897 | 0.603 | 0.286 - 0.906 | 0.412 | 0.688 | 0.714 | 0.667 |
| pF-LF | 1.585 | 0.552 - 4.555 | 0.392 | 0.852 | 0.365 | 0.087 - 0.716 | 0.411 | 0.563 | 0.571 | 0.556 |
| pF-LT | 2.019 | 0.635 - 6.417 | 0.234 | 0.873 | 0.349 | 0.000 - 0.711 | 0.392 | 0.625 | 0.429 | 0.778 |
| pF-LC | 0.564 | 0.148 - 2.143 | 0.400 | 0.852 | 0.270 | 0.033 - 0.578 | 0.000 | 0.438 | 1.000 | 0.000 |
| pF-LP | 0.393 | 0.098 - 1.585 | 0.189 | 0.852 | 0.460 | 0.154 - 0.784 | 0.399 | 0.625 | 0.857 | 0.444 |
| pF-pF | 1.373 | 0.473 - 3.982 | 0.560 | 0.852 | 0.000 | 0.000 - 0.000 | 0.000 | 0.438 | 1.000 | 0.000 |
| pF-F | 0.955 | 0.341 - 2.671 | 0.929 | 0.873 | 0.000 | 0.000 - 0.000 | 0.000 | 0.438 | 1.000 | 0.000 |
| pF-C | 1.138 | 0.410 - 3.156 | 0.804 | 0.891 | 0.000 | 0.000 - 0.000 | 0.000 | 0.438 | 1.000 | 0.000 |
| pF-P | 0.708 | 0.247 - 2.032 | 0.521 | 0.852 | 0.254 | 0.033 - 0.544 | 0.000 | 0.438 | 1.000 | 0.000 |
| pF-O | 0.833 | 0.288 - 2.409 | 0.736 | 0.852 | 0.000 | 0.000 - 0.000 | 0.000 | 0.438 | 1.000 | 0.000 |
| pF-RF | 1.051 | 0.380 - 2.909 | 0.923 | 0.852 | 0.000 | 0.000 - 0.000 | 0.000 | 0.438 | 1.000 | 0.000 |
| pF-RT | 1.553 | 0.538 - 4.479 | 0.416 | 0.955 | 0.333 | 0.078 - 0.650 | 0.377 | 0.500 | 0.714 | 0.333 |
| pF-RC | 1.112 | 0.401 - 3.083 | 0.838 | 0.955 | 0.000 | 0.000 - 0.000 | 0.000 | 0.438 | 1.000 | 0.000 |
| pF-RP | 1.039 | 0.376 - 2.873 | 0.941 | 0.969 | 0.000 | 0.000 - 0.000 | 0.000 | 0.438 | 1.000 | 0.000 |
| F-LF | 0.811 | 0.275 - 2.392 | 0.704 | 0.852 | 0.000 | 0.000 - 0.000 | 0.000 | 0.438 | 1.000 | 0.000 |
| F-LT | 1.365 | 0.485 - 3.840 | 0.556 | 0.873 | 0.079 | 0.000 - 0.254 | 0.000 | 0.438 | 1.000 | 0.000 |
| F-LC | 0.133 | 0.001 - 18.103 | 0.421 | 0.852 | 0.413 | 0.121 - 0.730 | 0.021 | 0.563 | 1.000 | 0.222 |
| F-LP | 0.067 | 0.001 - 4.340 | 0.204 | 0.852 | 0.556 | 0.262 - 0.857 | 0.451 | 0.688 | 0.857 | 0.556 |
| F-pF | 4.648 | 0.163 - 132.690 | 0.369 | 0.852 | 0.444 | 0.133 - 0.750 | 0.660 | 0.625 | 0.143 | 1.000 |
| F-F | 1.708 | 0.554 - 5.265 | 0.351 | 0.873 | 0.302 | 0.000 - 0.000 | 0.457 | 0.625 | 0.286 | 0.889 |
| F-C | 1.573 | 0.539 - 4.589 | 0.407 | 0.892 | 0.254 | 0.000 - 0.582 | 0.000 | 0.438 | 1.000 | 0.000 |
| F-P | 2.018 | 0.630 - 6.463 | 0.237 | 0.908 | 0.460 | 0.167 - 0.763 | 0.547 | 0.625 | 0.286 | 0.889 |
| F-O | 1.160 | 0.419 - 3.211 | 0.775 | 0.852 | 0.000 | 0.000 - 0.000 | 0.000 | 0.438 | 1.000 | 0.000 |
| F-RF | 6.752 | 0.656 - 69.477 | 0.108 | 0.873 | 0.762 | 0.467 - 1.000 | 0.448 | 0.813 | 0.714 | 0.889 |
| F-RT | 1.717 | 0.588 - 5.011 | 0.323 | 0.852 | 0.397 | 0.094 - 0.700 | 0.373 | 0.563 | 0.571 | 0.556 |
| F-RC | 1.936 | 0.643 - 5.829 | 0.240 | 0.852 | 0.524 | 0.207 - 0.843 | 0.375 | 0.688 | 0.714 | 0.667 |
| F-RP | 3.885 | 0.083 - 182.161 | 0.489 | 0.941 | 0.413 | 0.133 - 0.763 | 0.799 | 0.625 | 0.143 | 1.000 |
| C-LF | 3.759 | 0.706 - 20.020 | 0.121 | 0.852 | 0.587 | 0.252 - 0.902 | 0.583 | 0.750 | 0.429 | 1.000 |
| C-LT | 1.228 | 0.441 - 3.421 | 0.694 | 0.852 | 0.000 | 0.000 - 0.000 | 0.000 | 0.438 | 1.000 | 0.000 |
| C-LC | 0.777 | 0.263 - 2.291 | 0.648 | 0.873 | 0.000 | 0.000 - 0.000 | 0.000 | 0.438 | 1.000 | 0.000 |
| C-LP | 0.574 | 0.166 - 1.983 | 0.380 | 0.897 | 0.349 | 0.079 - 0.667 | 0.429 | 0.563 | 0.714 | 0.444 |
| C-pF | 1.714 | 0.578 - 5.085 | 0.331 | 0.891 | 0.317 | 0.000 - 0.667 | 0.423 | 0.563 | 0.429 | 0.667 |
| C-F | 1.503 | 0.510 - 4.431 | 0.460 | 0.889 | 0.175 | 0.000 - 0.435 | 0.000 | 0.438 | 1.000 | 0.000 |
| C-C | 0.422 | 0.095 - 1.867 | 0.255 | 0.852 | 0.413 | 0.114 - 0.745 | 0.319 | 0.625 | 1.000 | 0.333 |
| C-P | 0.744 | 0.255 - 2.175 | 0.589 | 0.873 | 0.016 | 0.000 - 0.095 | 0.000 | 0.438 | 1.000 | 0.000 |
| C-O | 0.728 | 0.249 - 2.126 | 0.561 | 0.852 | 0.175 | 0.000 - 0.437 | 0.000 | 0.438 | 1.000 | 0.000 |
| C-RF | 1.189 | 0.428 - 3.302 | 0.740 | 0.852 | 0.000 | 0.000 - 0.000 | 0.000 | 0.438 | 1.000 | 0.000 |
| C-RT | 3.955 | 0.838 - 18.668 | 0.082 | 0.873 | 0.667 | 0.299 - 0.968 | 0.349 | 0.750 | 0.714 | 0.778 |
| C-RC | 0.686 | 0.223 - 2.104 | 0.509 | 0.873 | 0.175 | 0.000 - 0.417 | 0.000 | 0.438 | 1.000 | 0.000 |
| C-RP | 0.717 | 0.243 - 2.118 | 0.548 | 0.873 | 0.111 | 0.000 - 0.333 | 0.000 | 0.438 | 1.000 | 0.000 |
| P-LF | 1.318 | 0.465 - 3.737 | 0.603 | 0.955 | 0.032 | 0.000 - 0.144 | 0.000 | 0.438 | 1.000 | 0.000 |
| P-LT | 1.448 | 0.507 - 4.137 | 0.490 | 0.852 | 0.143 | 0.000 - 0.400 | 0.000 | 0.438 | 1.000 | 0.000 |
| P-LC | 1.021 | 0.369 - 2.829 | 0.968 | 0.852 | 0.000 | 0.000 - 0.000 | 0.000 | 0.438 | 1.000 | 0.000 |
| P-LP | 1.144 | 0.414 - 3.164 | 0.795 | 0.897 | 0.000 | 0.000 - 0.000 | 0.000 | 0.438 | 1.000 | 0.000 |
| P-pF | 3.777 | 0.377 - 37.843 | 0.258 | 0.892 | 0.444 | 0.121 - 0.800 | 0.615 | 0.688 | 0.286 | 1.000 |
| P-F | 1.907 | 0.551 - 6.596 | 0.308 | 0.979 | 0.381 | 0.095 - 0.718 | 0.402 | 0.563 | 0.429 | 0.667 |
| P-C | 0.456 | 0.109 - 1.903 | 0.281 | 0.852 | 0.460 | 0.164 - 0.783 | 0.440 | 0.625 | 0.714 | 0.556 |
| P-P | 0.715 | 0.220 - 2.322 | 0.577 | 0.852 | 0.032 | 0.000 - 0.141 | 0.000 | 0.438 | 1.000 | 0.000 |
| P-O | 0.911 | 0.323 - 2.570 | 0.861 | 0.852 | 0.000 | 0.000 - 0.000 | 0.000 | 0.438 | 1.000 | 0.000 |
| P-RF | 6.035 | 1.032 - 35.282 | 0.046 | 0.852 | 0.746 | 0.400 - 1.000 | 0.343 | 0.813 | 0.857 | 0.778 |
| P-RT | 3.674 | 0.484 - 27.885 | 0.208 | 0.873 | 0.571 | 0.235 - 0.854 | 0.494 | 0.625 | 0.429 | 0.778 |
| P-RC | 1.299 | 0.463 - 3.644 | 0.619 | 0.852 | 0.000 | 0.000 - 0.000 | 0.000 | 0.438 | 1.000 | 0.000 |
| P-RP | 1.539 | 0.538 - 4.403 | 0.422 | 0.873 | 0.238 | 0.000 - 0.556 | 0.000 | 0.438 | 1.000 | 0.000 |
| O-LF | 1.227 | 0.441 - 3.419 | 0.695 | 0.873 | 0.000 | 0.000 - 0.000 | 0.000 | 0.438 | 1.000 | 0.000 |
| O-LT | 2.452 | 0.524 - 11.480 | 0.255 | 0.873 | 0.460 | 0.129 - 0.800 | 0.504 | 0.625 | 0.429 | 0.778 |
| O-LC | 1.178 | 0.424 - 3.275 | 0.753 | 0.852 | 0.000 | 0.000 - 0.000 | 0.000 | 0.438 | 1.000 | 0.000 |
| O-LP | 0.861 | 0.307 - 2.421 | 0.777 | 0.979 | 0.000 | 0.000 - 0.000 | 0.000 | 0.438 | 1.000 | 0.000 |
| O-pF | 3.140 | 0.730 - 13.505 | 0.124 | 0.873 | 0.524 | 0.190 - 0.858 | 0.490 | 0.688 | 0.429 | 0.889 |
| O-F | 1.295 | 0.465 - 3.607 | 0.620 | 0.891 | 0.000 | 0.000 - 0.000 | 0.000 | 0.438 | 1.000 | 0.000 |
| O-C | 0.462 | 0.116 - 1.837 | 0.273 | 0.852 | 0.381 | 0.100 - 0.700 | 0.254 | 0.625 | 1.000 | 0.333 |
| O-P | 0.821 | 0.282 - 2.393 | 0.718 | 0.852 | 0.000 | 0.000 - 0.000 | 0.000 | 0.438 | 1.000 | 0.000 |
| O-O | 0.689 | 0.243 - 1.951 | 0.483 | 0.897 | 0.349 | 0.083 - 0.667 | 0.421 | 0.563 | 0.571 | 0.556 |
| O-RF | 10.730 | 0.958 - 120.200 | 0.054 | 0.852 | 0.746 | 0.460 - 0.980 | 0.330 | 0.750 | 0.714 | 0.778 |
| O-RT | 1.145 | 0.413 - 3.173 | 0.794 | 0.908 | 0.000 | 0.000 - 0.000 | 0.000 | 0.438 | 1.000 | 0.000 |
| O-RC | 1.521 | 0.533 - 4.338 | 0.433 | 0.852 | 0.333 | 0.000 - 0.667 | 0.434 | 0.625 | 0.429 | 0.778 |
| O-RP | 0.590 | 0.203 - 1.711 | 0.331 | 0.873 | 0.365 | 0.083 - 0.700 | 0.391 | 0.563 | 0.571 | 0.556 |
| RF-LF | 0.995 | 0.358 - 2.760 | 0.992 | 0.891 | 0.000 | 0.000 - 0.000 | 0.000 | 0.438 | 1.000 | 0.000 |
| RF-LT | 2.286 | 0.722 - 7.241 | 0.160 | 0.891 | 0.492 | 0.150 - 0.857 | 0.418 | 0.688 | 0.571 | 0.778 |
| RF-LC | 0.543 | 0.141 - 2.087 | 0.374 | 0.852 | 0.444 | 0.165 - 0.765 | 0.442 | 0.563 | 0.571 | 0.556 |
| RF-LP | 0.110 | 0.003 - 3.824 | 0.223 | 0.873 | 0.508 | 0.195 - 0.831 | 0.238 | 0.625 | 1.000 | 0.333 |
| RF-pF | 1.515 | 0.471 - 4.875 | 0.486 | 0.852 | 0.000 | 0.000 - 0.000 | 0.000 | 0.438 | 1.000 | 0.000 |
| RF-F | 2.402 | 0.681 - 8.475 | 0.173 | 0.955 | 0.476 | 0.143 - 0.825 | 0.659 | 0.688 | 0.286 | 1.000 |
| RF-C | 1.099 | 0.396 - 3.049 | 0.857 | 0.852 | 0.000 | 0.000 - 0.000 | 0.000 | 0.438 | 1.000 | 0.000 |
| RF-P | 0.738 | 0.255 - 2.136 | 0.575 | 0.955 | 0.159 | 0.000 - 0.403 | 0.000 | 0.438 | 1.000 | 0.000 |
| RF-O | 0.840 | 0.287 - 2.458 | 0.751 | 0.873 | 0.000 | 0.000 - 0.000 | 0.000 | 0.438 | 1.000 | 0.000 |
| RF-RF | 1.742 | 0.551 - 5.502 | 0.344 | 0.852 | 0.349 | 0.000 - 0.716 | 0.436 | 0.625 | 0.429 | 0.778 |
| RF-RT | 1.494 | 0.523 - 4.269 | 0.453 | 0.889 | 0.317 | 0.070 - 0.638 | 0.403 | 0.500 | 0.571 | 0.444 |
| RF-RC | 2.032 | 0.654 - 6.313 | 0.220 | 0.852 | 0.492 | 0.172 - 0.795 | 0.415 | 0.625 | 0.571 | 0.667 |
| RF-RP | 1.238 | 0.440 - 3.486 | 0.686 | 0.852 | 0.000 | 0.000 - 0.000 | 0.000 | 0.438 | 1.000 | 0.000 |
| RT-LF | 1.323 | 0.460 - 3.806 | 0.604 | 0.852 | 0.000 | 0.000 - 0.000 | 0.000 | 0.438 | 1.000 | 0.000 |
| RT-LT | 0.733 | 0.219 - 2.452 | 0.615 | 0.873 | 0.000 | 0.000 - 0.000 | 0.000 | 0.438 | 1.000 | 0.000 |
| RT-LC | 0.798 | 0.284 - 2.244 | 0.669 | 0.852 | 0.000 | 0.000 - 0.000 | 0.000 | 0.438 | 1.000 | 0.000 |
| RT-LP | 0.383 | 0.062 - 2.379 | 0.303 | 0.852 | 0.444 | 0.154 - 0.762 | 0.366 | 0.563 | 0.857 | 0.333 |
| RT-pF | 0.680 | 0.228 - 2.023 | 0.488 | 0.873 | 0.286 | 0.063 - 0.583 | 0.434 | 0.500 | 0.571 | 0.444 |
| RT-F | 1.728 | 0.518 - 5.764 | 0.373 | 0.985 | 0.127 | 0.000 - 0.429 | 0.489 | 0.563 | 0.143 | 0.889 |
| RT-C | 0.477 | 0.135 - 1.689 | 0.251 | 0.852 | 0.460 | 0.143 - 0.780 | 0.275 | 0.563 | 1.000 | 0.222 |
| RT-P | 0.873 | 0.313 - 2.437 | 0.795 | 0.873 | 0.000 | 0.000 - 0.000 | 0.000 | 0.438 | 1.000 | 0.000 |
| RT-O | 0.730 | 0.248 - 2.150 | 0.568 | 0.852 | 0.238 | 0.000 - 0.504 | 0.426 | 0.500 | 0.714 | 0.333 |
| RT-RF | 1.029 | 0.371 - 2.853 | 0.956 | 0.908 | 0.000 | 0.000 - 0.000 | 0.000 | 0.438 | 1.000 | 0.000 |
| RT-RT | 1.180 | 0.425 - 3.273 | 0.751 | 0.873 | 0.000 | 0.000 - 0.000 | 0.000 | 0.438 | 1.000 | 0.000 |
| RT-RC | 2.262 | 0.698 - 7.330 | 0.173 | 0.852 | 0.571 | 0.254 - 0.867 | 0.426 | 0.688 | 0.429 | 0.889 |
| RT-RP | 2.169 | 0.480 - 9.805 | 0.315 | 0.951 | 0.270 | 0.000 - 0.650 | 0.499 | 0.625 | 0.286 | 0.889 |
| RC-LF | 2.672 | 0.341 - 20.914 | 0.349 | 0.852 | 0.365 | 0.073 - 0.705 | 0.804 | 0.625 | 0.143 | 1.000 |
| RC-LT | 1.217 | 0.437 - 3.395 | 0.707 | 0.873 | 0.000 | 0.000 - 0.000 | 0.000 | 0.438 | 1.000 | 0.000 |
| RC-LC | 0.969 | 0.349 - 2.694 | 0.952 | 0.873 | 0.000 | 0.000 - 0.000 | 0.000 | 0.438 | 1.000 | 0.000 |
| RC-LP | 0.315 | 0.030 - 3.337 | 0.337 | 0.852 | 0.413 | 0.143 - 0.723 | 0.485 | 0.563 | 0.571 | 0.556 |
| RC-pF | 1.081 | 0.391 - 2.988 | 0.880 | 0.852 | 0.000 | 0.000 - 0.000 | 0.000 | 0.438 | 1.000 | 0.000 |
| RC-F | 2.124 | 0.339 - 13.308 | 0.421 | 0.852 | 0.079 | 0.000 - 0.286 | 0.000 | 0.438 | 1.000 | 0.000 |
| RC-C | 0.477 | 0.127 - 1.797 | 0.274 | 0.852 | 0.444 | 0.125 - 0.778 | 0.317 | 0.688 | 1.000 | 0.444 |
| RC-P | 0.729 | 0.251 - 2.117 | 0.562 | 0.852 | 0.206 | 0.016 - 0.474 | 0.000 | 0.438 | 1.000 | 0.000 |
| RC-O | 0.749 | 0.252 - 2.221 | 0.602 | 0.852 | 0.032 | 0.000 - 0.143 | 0.000 | 0.438 | 1.000 | 0.000 |
| RC-RF | 1.185 | 0.428 - 3.282 | 0.744 | 0.852 | 0.000 | 0.000 - 0.000 | 0.000 | 0.438 | 1.000 | 0.000 |
| RC-RT | 2.645 | 0.635 - 11.019 | 0.182 | 0.891 | 0.524 | 0.186 - 0.867 | 0.575 | 0.688 | 0.429 | 0.889 |
| RC-RC | 2.631 | 0.585 - 11.825 | 0.207 | 0.999 | 0.460 | 0.127 - 0.810 | 0.575 | 0.688 | 0.286 | 1.000 |
| RC-RP | 1.618 | 0.488 - 5.365 | 0.431 | 0.852 | 0.079 | 0.000 - 0.267 | 0.000 | 0.438 | 1.000 | 0.000 |
| RP-LF | 0.767 | 0.265 - 2.221 | 0.625 | 0.852 | 0.000 | 0.000 - 0.000 | 0.000 | 0.438 | 1.000 | 0.000 |
| RP-LT | 9.210 | 0.471 - 180.104 | 0.143 | 0.891 | 0.651 | 0.286 - 0.967 | 0.448 | 0.750 | 0.714 | 0.778 |
| RP-LC | 2.440 | 0.492 - 12.106 | 0.275 | 0.873 | 0.397 | 0.000 - 0.764 | 0.485 | 0.688 | 0.429 | 0.889 |
| RP-LP | 0.634 | 0.203 - 1.982 | 0.433 | 0.852 | 0.302 | 0.063 - 0.604 | 0.000 | 0.438 | 1.000 | 0.000 |
| RP-pF | 2.080 | 0.407 - 10.644 | 0.379 | 0.852 | 0.111 | 0.000 - 0.375 | 0.000 | 0.438 | 1.000 | 0.000 |
| RP-F | 1.437 | 0.498 - 4.144 | 0.502 | 0.897 | 0.095 | 0.000 - 0.333 | 0.000 | 0.438 | 1.000 | 0.000 |
| RP-C | 0.343 | 0.067 - 1.744 | 0.197 | 0.852 | 0.524 | 0.200 - 0.874 | 0.377 | 0.688 | 0.857 | 0.556 |
| RP-P | 0.800 | 0.280 - 2.283 | 0.677 | 0.852 | 0.000 | 0.000 - 0.000 | 0.000 | 0.438 | 1.000 | 0.000 |
| RP-O | 0.581 | 0.176 - 1.921 | 0.374 | 0.908 | 0.349 | 0.092 - 0.656 | 0.312 | 0.500 | 0.857 | 0.222 |
| RP-RF | 2.103 | 0.614 - 7.207 | 0.237 | 0.908 | 0.492 | 0.190 - 0.798 | 0.372 | 0.625 | 0.714 | 0.556 |
| RP-RT | 4.537 | 0.207 - 99.534 | 0.337 | 0.892 | 0.476 | 0.164 - 0.766 | 0.335 | 0.563 | 0.714 | 0.444 |
| RP-RC | 1.809 | 0.436 - 7.511 | 0.415 | 0.852 | 0.063 | 0.000 - 0.218 | 0.000 | 0.438 | 1.000 | 0.000 |
| RP-RP | 0.999 | 0.360 - 2.772 | 0.999 | 0.873 | 0.000 | 0.000 - 0.000 | 0.000 | 0.438 | 1.000 | 0.000 |

OR, odds ratio; CI, confidence interval; AUC, area under the curve; Cut-off, optimal threshold for classification; q value, false discovery rate–adjusted p value using the Benjamini–Hochberg procedure.

Note. ROI indicates pairs of brain regions analyzed for stimulus-driven directional connectivity (ΔsGC) to predict CF increase after intervention.
